# Interventions to control nosocomial transmission of SARS-CoV-2: a modelling study

**DOI:** 10.1101/2021.02.26.21252327

**Authors:** Thi Mui Pham, Hannan Tahir, Janneke H.H.M. van de Wijgert, Bastiaan Van der Roest, Pauline Ellerbroek, Marc J.M. Bonten, Martin C.J. Bootsma, Mirjam E. Kretzschmar

## Abstract

**Background:** Emergence of more transmissible SARS-CoV-2 variants requires more efficient control measures to limit nosocomial transmission and maintain healthcare capacities during pandemic waves. Yet, the relative importance of different strategies is unknown.

**Methods:** We developed an agent-based model and compared the impact of personal protective equipment (PPE), screening of healthcare workers (HCWs), contact tracing of symptomatic HCWs, and restricting HCWs from working in multiple units (HCW cohorting) on nosocomial SARS-CoV-2 transmission. The model was fit on hospital data from the first wave in the Netherlands (February until August 2020) and assumed that HCWs used 90% effective PPE in COVID-19 wards and self-isolated at home for seven days immediately upon symptom onset. Intervention effects on the effective reproduction number (*R*_*E*_), HCW absenteeism and the proportion of infected individuals among tested individuals (positivity rate) were estimated for a more transmissible variant.

**Results:** Introduction of a variant with 56% higher transmissibility increased – all other variables kept constant – *R*_*E*_ from 0.4 to 0.65 (+63%) and nosocomial transmissions by 303%, mainly because of more transmissions caused by pre-symptomatic patients and HCWs. Compared to baseline, PPE use in all hospital wards (assuming 90% effectiveness) reduced *R*_*E*_ by 85% and absenteeism by 57%. Screening HCWs every three days with perfect test sensitivity reduced *R*_*E*_ by 67%, yielding a maximum test positivity rate of 5%. Screening HCWs every three or seven days assuming time-varying test sensitivities reduced *R*_*E*_ by 9% and 3%, respectively. Contact tracing reduced *R*_*E*_ by at least 32% and achieved higher test positivity rates than screening interventions. HCW cohorting reduced *R*_*E*_ by 5%. Sensitivity analyses for 50% and 70% effectiveness of PPE use did not change interpretation.

**Conclusions:** In response to the emergence of more transmissible SARS-CoV-2 variants, PPE use in all hospital wards might still be most effective in preventing nosocomial transmission. Regular screening and contact tracing of HCWs are also effective interventions, but critically depend on the sensitivity of the diagnostic test used.

## Background

Effective interventions to limit nosocomial transmission of the severe acute respiratory syndrome coronavirus 2 (SARS-CoV-2) are pivotal to maintain healthcare capacities during pandemic waves [1,2]. During the first epidemic wave many hospitals around the world restricted visits and canceled non-essential medical procedures in order to maintain adequate staffing levels for patients with COVID-19. In the Netherlands, specific infection control measures were implemented but nosocomial transmission may have been facilitated by temporary shortness of supplies of personal protective equipment (PPE), including gloves, goggles, face shields, gowns, and (N95) masks, at the onset of the pandemic.

Indeed, HCWs experienced a higher incidence of SARS-CoV-2 infections, compared to other professions, during the first pandemic wave [3–5]. Front-line HCWs in the UK and USA tested three times more frequently positive during the first epidemic wave than the general population after accounting for the frequency of testing [3]. Other studies from the UK and the Netherlands found higher SARS-CoV-2 incidences after the first epidemic wave among staff working in COVID-19 wards than staff working elsewhere in the hospital [5,6]. In addition to direct contact with infectious patients, HCW-to-HCW transmission most likely also contributed to these elevated incidence rates.

Only a few studies incorporated modelling of SARS-CoV-2 transmission in healthcare settings [7–11]. In a stochastic within-hospital model, combined with a deterministic model reflecting SARS-CoV-2 transmission in the community, PPE use by HCWs and patients in the entire hospital substantially reduced nosocomial infections, while random weekly testing of asymptomatic HCWs and patients was less effective [9]. Moreover, strict cohorting of undiagnosed patients and HCWs in small units reduced the probability that SARS-CoV-2 introduction would lead to a large outbreak. In a deterministic within-hospital Susceptible-Exposed-Infectious-Recovered (SEIR) model isolating COVID-19 patients in single rooms or bays reduced infection acquisition in patients by up to 80% [8]. The model predicted that periodic testing of HWCs would have a smaller effect on the COVID-19 patient-burden than isolating patients but could reduce HCW infections by up to 64% and lead to a reduction of staff absenteeism. Both aforementioned models assumed a time-invariant SARS-CoV-2 infectiousness and diagnostic PCR test with 100% sensitivity. An individual-based modelling study assessed the impact of different interventions for SARS-CoV-2 transmission in a non-COVID-19 hospital unit [11]. The model was calibrated to COVID-19 outbreak data in a neurosurgery hospital unit in Wuhan (January until February 2020). High-efficacy face-masks were shown to be most effective for reducing infection cases and workday loss. Reduction of contact rates had only a marginal effect on mitigating the outbreak in the long run. Another model (stochastic, individual-based, aimed at patients and HCWs in long-term care facilities (LTCF)) did incorporate a test sensitivity that varies with time since infection [7]. This model concluded that pooled testing (combining clinical specimens from multiple individuals into a single biological sample for a single RT-PCR test) was the most effective and efficient surveillance strategy for resource-limited LTCFs.

While these previous studies investigated interventions such as the PPE use, social distancing among HCWs, various testing strategies, and cohorting of patients and HCWs, the impact of contact tracing within hospital settings has not been modeled yet. Observational evidence from 5,700 HCWs in two large hospitals and 40 outpatient units in Milan, Italy, suggested that random testing (positivity rate of 2·6%) was less efficient than contact tracing (10%) [12].

In Dutch hospitals patients and HCWs were cohorted in COVID-wards, where HCWs used PPE during patient care, in addition to the basic infection control measures applied. With these measures, nosocomial transmission was considered well-controlled during the first wave of the pandemic, although outbreaks have been reported sporadically [13]. Yet, with the emergence of more transmissible variants, current infection control measures may become less effective. While COVID-19 vaccine rollout is underway, it is still unclear how they affect transmission and how their efficacy is affected by the new SARS-CoV-2 variants. We, therefore, explored the relative effectiveness of different infection prevention strategies for HCWs in hospitals in the absence of vaccination using an agent-based model of nosocomial SARS-CoV-2 transmission. First, we fitted the model to real-life data from the University Medical Center Utrecht (UMCU) during the period February-August 2020. Next, we evaluated the impact of various interventions on transmission, HCW absenteeism and test positivity as a marker of intervention efficiency for a more transmissible variant (e.g., B.1.1.7) and draw general conclusions for infection control in hospitals with a similar structure.

## Methods

### Agent-based model

We developed an agent-based model that describes the dynamics of SARS-CoV-2 transmission in a hospital allowing for importations of infections from the community (Fig 1A). We modeled a hospital comprising four ward types: 1) general COVID wards, 2) general non-COVID wards, 3) COVID intensive-care units (ICUs), and 4) non-COVID ICUs. Within the hospital we distinguish patients, nurses, and doctors. Patients are assumed to occupy a hospital bed in a single room. HCWs (nurses and doctors) work in duty shifts. HCWs meet patients in a number of rounds per shift (Appendix Table 1), and HCWs meet other HCWs in the common staff room of each ward.

**Fig 1.**
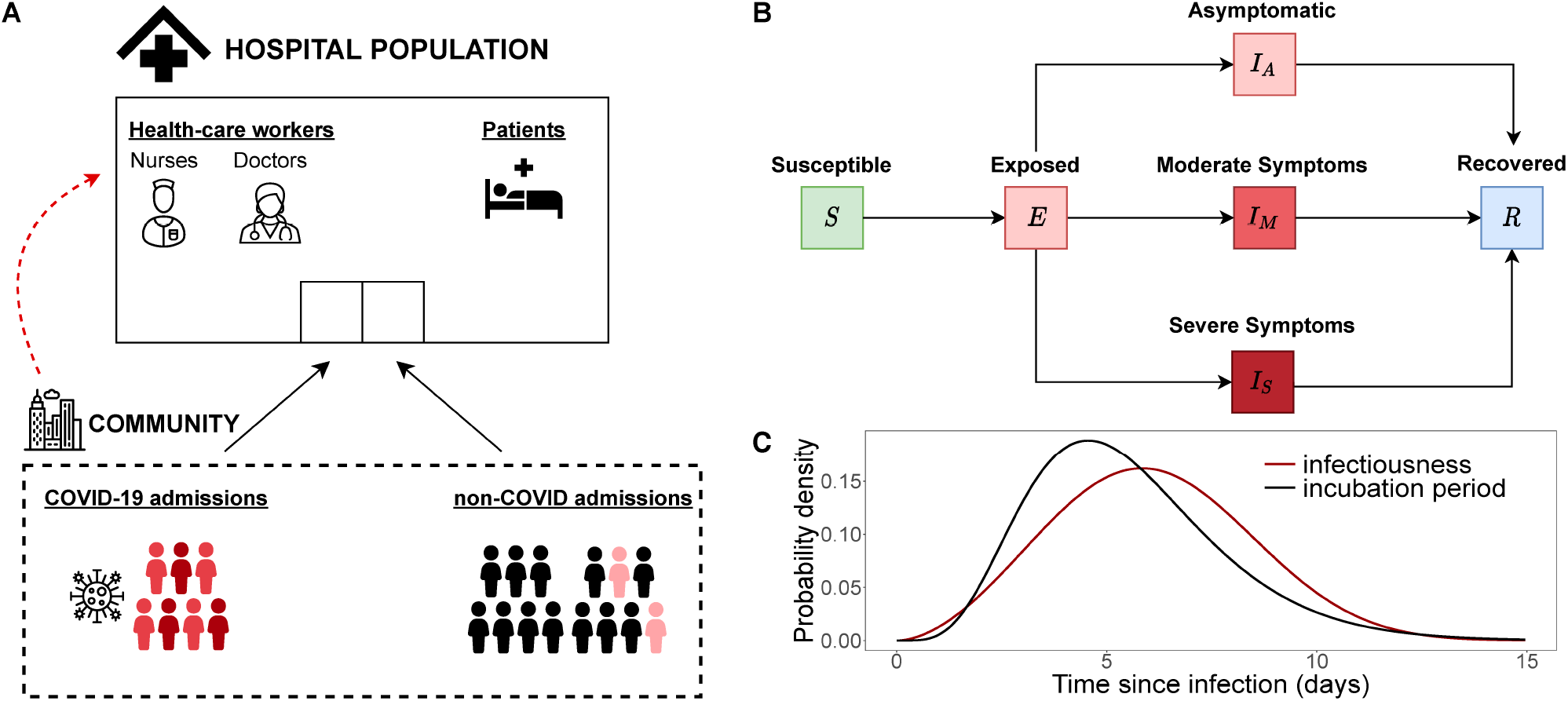
Schematics for agent-based model. (A) Diagram of the agent-based model including the agents in the main environment (hospital) and community importations. The hospital population is divided into healthcare workers (nurses and doctors) and patients. Patients may be admitted from the community either with moderate (red) or severe (dark red) COVID-19 symptoms or for non-COVID reasons. Patients may be in a pre-symptomatic stage (light red) when hospitalized to non-COVID wards. Healthcare workers may get infected in the community (red dashed line). (B) Disease progression diagram. Individuals are in either of the following categories: Susceptible (S), Exposed (E), Asymptomatically Infected (*I*_*A*_), Moderately infected (*I*_*M*_), Severely infected (*I*_*S*_), and Recovered (*R*). (C) Probability density of infectiousness of an infected individual and incubation period over time since infection.

Individuals may be in one of the disease states: susceptible (S), exposed (E), asymptomatically infected (IA), infected with moderate symptoms (IM), infected with severe symptoms (IS), and recovered (IR). We did not explicitly model other respiratory tract infections with similar symptoms. Hence, all symptomatic individuals are necessarily infected with SARS-CoV-2. We did not model death in our simulations. Patients may be admitted to the hospital for non-COVID reasons or with moderate or severe COVID-19 symptoms. In the first case, they may be susceptible, pre-symptomatically, or asymptomatically infected. Symptomatically infected patients are admitted to COVID wards (moderate symptoms) or COVID ICUs (severe symptoms). Patients in non-COVID wards that develop symptoms during their stay are immediately transferred to COVID wards. We assumed that moderately and severely infected patients recover after 14 and 35 days, respectively [14].

Transmission events can occur between patients and HCWs, and among HCWs. We assumed no patient-to-patient transmission as patients are assumed to occupy single-bed rooms. Only HCWs in their asymptomatic or pre-symptomatic phase contribute to transmission. The reproduction number (average number of secondary cases caused by an infected individual) is assumed to differ between symptomatically (*R*_*S*_) and asymptomatically (*R*_*A*_) infected individuals. We assumed that the incubation period has a Gamma distribution with mean 5·5 days and that the individual’s infectiousness over time has a Weibull distribution with a mean of 6 days (Fig 1C) [15,16].

### Data and parametrization

We used data from the UMCU to parametrize the number of wards and beds per ward (Appendix pp. 2). We used the number of patients admitted to the UMCU for non-COVID reasons and their length of stay for the time period 2014-2017 and assumed a 50% decrease in admissions during the study period (Appendix Table 1). The daily number of COVID-19 hospitalizations and their length of stay distribution was based on UMCU data from 27 February until 24 August 2020. The simulations started on 30 December 2019 with a hospital at 100% occupancy without any SARS-CoV-2-infected individuals.

The first COVID-19 admissions occurred on 27 February 2020. To account for admissions of patients that are infected but not (yet) symptomatic and HCWs who were (unknowingly) infected in the community, we used daily national numbers of SARS-CoV2 infectious individuals estimated by the Dutch National Institute for Public Health and the Environment (RIVM) from 17 February until 24 August 2020 (Appendix pp. 2) [17]. We additionally used publicly available age-specific hospitalization rates in the Netherlands in 2012 and age-specific SARS-CoV-2 infection incidence rates in Utrecht province to scale the daily probability of being infected in the community for non-COVID patients and HCWs arriving in the hospital [18,19].

Based on a published meta-analysis, we assumed that 20% and 31% of SARS-CoV-2 infections in patients and HCWs, respectively, were asymptomatic (Table 1) [20].

**Table 1.**
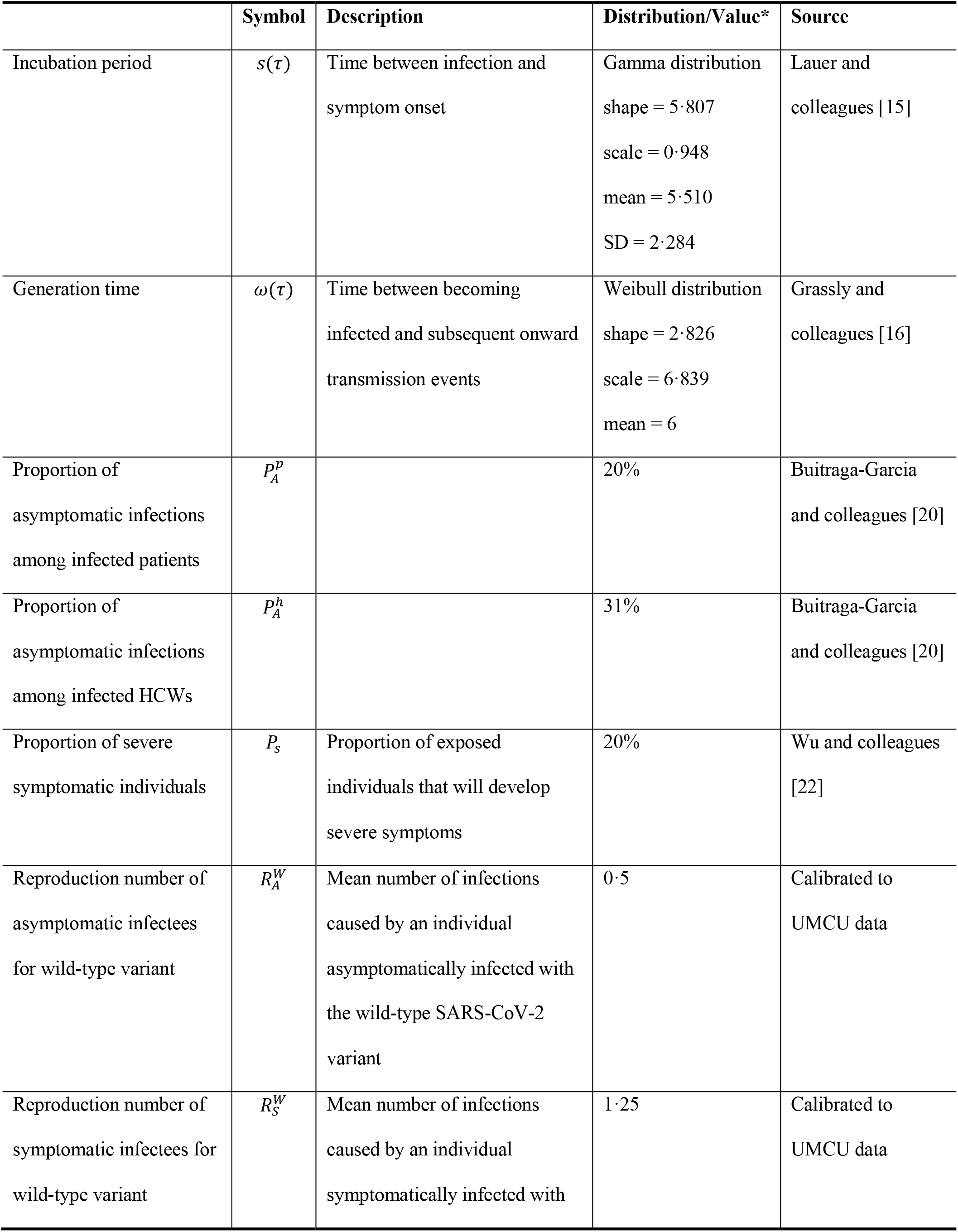
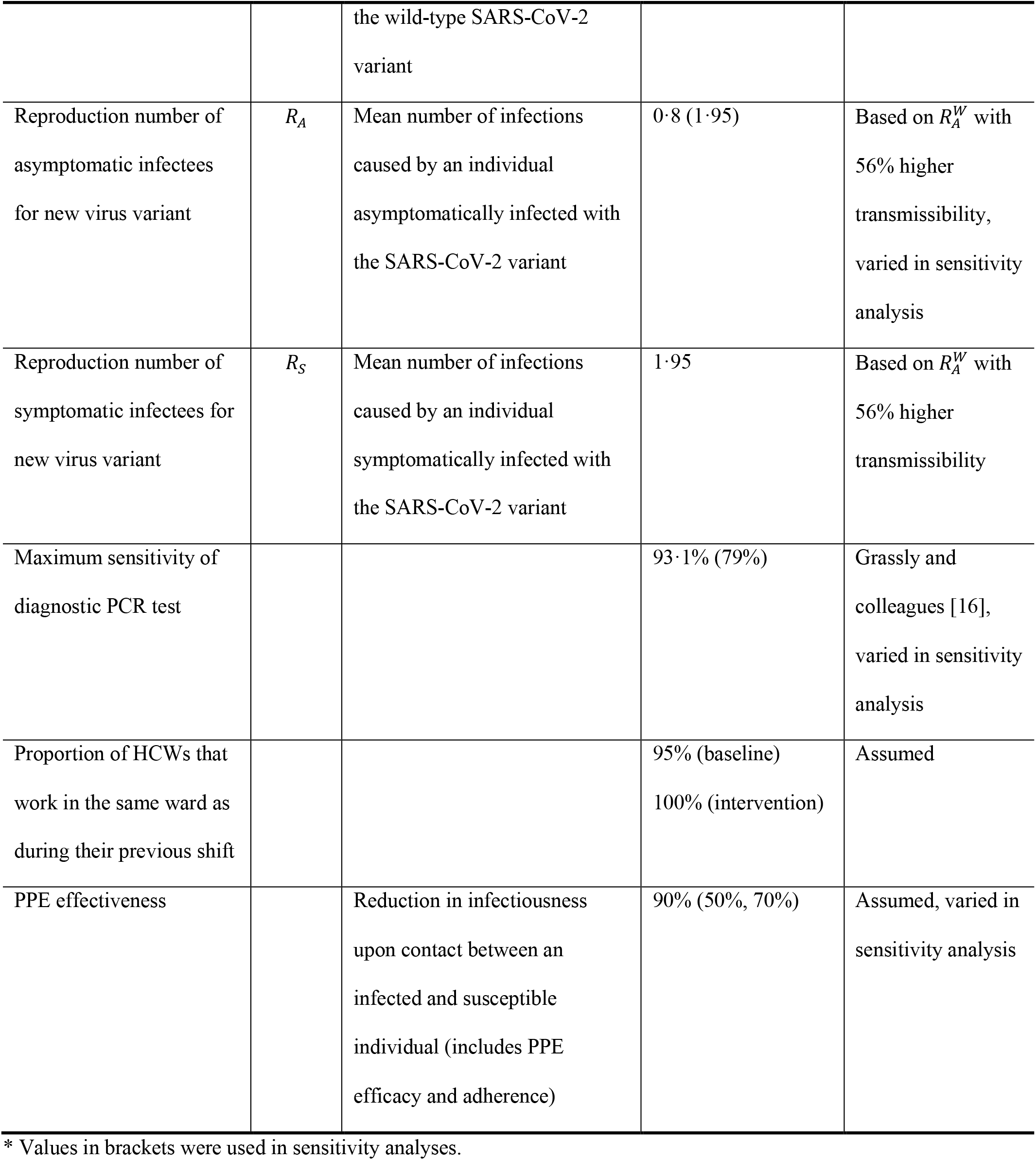
Parameter values for the agent-based model.

First, we chose the reproduction numbers *R*_*S*_ and *R*_*A*_ such that the numbers of occupied beds by COVID-19 patients predicted by our model were in good agreement with real-life UMCU data on the number of COVID-19 patients at UMCU during the first epidemic wave (Table 1 and Fig 2A). These reproduction numbers incorporated the effects of typical (but not COVID-specific) infection prevention measures in the hospital. We will refer to the model parameterized with these reproduction numbers as the *wild-type scenario*. This scenario also assumed that HCWs use 90% effective PPE in COVID wards and isolate at home immediately upon symptom onset for seven days, after which they return recovered to work. Next, we introduced a more transmissible SARS-CoV-2 variant into the hospital, keeping all other parameters – including PPE use in COVID wards and self-isolation after symptom-onset – the same. Based on recent results for B.1.1.7, we assumed a 56% increase in transmissibility [21]. We will refer to the model parametrized with these higher reproduction numbers as our *baseline scenario*. Various intervention scenarios were compared to this baseline scenario.

**Fig 2.**
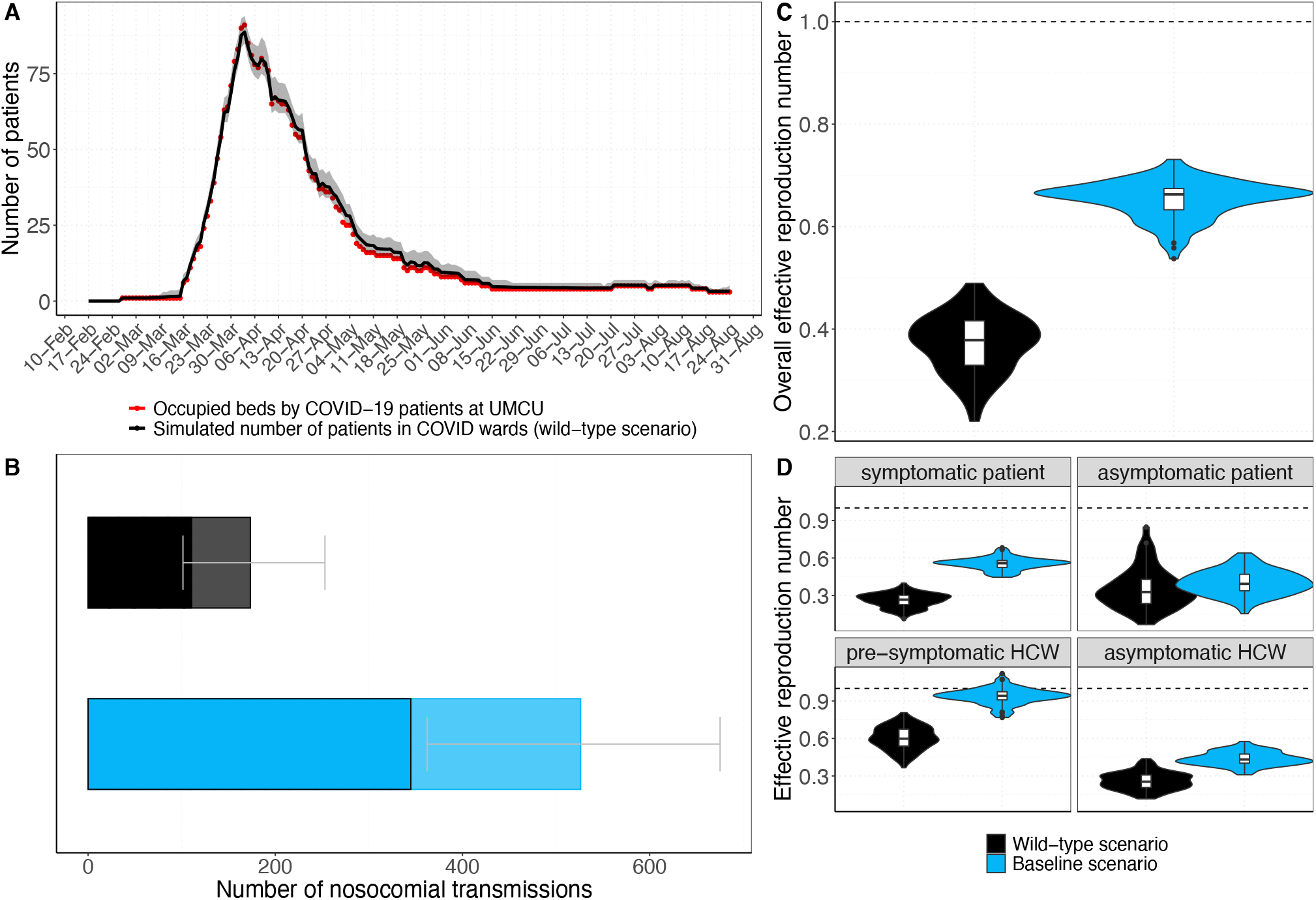
Comparison of the scenarios with the wild-type and a more transmissible SARS-CoV-2 variant. Both scenarios entail 90% effective PPE use in COVID wards. For the wild-type scenario (black), model simulations were performed with *R*_*S*_=1·25 (reproduction number of symptomatically infected individuals) and *R*_*A*_=0·5 (reproduction number of asymptomatically infected individuals). For the baseline scenario (blue), model simulations were performed with *R*_*S*_=1·95 and *R*_*E*_=0·8 (with 56% higher transmissibility with respect to the wild-type SARS-CoV-2 variant). (A) The simulated mean number of beds occupied by patients in COVID wards per day (black curve) and the corresponding 95% uncertainty interval (grey shaded area) over 100 simulation runs is shown. The red points represent the real-life data on the daily number of beds occupied by COVID-19 patients at the UMCU for the time period between 27 February and 24 August 2020. (B) Number of nosocomial transmissions as predicted by the baseline models. The full rectangular bar height represents the mean total Hnumber of nosocomial transmissions during the whole study period (over 100 simulation runs). The grey error bars represent the corresponding 95% uncertainty intervals. Patients that acquire a SARS-CoV-2 nosocomial infection may be diagnosed in the hospital (due to symptom onset during hospital stay or due to detection by an intervention) or discharged to the community in a pre-symptomatic or asymptomatic state. The rectangular bars with the black border represent the mean number of individuals (patients and HCWs) infected with SARS-CoV-2 and diagnosed in the hospital. The lighter rectangular bars represent the remaining mean number of patients discharged to community undiagnosed. (C) Overall effective reproduction numbers for the nosocomial spread in the baseline scenarios. Violin and box plots of the overall effective reproduction numbers (for pre-/symptomatic and asymptomatic patients and HCWs combined) are shown (over 100 simulations). The horizontal dashed line represents a reproduction number of 1. (D) Effective reproduction numbers for the nosocomial spread in the baseline scenarios. Violin and box plots of the effective reproduction numbers for pre-/symptomatic and asymptomatic individuals are shown separately (over 100 simulations). Since HCWs are assumed to immediately self-isolate upon symptom onset, the reproduction number is assigned to the pre-symptomatic state. The horizontal dashed line represents a reproduction number of 1.

### Diagnostic performance of the PCR test

We assumed a PCR test specificity of 100% and distinguished two scenarios for the test sensitivity: 1) a time-invariant perfect sensitivity of 100%; and 2) a sensitivity increasing with time since infection with a maximum sensitivity of 93·1% close to symptom onset and declining afterward (time-varying sensitivity) [16]. We considered two sensitivity analyses to test the impact of PCR test sensitivity assumptions on our results (Appendix pp.3). Hospital staff typically self-quarantine from symptom onset, get tested and receive their test results within hours (based on UMCU data). We, therefore, assumed no delay between testing and receiving test results, and that HCWs do not contribute to virus transmission after symptom onset.

### Infection control interventions

#### Baseline scenario

In the baseline scenario, HCWs were assumed to use PPE in COVID wards when attending to patients, but not during breaks or in other parts of the hospital. The baseline reduction factor (PPE effectiveness) was assumed to be 90%, which includes both perfect-use PPE efficacy and expected PPE use adherence level. We assumed that 95% of the HCWs work in the same ward as during their previous shift.

All interventions described below were in addition to the baseline scenario.

#### Intervention: PPE in all wards

In this scenario, all HCWs used 90% effective PPE in all (non-COVID and COVID) wards. However, no PPE was used when HCWs meet each other off-ward. We performed sensitivity analyses assuming PPE effectiveness of 50% and 70%.

#### Intervention: HCW cohorting (no ward change)

This scenario restricted HCWs to work only in specific wards and did not allow any ward changes.

#### Intervention: Regular HCW screening

All HCWs were tested for SARS-CoV-2 either with a) a test with perfect sensitivity every three days, or a test with time-varying sensitivity, b) every three days, or c) every seven days. If tested positive, HCWs were assumed to immediately self-isolate at home for seven days.

#### Intervention: HCW contact-tracing

If a HCW developed symptomatic SARS-CoV-2 infection, all contacts in the hospital during a time window of either two or seven days before symptom onset were traced and tested. We will refer to these scenarios as *2-day Contact tracing* and *7-day contact tracing*. For *2-day contact tracing*, contacts were always tested assuming a time-varying test sensitivity. For *7-day contact tracing*, we distinguished between perfect and time-varying sensitivity sub-scenarios. In the perfect sensitivity sub-scenario, contacts were instantaneously tested on the day of symptom onset of the index (the HCW). In the time-varying test sensitivity sub-scenario, the test was performed on the day of symptom onset if the contact with the index was more than five days ago. Otherwise, it was performed on day five after the contact. Exposed HCWs awaiting tests were assumed to wear PPE during contact with any patient and with other HCWs. In case of a positive test, patients were moved to a COVID ward while infected HCWs were sent home for self-isolation for seven days and replaced by susceptible HCW. We did not model any absences of HCWs with disease symptoms caused by other respiratory pathogens.

#### Outcome measures

We computed the effective reproduction number *R*_*E*_ (average number of secondary cases caused by an infected individual) to evaluate an intervention’s effectiveness. We calculated an overall *R*_*E*_ for an average individual (patients and HCWs combined) but also stratified *R*_*E*_ by patients, HCWs, and symptom status. The reproduction numbers of patients were calculated for those who eventually developed symptoms 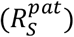 and those who remained without symptoms 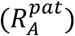. Since HCWs were assumed to immediately self-isolate upon symptom onset, we calculated *R* during pre-symptomatic 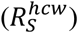 and asymptomatic states 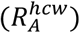. To evaluate the maximum demand on hospital capacity, we considered the total number of nosocomial infections among patients and HCWs over time. In addition, we computed the percentage of absent HCWs due to self-isolation (because of symptom onset or detection via screening or contact-tracing) over time. We assessed the efficiency of screening and contact-tracing interventions by their positivity rates (percentage of detected infected individuals among tested individuals). We did not include individuals that developed symptoms prior to being tested in the positivity rate calculations since those were already detected and isolated in our model. For every scenario, we calculated the mean and 95% percentiles over 100 simulation runs (95% uncertainty interval). We calculated positivity rates over time merging data from all simulation runs and computed 95% Bayesian beta-binomial credibility intervals.

A detailed description of the full model and the parameters can be found in the appendix. We performed sensitivity analyses to test the robustness of our results (Table 1). The data and full code are available from https://github.com/htahir2/covid_intra-hospital_model.git.

## Results

We observed good agreement between the number of patients in COVID wards predicted by our wild-type scenario and the real-life UMCU data during the first wave for *R*_*S*_=1·25 and *R*_*A*_=0·5. However, the model slightly overestimates hospitalizations for the second half of the first wave (Fig 2A). We subsequently assumed the introduction of a SARS-CoV-2 variant with a 56% increase in transmissibility (based on B.1.1.7 data), resulting in *R*_*S*_=1·95 and *R*_*A*_=0·8. Keeping all other parameters the same, including HCWs using PPE in COVID wards and self-isolating at symptom-onset, the total number of nosocomial transmissions increased by 303% (Fig 2B) and the overall effective reproduction number increased by 62·5% (Fig 2C). 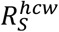 and 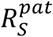 increased the most to 0.94 and 0.6, respectively (Fig 2D), indicating that pre-symptomatic individuals pose the highest risk for onward transmissions.

### Intervention effects on reproduction numbers

In the context of this SARS-CoV-2 variant with higher transmissibility, the baseline scenario of 90% effective PPE use in COVID wards yielded an overall *R*_*E*_ of 0·65 (Fig 3A). Extending PPE use to non-COVID wards reduced *R*_*E*_ by an additional 85%, to 0·1. The effect of HCW screening on *R*_*E*_ highly depended on the test sensitivity. With time-varying test sensitivity, screening every three or seven days reduced *R*_*E*_ to 0·59 and 0·63 (reductions of 9% and 3%), respectively. When perfect sensitivity was assumed, screening every three days reduced *R*_*E*_ by 63%, to 0·24. The impact of contact-tracing also depended on the test sensitivity assumptions, but to a lesser extent. For perfect test sensitivity, 7-day contact-tracing reduced *R*_*E*_ by 32%, to 0·44. For time-varying test sensitivity, the 2-day and 7-day contact-tracing scenarios reduced *R*_*E*_ to 0·41 and 0·39 (reductions of 37% and 40%), respectively. The additional reductions of *R*_*E*_ by the intervention scenarios over and above the baseline scenario were most prominent for pre-symptomatic HCWs (Fig 3B).

**Fig 3.**
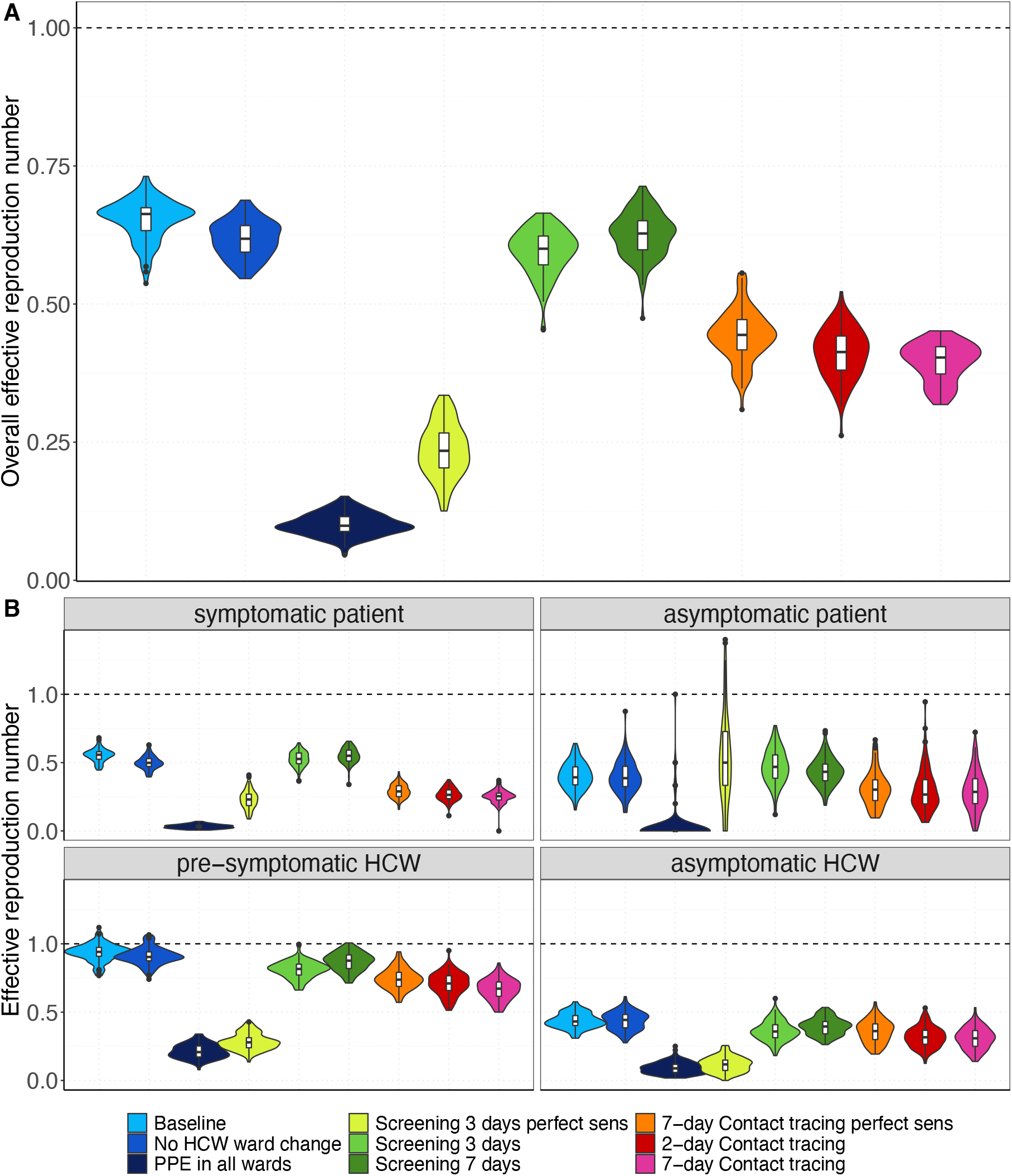
Effective reproduction numbers for the nosocomial spread of the SARS-CoV-2 variant for each simulation scenario. Results shown are based on *R*_*S*_=1·95 and *R*_*A*_=0·8 (reproduction numbers for the SARS-CoV-2 variant with 56% higher transmissibility with respect to the wild-type SARS-CoV-2 variant). (A) For each intervention scenario, violin and boxplots of the overall effective reproduction numbers (for pre-/symptomatic and asymptomatic patients and HCWs combined) are shown (over 100 simulations). (B) For each intervention scenario, violin and boxplots of the effective reproduction numbers for pre-/symptomatic and asymptomatic individuals are shown (over 100 simulations). Since HCWs are assumed to immediately self-isolate upon symptom onset, the reproduction number is assigned to the pre-symptomatic state. The horizontal dashed line represents a reproduction number of 1. For screening every 3 days and 7-day contact tracing prior to symptom onset of SARS-CoV-2 infected HCWs, we considered two different test sensitivity scenarios: time-invariant perfect test sensitivity (perfect sens) and time-varying test sensitivity.

### Intervention effects on numbers of nosocomial infections

PPE use in all wards or HCW screening every three days with perfect test sensitivity would prevent 93·7% and 82·7% of all transmissions, respectively (Fig 4), and both interventions would also prevent outbreaks among patients and HCWs (Fig 5). Reductions in nosocomial infections were much smaller for regular screening interventions with time-varying test sensitivity: screening every three days would lead to a 20·4% reduction and screening once a week to a 10·1% reduction. Testing with perfect test sensitivity followed by 7-day contact-tracing was more effective (55·8% reduction of transmissions) than regular screening every three or seven days. Testing with time-varying sensitivity followed by 2-day or 7-day contact tracing were similarly effective as testing with perfect sensitivity followed by 7-day contact tracing (reductions of 61·4% and 64·1%, respectively). HCW cohorting would decrease the total number of nosocomial infections by 13%. Note that our model predicted that 62%-78% of all nosocomial infections are diagnosed in the hospital either due to testing after symptom onset or testing as part of an intervention (Appendix Fig 2). The remaining 22%-38% of nosocomial infections are undiagnosed infections in patients without symptoms (yet) at the time of discharge.

**Fig 4.**
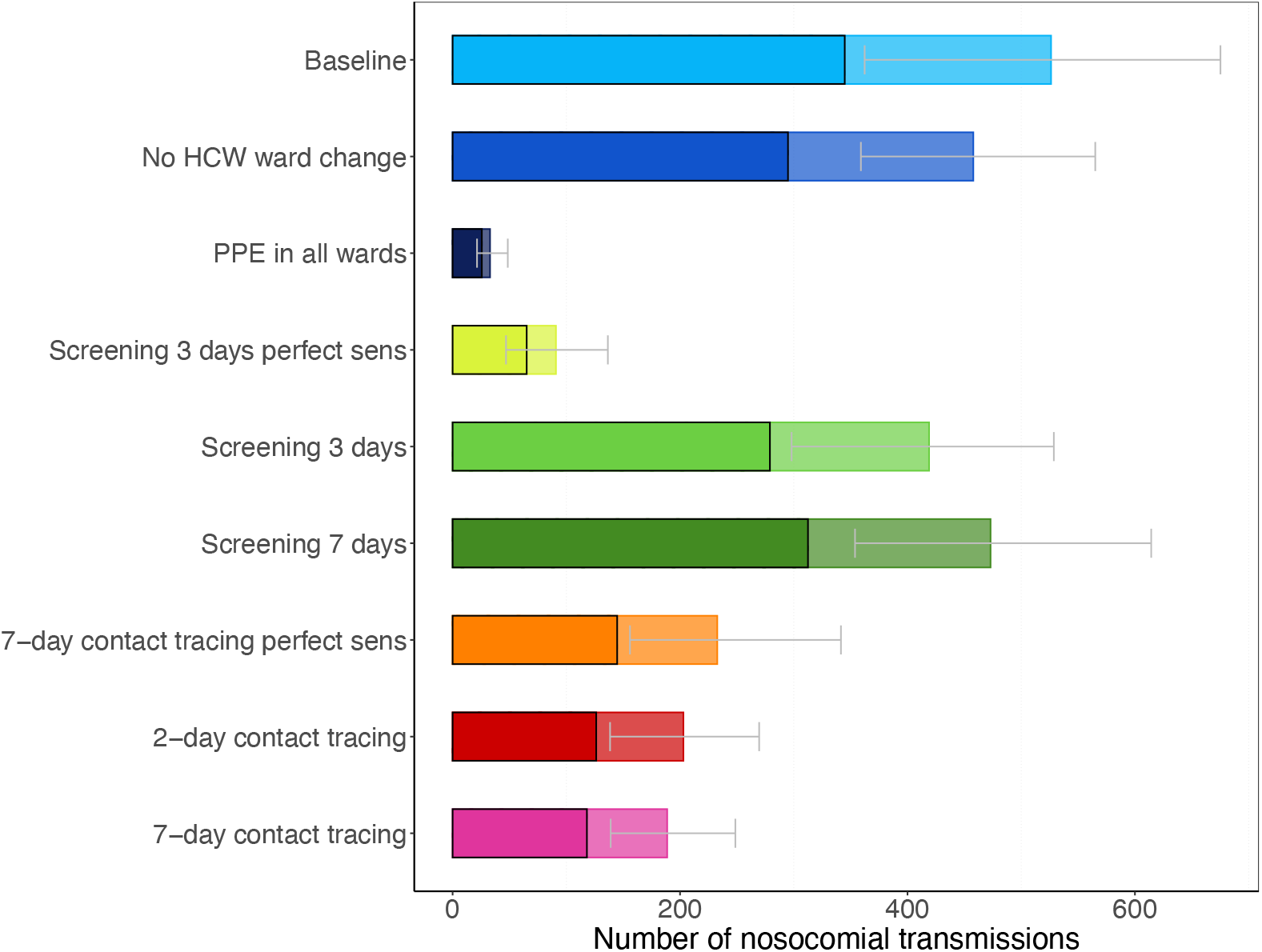
Number of nosocomial transmissions of the SARS-CoV-2 variant for each simulation scenario. Results shown are based on *R*_*S*_=1·95 and *R*_*A*_=0·8 (reproduction numbers for the SARS-CoV-2 variant with 56% higher transmissibility with respect to the wild-type SARS-CoV-2 variant). The full rectangular bar height represents the mean total number of nosocomial transmissions during the whole study period (over 100 simulation runs). The grey error bars represent the corresponding 95% uncertainty intervals. Patients that acquire a SARS-CoV-2 nosocomial infection may be diagnosed in the hospital (due to symptom onset during hospital stay or due to detection by an intervention) or discharged to the community in a pre-symptomatic or asymptomatic state. The rectangular bars with the black border represent the mean number of individuals (patients and HCWs) infected with SARS-CoV-2 and diagnosed in the hospital. The lighter rectangular bars represent the remaining mean number of patients discharged to community undiagnosed. For screening every 3 days and 7-day contact tracing, we considered two different test sensitivity scenarios: time-invariant perfect test sensitivity (perfect sens) and time-varying imperfect test sensitivity.

**Fig 5.**
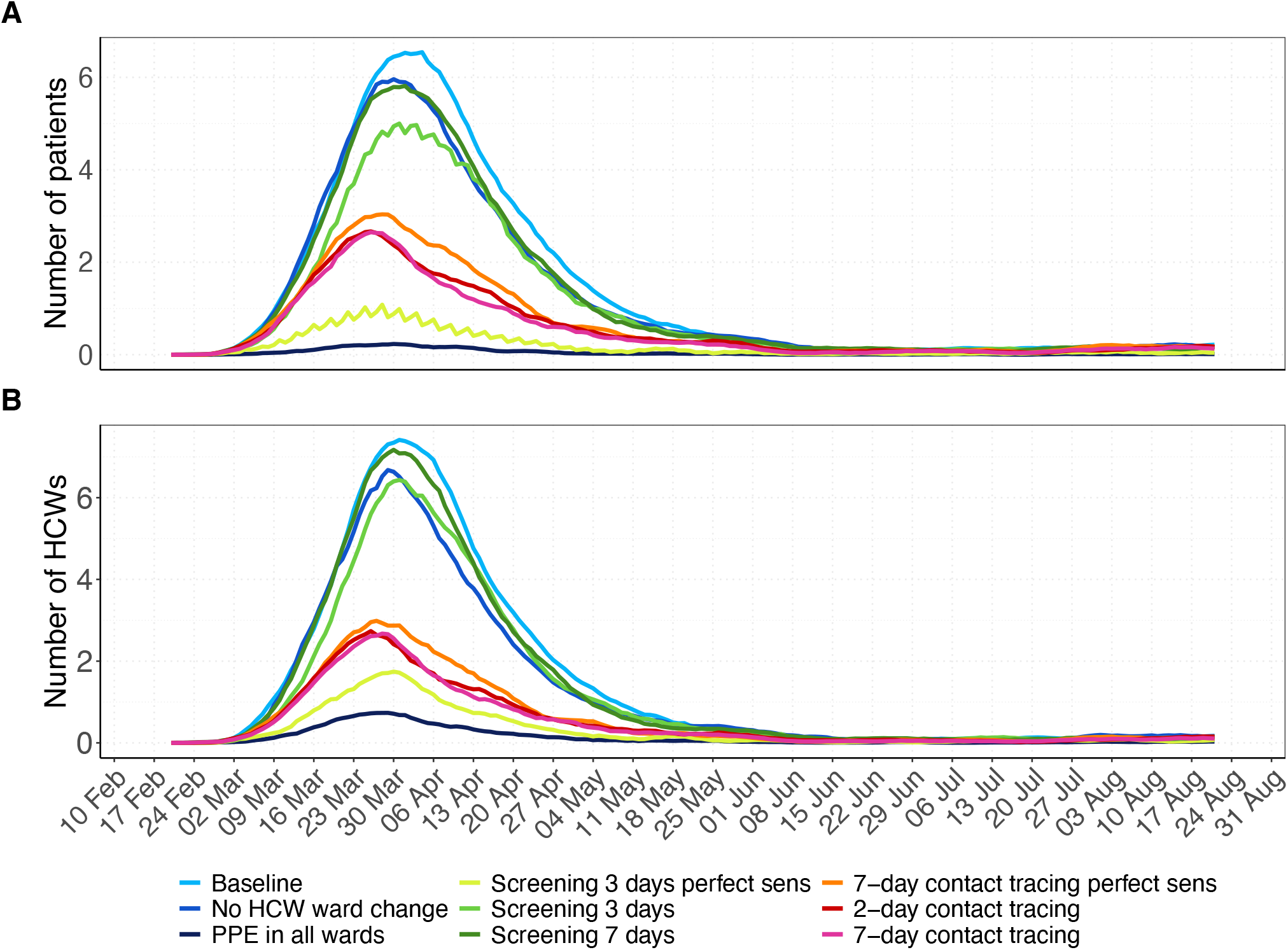
Number of nosocomial infections among patients and HCWs over time for all simulation scenarios with the SARS-CoV-2 variant. Results shown are based on *R*_*S*_=1·95 and *R*_*A*_=0·8 (reproduction numbers for the SARS-CoV-2 variant with 56% higher transmissibility with respect to the wild-type SARS-CoV-2 variant). For each scenario, the 7-day moving average of the mean prevalence (over 100 simulation runs) is shown. A) Number of hospital-acquired infections among patients. B) Number of hospital-acquired infections among HCWs. For screening every 3 days and contact tracing 7 days prior to symptom onset of SARS-CoV-2 infected HCWs, we considered two different test sensitivity scenarios: time-invariant perfect test sensitivity (perfect sens) and time-varying imperfect test sensitivity.

### Intervention effects on HCW absenteeism

Our baseline scenario predicted a maximum HCW absenteeism of 5·4%, including absenteeism due to symptoms or home isolation (Fig 6). When comparing intervention scenarios to the baseline scenario, HCW absenteeism is lowest for PPE use in all wards (a maximum of 2·3%). The maximum absenteeism percentages were 5·2% for HCW cohorting, 5·1% for regular screening with perfect test sensitivity, 8·6% for regular screening with time-varying test sensitivity every seven days and 6·6% every three days, 4·0% for 7-day contact tracing with testing assuming perfect sensitivity, 3·6% for 2-day contact tracing with testing assuming time-varying sensitivity, and 3·9% for 7-day contact tracing with testing assuming time-varying sensitivity.

**Fig 6.**
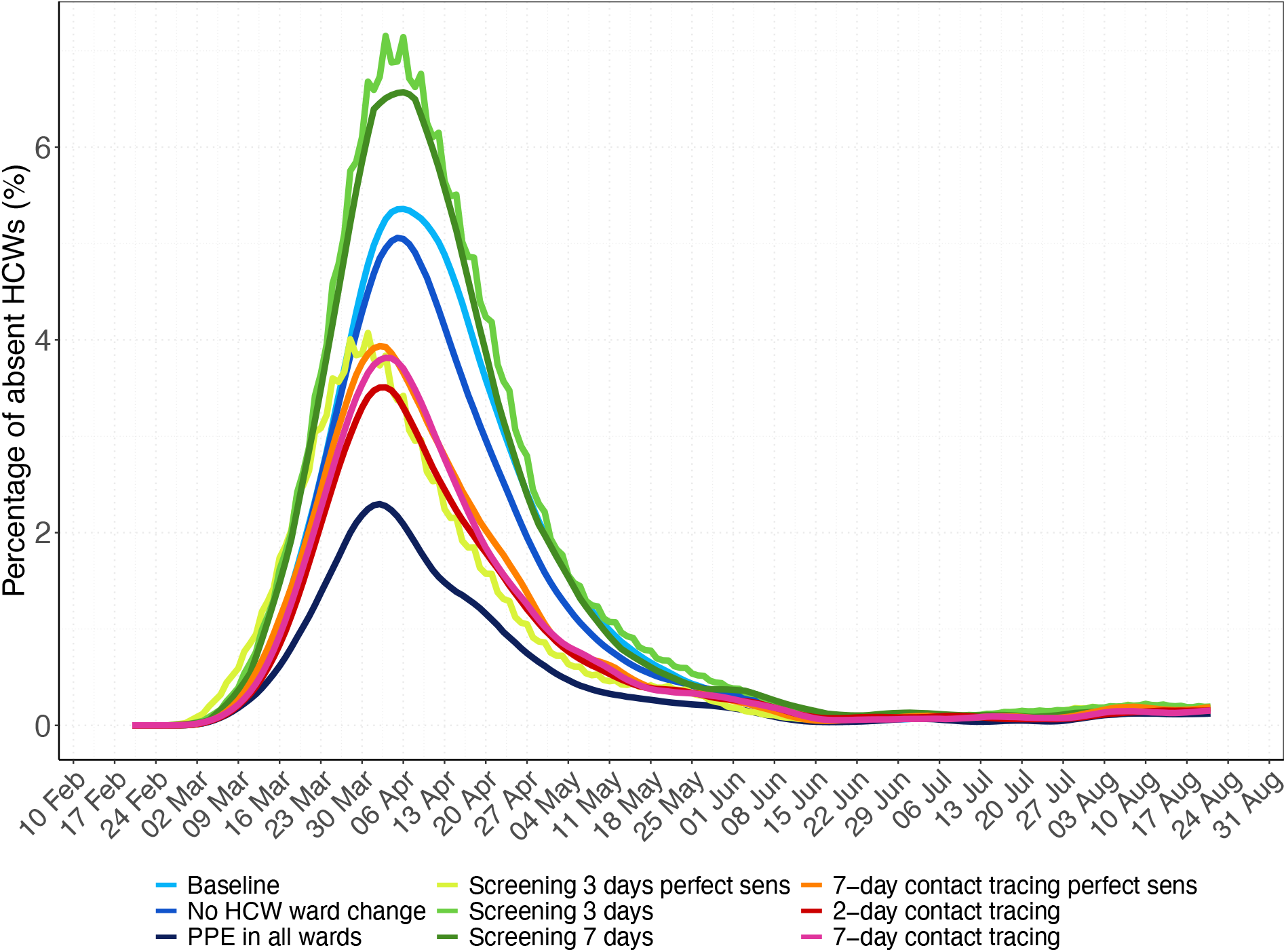
Daily percentage of absent HCWs during the hospital epidemic for each simulation scenario. Results shown are based on *R*_*S*_=1·95 and *R*_*A*_=0·8 (reproduction numbers for the SARS-CoV-2 variant with 56% higher transmissibility with respect to the wild-type SARS-CoV-2 variant). The 7-day moving average of the mean percentage (over 100 simulation runs) of HCWs absent from work due to symptom onset or a detected SARS-CoV-2 infection screening or contact tracing is shown. For screening every 3 days and contact tracing 7 days prior to symptom onset of SARS-CoV-2 infected HCWs, we considered two different test sensitivity scenarios: time-invariant perfect test sensitivity (perfect sens) and time-varying imperfect test sensitivity.

### Efficiency of screening and contact-tracing interventions

HCW screening every three days with a perfect test would lead to the lowest test positivity rate of all testing-based interventions (Fig 7A). Screening of HCWs every week compared to every three days yields higher positivity rates with its mean reaching a maximum value of 5·1%. The positivity rate of screening interventions linearly increases with increasing prevalence (Appendix Fig 8).

**Fig 7.**
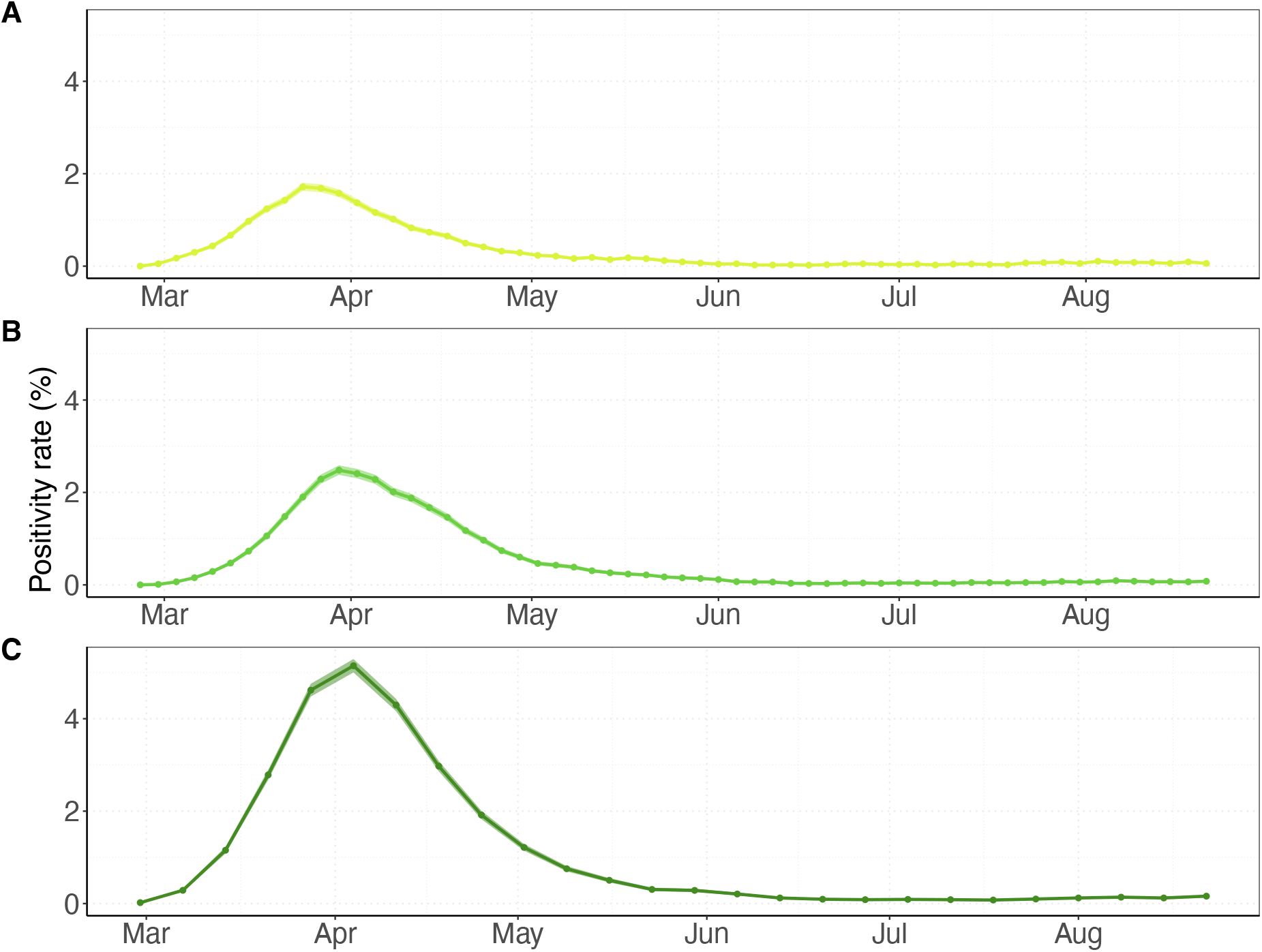
Positivity rates over time for screening interventions. Results shown are based on *R*_*S*_=1·95 and *R*_*A*_=0·8 (reproduction numbers for the SARS-CoV-2 variant with 56% higher transmissibility with respect to the wild-type SARS-CoV-2 variant). Positivity rates were calculated by the number of positive detected HCWs among the number of tested HCWs using data of all simulation runs combined (points). The shaded regions represent the 95% Bayesian beta-binomial credibility intervals. HCWs who developed symptoms prior to the day of testing were not included in the positivity rate as we assume that they were already correctly identified. (A) Screening every three days with time-invariant perfect test sensitivity. (B) Screening every three days with time-varying imperfect test sensitivity. (C) Screening every seven days with time-varying test sensitivity.

**Fig 8.**
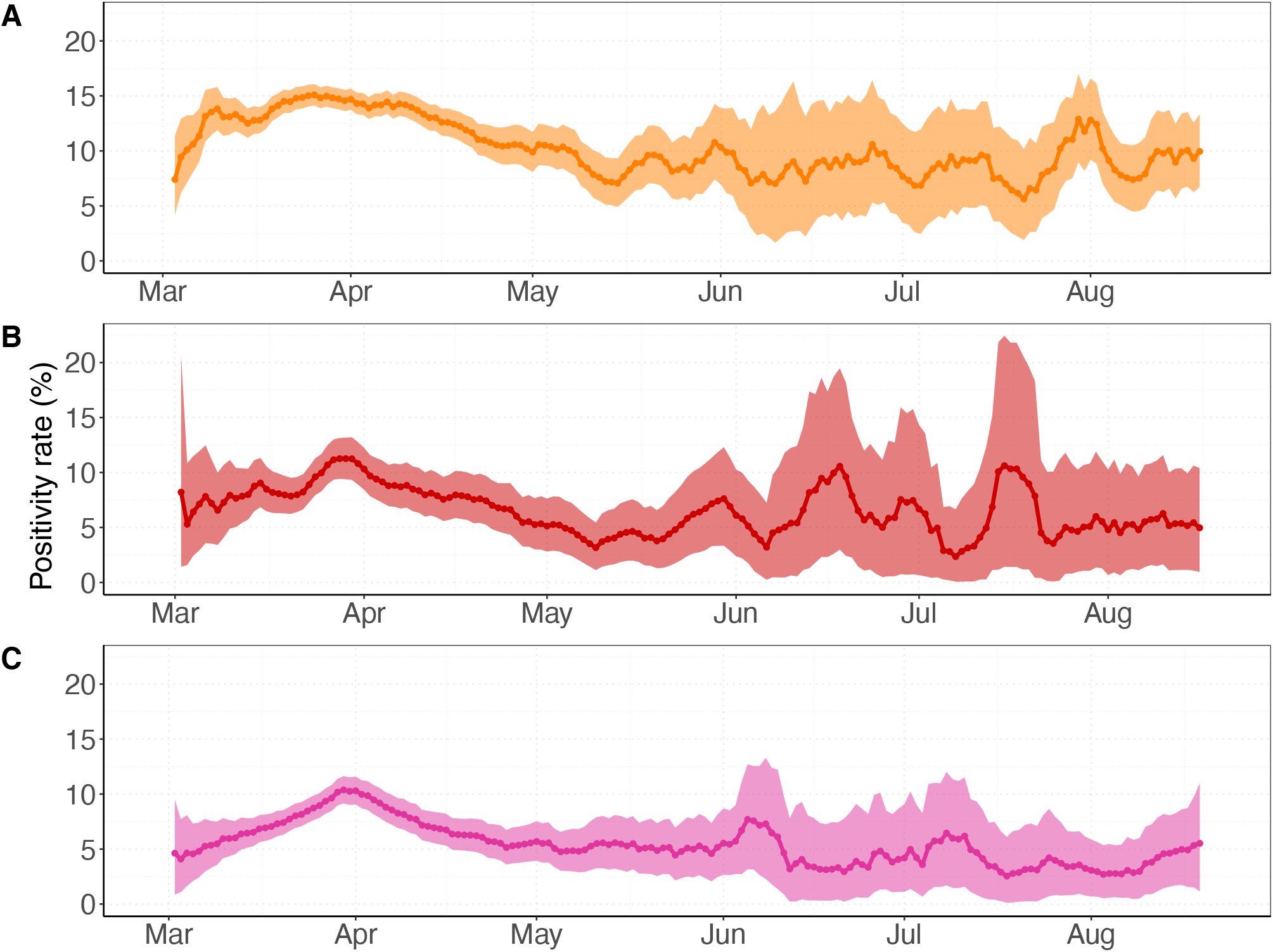
Positivity rates over time for contact tracing interventions. Results shown are based on *R*_*S*_=1·95 and *R*_*A*_=0·8 (reproduction numbers for the SARS-CoV-2 variant with 56% higher transmissibility with respect to the wild-type SARS-CoV-2 variant). The positivity rate is computed by the percentage of positive tested contacts among all traced contacts using data of all 100 simulation runs merged. Positivity rates are assigned to the day of symptom onset of the index case, i.e., HCW that developed symptoms due to a SARS-CoV-2 infection. Traced contacts who developed symptoms due to a SARS-CoV-2 infection are excluded from contact tracing as we assume that they are always correctly identified. The plot shows the 7-day moving average (colored line) and the 95% Bayesian beta-binomial confidence interval (shaded area). (A) Tracing contacts of symptomatically infected HCWs of the last two days before symptom onset using a diagnostic test with perfect test sensitivity. (B) Tracing contacts of symptomatically infected HCWs of the last two days before symptom onset with testing five days after contact with the index case assuming time-varying test sensitivity. (C) Tracing contacts of symptomatically infected HCWs of the last seven days before symptom onset with testing five days after contact with the index case assuming time-varying test sensitivity.

Positivity rates for contact-tracing interventions are much higher than for screening interventions, reaching as high as 15·1% when a perfect test sensitivity is assumed (Fig 8A). The maximum positivity rates for 2-day and 7-day contact tracing with time-varying test sensitivities are only slightly lower at 11·3% and 10·4%, respectively (Fig 8B-C). Positivity rates of contact-tracing interventions are stable across prevalence values (Appendix Fig 9).

Sensitivity analyses show that our findings do not change significantly when the assumed PPE effectiveness is reduced to 70%. When PPE effectiveness is assumed to be as low as 50%, screening every three days with perfect sensitivity becomes more effective than PPE use in all wards. However, PPE use in all wards is still more effective than all other interventions (Appendix pp. 6).

## Discussion

During the first epidemic wave of the wild-type SARS-CoV-2 in the Netherlands, nosocomial transmission was considered to be of relative minor importance. Our results suggest that a more transmissible virus variant could significantly increase the total number of nosocomial transmissions if hospital prevention measures would not be expanded beyond those implemented during the first wave (HCWs using PPE with assumed 90% effectiveness in COVID-19 wards and self-isolating at home after symptom onset). Our findings suggest that universal PPE use in all hospital wards is the most effective intervention to reduce the reproduction number and absenteeism. These results are consistent with a previous modelling study and previous findings on significant reductions of nosocomial-acquired SARS-CoV-2 infections after implementation of universal masking policies [1,11,13,23–26].

In our model, HCW cohorting only had a small impact on nosocomial transmissions, which is due to the fact that we assumed 90% effective PPE use in the COVID wards in all scenarios. Several studies have reported elevated risks for HCWs working in COVID-19 patient care [5,6]. Our results suggest that maintaining sufficient PPE supplies in hospital settings may reduce the need for implementing additional HCW cohorting strategies.

Our model also suggested that regular screening of HCWs could have a strong impact, but only if the test sensitivity is high throughout the infectious period. Tests with imperfect time-varying sensitivity miss many infections during the pre-symptomatic phase. Indeed, our model identified pre-symptomatically infected HCWs as drivers of transmission both to patients and to other staff. This is consistent with a descriptive study on HCWs in France where contacts causing the transmissions took place in the pre-symptomatic phase of the index case in 30% of all cases and in almost 50% of HCW-HCW transmissions. Our results also agree with previous modelling studies suggesting that regular screening of HCWs was less effective than effective PPE use.

Contact tracing was highly effective in limiting nosocomial transmissions in our model, especially when traced contacts are tested at least five days after their exposure and precautionary measures are undertaken in the meantime. If traced HCWs are immediately tested, self-isolated, and replaced by susceptible HCWs, this can lead to increased transmission, a phenomenon that was also observed by Scarpino and colleagues [27]. The authors used a network model and evidence from data on influenza and dengue outbreaks to show that replacing infected individuals in essential societal roles with susceptibles may lead to accelerated transmission. Our results indicate that allowing traced HCWs to work with PPE in all hospital wards is more effective in limiting transmission. Finally, our model suggests that contact tracing yields higher positivity rates than screening interventions, not only at high prevalence but also during periods of low infection rates, making this also a potentially successful and cost-effective infection control strategy in hospital settings. Our findings reinforce the recommendation by Paltansing and colleagues to test all close contacts of a SARS-CoV-2 positive case immediately and subsequently on day 3 and 7 regardless of symptoms and to allow HCWs to work with surgical masks while awaiting their test results [13].

Our study has several limitations. First, we assumed that transmission occurs solely via HCWs in the absence of a direct patient-to-patient contact pathway, as has been used before in an individual-based model of nosocomial influenza transmission [28]. Assuming similar transmission modes for SARS-COV-2, we consider this assumption reasonable for hospital settings in Western countries where direct patient-to-patient contact is rare. When this assumption is violated, our estimated impact of HCW-based interventions is likely to be overestimated. Second, we considered SARS-CoV-2 as a cause of symptoms and neglected other respiratory tract infections. Thus, real-life positivity rates of contact tracing may be lower than presented in this study. Finally, duration of contacts, SARS-CoV-2 reinfections, visitors or other ancillary staff, delays between symptom onset and isolation, or delays between test application and test result were not included. We have not used formal fitting procedures to match our model results to the data given the large number of parameters. However, qualitatively, our conclusions were robust in sensitivity analyses to variation of the most important model parameters. While our model was developed using data of a large Dutch teaching hospital and of the first wave of the COVID-19 epidemic in the Netherlands, our results can be generalised to other hospitals with a similar structure and may be relevant for subsequent waves and future infectious disease outbreaks.

## Conclusions

In conclusion, our model demonstrates that PPE use in all wards is the most effective measure to substantially reduce nosocomial spread of SARS-CoV-2 variants with higher transmissibility. However, contact-tracing and regular screening using high-sensitivity tests are also effective interventions, which might be preferred in some settings.

## Data Availability

All data and code used in the manuscript are available from:
https://github.com/htahir2/covid_intra-hospital_model

https://data.rivm.nl/covid-19/COVID-19_prevalentie.json

https://opendata.cbs.nl/statline/#/CBS/nl/dataset/71857ned/table?ts=1517582466533

http://doi.org/10.5281/zenodo.3711575

http://doi.org/10.5281/zenodo.3711575

https://github.com/htahir2/covid_intra-hospital_model/blob/main/data/covid_patient_admissions_los_UMCU.csv

## Availability of data and materials

The datasets used and/or analysed as well as the full code reproducing the results in the current study are available from https://github.com/htahir2/covid_intra-hospital_model.git.

### Abbreviations

COVID-19: Coronavirus disease 2019
HCW: Healthcare worker
ICU: Intensive-care unit
LTCF: Long-term care facilities
PPE: Personal protective equipment
RT-PCR: Reverse transcriptase polymerase chain reaction
R_E_: Effective reproduction number
RIVM: Rijksinstituut voor Volksgezondheid en Milieu (Dutch National Institute for Public Health and the Environment)
SARS-CoV-2: Severe acute respiratory syndrome coronavirus 2
SEIR: Susceptible-Exposed-Infectious-Recovered
UMCU: University Medical Center Utrecht

## Acknowledgements

We thank Jantien Backer (National Institute for Public Health and Environment of the Netherlands, RIVM) for helpful explanations on the data provided by the RIVM.

## Author contributions

TMP and HT have contributed equally to this work. TMP, HT, MK, MCJB, and JHHMvdW developed the conceptual framework of the study. TMP, HT, MK and MCJB developed the model. HT programmed the model and produced the output. HT and TMP produced the results of the model. TMP produced the visualization for the main text and the appendix. TMP, MK, BvdR and JHHMvdW conducted the literature research. PE and BvdR collected the data. TMP and HT have verified the underlying data. MK, MCJB, MB, JHHMvdW and PE contributed to interpretation of the results. TMP wrote the original draft of the main text. TMP and HT wrote the appendix. All authors provided critical review of the manuscript, and approved its final version for submission.

