## Supplementary material for "Interventions to control nosocomial transmission of SARS-CoV-2: a modelling study"

**Includes:**
Appendix Methods
Appendix Table
Appendix Figures

[#] **These authors contributed equally to this work.**
[*] **Corresponding author:**
Thi Mui Pham
Address: Julius Center for Health Sciences and Primary Care
University Medical Center Utrecht
P.O. Box 85500 Utrecht
The Netherlands


### Appendix Methods

We simulated nosocomial COVID-19 epidemics using an agent-based model coded in Python. The code is available from: https://github.com/htahir2/covid_intra-hospital_model.git. First, we fitted the model to real-life data from the University Medical Center Utrecht (UMCU) during the period February-August 2020. Next, we evaluated the impact of various intervention strategies aimed at healthcare workers (HCWs) on the nosocomial spread of a more transmissible SARS-CoV-2 variant (e.g., B.1.1.7) in a hospital.

We first outline the data used to inform and parametrize our model. We continue by explaining the details of our agent-based model, the transmission model, and the underlying assumptions. We further describe the intervention strategies implemented in our model, the considered outcome measures, and the results of our sensitivity analyses. Lastly, we elaborate on the algorithm of our model.

### Data

We used data from the University Medical Center Utrecht (UMCU), The Netherlands, and data provided by the National Institute for Public Health and the Environment (RIVM), The Netherlands, during the first wave of the SARS-CoV2 epidemic to inform and parametrize our model.

*Hospitalization data*
We used hospitalization data of patients in COVID wards at the UMCU between 27 February and 24 August 2020. The data set comprises 167 admissions of which 82 patients were admitted to an intensive care unit, and 85 patients were admitted to regular wards. Based on information of the infection control department of the UMCU, we assumed that 95% of those admissions were COVID-19 admissions, leaving 5% of the patients admitted for non-COVID reasons but later diagnosed with a SARS-CoV-2 infection in the hospital. The number of admissions per day are shown in Appendix Figure 10. The data further comprises discharge dates of the respective patients. We used the resulting number of beds occupied by COVID-19 patients to fit the reproduction numbers in our model. The data is available from: https://github.com/htahir2/covid_intra-hospital_model/blob/main/data/covid_patient_admissions_los_UMCU.csv

*Length of stay distributions*
We calculated the respective length of stay (LoS) from admission and discharge times for each of the COVID-19 patients at UMCU. Data on patient admissions to the UMCU prior to the COVID-19 pandemic (2014-2017) were used to estimate the length of stay distributions for non-COVID admissions to the hospital. We fitted probability distributions to the length of stay data for admissions to regular wards and ICUs, separately for COVID or for non-COVID related admissions. We considered exponential, log-normal, gamma, and Weibull distribution and chose the best fit by visual inspection of the empirical vs theoretical densities, the Q-Q plot, and the P-P plot. The length of stay data and fitted distributions are shown in Appendix Figure 11. The respective parameters can be found in Appendix Table 1.

*Importation from community*
We assume that 40 patients are admitted to the hospital for reasons unrelated to COVID-19 per day in the time period 27 February to 24 August 2020 (Appendix Table 1). We based this number on UMCU admission data in the time period 2014-2017 and the assumption that admissions decreased by 50% during the first wave of the COVID-19 epidemic. Those admitted patients might be infected with SARS-CoV-2 due to transmissions in the community. We further assume that HCWs go home after each daily shift and therefore may acquire infection in the community as well. They may be in their pre-symptomatic phase or asymptomatically infected with SARS-CoV-2 when they arrive at work in the hospital. These patients and HCWs do not experience any symptoms (yet), and therefore do not know that they are infected. We approximate the probability of being infected in the community for non-COVID patient admissions and HCWs arriving at work as follows:
We used data on the number of infectious people in the Netherlands estimated by the National Institute for Public Health and the Environment (RIVM) from 17 February till 24 August 2020.[1] They used data from serological surveys in the Netherlands and related these to numbers of hospitalized cases (stratified by age group), leading to the number of "actual" infections per hospitalized case. They thereby included all infections that led to an immunological response, not only those that were detected in real time by PCR testing. Note that this method might be less reliable when the number of hospitalizations is low. This estimated number of infectious individuals includes hospitalized COVID-19 patients as well as individuals that are isolated at home (e.g., due to detection in the community via testing or contact tracing). To roughly account for this, we subtracted the total number of reported cases in the province of Utrecht from the estimated number of infectious individuals (RIVM estimate described before). We hereby assume that all individuals in Utrecht are eligible for admission at the UMCU. We additionally used publicly available age-specific hospitalization rates of the Netherlands of 2012 and age-specific COVID-19 incidence rates in the Netherlands to scale the daily probability of being infected in the community for non-COVID patient admissions and HCWs arriving at work.[2–4] For HCWs, we only used age-specific

COVID-19 incidence rates for age-groups between 20 and 65 years. Since age-specific prevalence values are not available to date, the previous calculation is based on the assumption that the distribution of age groups is roughly the same for incidence and prevalence. Furthermore, we assumed a catchment population size of 100,000 people for the hospital.

**Model**

*Environment*

We modelled a typical (Dutch) hospital comprising 28 wards which are divided as follows

- COVID ICU (4) with 17 beds each
- Normal (non-COVID) ICU (1) with 12 beds
- COVID ward (4) with 3 x 23 beds and one with 22 beds
- Normal (non-COVID) ward (19) with 2 x 20, 4 x 19, 13 x 18 beds

The total number of beds in the hospital is 521. The numbers are approximated in accordance to the number of beds and ward distribution of the UMCU (for patients who stayed at least one day at UMCU).

*Agent-types*

There are three different agents involved in the transmission process within the hospital of our model: Patients (non-COVID and COVID) and health-care workers (HCWs), separated into nurses, and doctors. Visitors or ancillary workers are excluded from the model.

Patients are assumed to occupy a hospital bed in a single room. This assumption is suitable for a setting where the transmission is mainly driven by HCWs as vectors and with no direct patient-to-patient transmission. HCWs have a number of duty shifts per day. We assume that they meet patients in a number of rounds per shift (see Appendix Table 1) and that HCWs meet other HCWs in the common staff room of each ward.

The ratio between HCWs and patients and the time HCWs spend with a patient are ward specific and assumed. The number of HCW duty shifts per day, and the number of rounds per shift are independent from the ward and assumed. The respective parameters can be found in Appendix Table 1.

*Disease progression*

The disease progression of an infection with SARS-CoV-2 was modelled using a Susceptible-Exposed-Infectious-Recovered (SEIR) model and is shown in Figure 1c of the main text. Individuals who have not been infected with SARS-CoV-2 are susceptible (S), and may transition to being exposed (E) upon contact with an infected individual. A proportion $(1 - P_A)$ of infected individuals develop symptoms. We based the incubation period (time between infection and appearance of symptoms) on a Gamma distribution with mean 5.5 days as described by Lauer and colleagues.[5] Symptomatically infected individuals may develop moderate symptoms ($I_M$) or severe symptoms ($I_S$). All infected individuals will eventually recover and become immune ($I_R$). Asymptomatically infected ($I_A$) are assumed to recover after 14 days. We assume that moderately and severely infected patients recover after 14 and 35 days, respectively.[6] We assumed that symptomatically infected HCWs are perfectly isolated at home for seven days immediately upon developing symptoms, after which they return recovered to work. Based on a meta-analysis by Buitraga-Garcia and colleagues, we assumed the asymptomatic proportion of COVID-19 infections among patients to be 20% and the proportion of asymptomatic infections among HCWs to be 31% (see also Appendix Table 1).[7] We used their overall estimate of the proportion of asymptomatic infected individuals for the patient population in our model, and their estimates obtained from studies with screened individuals for the HCW population in our model.

*Hospital admissions*

Patients can be hospitalized either for non-COVID reasons to normal wards and ICUs, or with moderate or severe COVID-19 symptoms to COVID wards or COVID-ICUs. The length of stay of a patient differs according to these four categories. Probability distributions were fitted to length of stay data of patients admitted to the UMCU. The data and fitted distributions are shown in Appendix Figure 10. The respective parameters can be found in Appendix Table 1. Patients admitted to normal wards and ICUs who are later detected with a SARS-CoV-2 infection are immediately transferred to COVID wards and ICUs upon diagnosis. If they develop severe symptoms, their length of stay is prolonged according to the length of stay distribution of admitted severe COVID-19 patients.

*Accuracy of the diagnostic test*

In our model, patients and HCWs are assumed to be tested using reverse transcriptase polymerase chain reaction (RT-PCR) either when being screened or after being identified as a contact of a symptomatic infected individual in contact tracing. These diagnostic tests can be inaccurate either because of a false positive or a false negative result. The latter are considered to be more consequential with a potential high impact on onward transmission due to undetected cases. It has been documented that the sensitivity of the PCR test varies with time from exposure and symptom onset.[8] We assumed a time-varying imperfect sensitivity of the diagnostic test (Appendix Figure 1) based on the results reported in Grassly and colleagues.[9] These authors used published data from three meta-analyses of the test sensitivity over time since symptom onset. They assume the pre-symptomatic sensitivity to be proportional to the infectiousness curve such that the estimate on day 5 matched the empirical data from the day of symptom onset. We performed a sensitivity analysis assuming 1) a 15% lower sensitivity (see Appendix Figure 1) and 2) a test sensitivity that stays at its peak value after reaching the maximum. Finally, we assessed the impact of perfect sensitivity of 100% which stays constant over time on the results of our model. Throughout the simulations, we assume test sensitivity to be the same for symptomatic and asymptomatic infections, and we assume a specificity of 100%.

*Infectiousness*

We used a time-varying infectiousness profile following Grassly and colleagues and shown in Figure 1c of the main text.[9] Infectiousness is assumed to vary over time since infection and to follow a Weibull distribution, with a mean of 6 days.[9] The average duration of the infectious period is therefore assumed to be 6 days. This approximation is consistent with published estimates of the serial interval for SARS-CoV-2.[10–12] We denote infectiousness over time since infection $\tau$ by $\beta(\tau)$. It is the mean rate at which an individual infects others at time $\tau$ after being itself. We use the infectiousness profile for calculating the probability of transmission from an infectious to a susceptible individual (see below). The reproduction number $R$ (average number of secondary cases caused by an infected individual) is given by integrating $\beta(\tau)$ over time since infection $R = \int_0^\infty \beta(\tau) d\tau$. The generation time distribution $\omega(\tau)$ is given by unit normalisation such that $\omega(\tau) = \beta(\tau)/R$. Assuming the mean generation time to be equivalent with the observed mean serial interval, we calculate the infectiousness profile by $\beta(\tau) = \omega(\tau)R$. We assumed the infectiousness over time since infection to differ between asymptomatic and symptomatic infected individuals, defined by $\beta_A(\tau)$ and $\beta_S(\tau)$, respectively. Integrating these two functions over time gives the respective reproduction numbers $R_A$ and $R_S$ (Appendix Table 1).
First, we chose the reproduction numbers $R_S$ and $R_A$ such that the numbers of occupied beds by COVID-19 patients predicted by our model were in good agreement with real-life UMCU data on the number of COVID-19 patients at UMCU during the first epidemic wave (**Error! Reference source not found.** and **Error! Reference source not found.**A). These re production numbers incorporated the effects of typical (but not COVID-specific) infection prevention measures in the hospital. We will refer to the model parameterized with these reproduction numbers as the *wild-type scenario*. This scenario also assumed that HCWs use 90% effective PPE in COVID wards and isolate at home immediately upon symptom onset for seven days, after which they return recovered to work. Next, we introduced a more transmissible SARS-CoV-2 variant into the hospital, keeping all other parameters – including PPE use in COVID wards and self-isolation after symptom-onset – the same. Based on recent results for B.1.1.7, we assumed a 56% increase in transmissibility.[13] We will refer to the model parametrized with these higher reproduction numbers as our *baseline scenario*. Various intervention scenarios were compared to this baseline scenario.

*Transmission*

Transmission events can occur between susceptible patients and HCWs, or between (asymptomatic or pre-symptomatic) HCWs. Thus, we assumed that there is no direct transmission between patients. Only HCWs in their pre-symptomatic stage, or HCWs who are asymptomatically infected, contribute to transmission, since we assume that HCWs are perfectly isolated at home for seven days immediately upon developing symptoms. Upon a contact between two individuals, a transmission may take place between an infected and a susceptible individual. The probability of transmission is dependent on the current infectiousness of the infected individual. If $\beta(\tau)$ is the infectiousness of the infected individual at time $\tau$ since infection, the average probability of transmission per contact with a susceptible person is given by

$$A(\tau) = \frac{\beta(\tau)}{c}$$

where $c$ is the average contact rate of the individual which can be determined by computing the largest eigenvalue of the respective contact matrix

$$M = \begin{bmatrix} c_{n,n} & c_{n,p} & c_{n,d} \\ c_{p,n} & 0 & c_{p,d} \\ c_{d,n} & c_{d,p} & c_{d,d} \end{bmatrix}$$

where $c_{i,j}$ is the contact rate of an individual of type $i$ with an individual of type $j$ (see Appendix Table 1). Let $N_n, N_{hc}$, and $N_p$ be the average number of nurses, doctors, and patients in the hospital population, respectively. Since the total number of contacts of individuals of type $i$ has to be the same as the total number of contacts of an individual of type $j$, $c_{n,p}N_n = c_{p,n}N_p$, $c_{d,p}N_p = c_{p,d}N_d$, and $c_{d,n}N_n = c_{n,d}N_d$ the contact matrix is given by

$$M = \begin{bmatrix} c_{n,n} & c_{n,p} & c_{n,d} \\ c_{n,p}\frac{N_p}{N_n} & 0 & c_{p,d} \\ c_{n,d}\frac{N_d}{N_n} & c_{p,d}\frac{N_d}{N_p} & c_{d,d} \end{bmatrix}$$

The values of the contact rates in the matrix are based on HCW to patient ratios and the number of rounds per shift of HCWs and estimated from our simulations. The respective values can be found in Appendix Table 1. We compared these values to a prospective contact survey of nurses in five hospitals in the German federal state of Bavaria conducted by Bernard and colleagues.[14] The authors reported a median work-related contact rate of $c_n = 34$ of nurses during 24 hours. Most work-related contacts were with patients (51%) or other staff member/other persons (49% = 40% + 9%). Thus, nurses meet approximately 17.3 patients and 16.7 other staff members per 24 hours. The contact rate of nurses with patients per duty shift from our simulations ($c_{n,d}$=19·07) is similar to the reported value by Bernard and colleagues. The contact rates between hospital staff are lower in our simulation than reported in the contact survey (see values in Appendix Table 1) and based on our assumption that contacts between hospital staff decreased during the first wave of the COVID-19 pandemic in the Netherlands.

*Time of infection*
For individuals infected in the community, the time of infection is unknown. For asymptomatic individuals, we assume an infectious period of 14 days and draw the infection time uniformly from 0 to 14 days prior to admission. For individuals that will develop symptoms, we draw an incubation period $t_{inc}$ from the respective distribution (see Appendix Table 1) and then draw the infection time uniformly from 0 to $t_{inc}$ prior to admission. Note that this approach neglects the fact that in an early stage of an outbreak when the epidemic grows at an exponential rate, it is likely that there are many more recently infected individuals.

**Infection control interventions**

We used the model to evaluate the effect of several interventions aimed at HCWs on the hospital epidemic using data from the first wave of the epidemic in the Netherlands but assuming the introduction of a SARS-CoV-2 variant with higher transmissibility in the hospital. As such, our model results show the impact of the interventions on the nosocomial spread of a new variant.
Throughout the simulations, we assume that HCWs use 90% effective PPE in COVID wards and isolate at home immediately upon symptom onset for seven days, after which they return recovered to work. Furthermore, we assume that there is no delay between testing and receiving the COVID-19 test result. This assumption is in particular reasonable for hospital staff tested in the hospital, as they receive their test result within hours (UMCU) and have to self-isolate until they receive the result. Thus, it can be assumed that they do not contribute to the transmission of the virus while waiting for the test result.

*Baseline scenario*
We assumed that HCWs used personal protective equipment (PPE) while working in COVID wards. PPE reduces the transfer of droplets or other body fluids onto HCWs' skin and clothing or directly onto the mucous membranes of the eye or nasopharynx. We define PPE efficacy as the percentage reduction of droplet transfer. Furthermore, we define the effectiveness of PPE as the reduction of infectiousness by a factor upon each contact between an infected and susceptible individual. This reduction factor includes PPE efficacy as well as adherence to adequate PPE use. In our baseline scenario, we assumed that all PPE use was 90% effective. We assumed HCWs do not use PPE when meeting each other in the common room and that per day 95% of the HCWs work in the same ward as during their previous shift.

*Internvetion: PPE in all hospital wards*
In this intervention scenario, we assumed that all HCWs wore 90% effective PPE in all (non-COVID and COVID) wards. Note that no PPE is worn when HCWs meet each other. We performed sensitivity analyses assuming PPE effectiveness of 50% and 70%.

*Intervention: HCW cohorting (no HCW ward change)*
In this intervention scenario, we restricted HCWs to work only in specific wards and did not allow any ward change.

*Intervention: Regular HCW screening*
All HCWs were tested for SARS-CoV-2 either with a) a test with perfect sensitivity every three days, or a test with time-varying sensitivity, b) every three days, or c) every seven days. If tested positive, HCWs were assumed to immediately self-isolate at home for seven days.

*Intevention: HCW contact tracing*
If a HCW developed symptomatic SARS-CoV-2 infection, all contacts in the hospital during a time window of either two or seven days before symptom onset were traced and tested. We will refer to these scenarios as *2-day Contact tracing* and *7-day contact tracing*. For *2-day contact tracing*, contacts were always tested assuming a time-varying test sensitivity. For *7-day contact tracing*, we distinguished between perfect and time-varying sensitivity sub-scenarios. In the perfect

sensitivity sub-scenario, contacts were instantaneously tested on the day of symptom onset of the index (the HCW). In the time-varying test sensitivity sub-scenario, the test was performed on the day of symptom onset if the contact with the index was more than five days ago. Otherwise, it was performed on day five after the contact. Exposed HCWs awaiting tests were assumed to wear PPE during contact with any patient and with other HCWs. In case of a positive test, patients were moved to a COVID ward while infected HCWs were sent home for self-isolation for seven days and replaced by susceptible HCW. We did not model any absences of HCWs with disease symptoms caused by other respiratory pathogens.

*Outcome measures*

We calculated the effective reproduction number $R_E$ (average number of secondary cases caused by an infected individual) to evaluate an intervention's effectiveness in suppressing outbreak expansion in the hospital. We approximated $R_E$ for an average individual (patients and HCWs combined) in the hospital (overall $R_E$) from our simulations by calculating the average number of secondary cases by an infected individual in our model. We further stratified this number by patients, HCWs, and symptom status. The reproduction numbers of patients were calculated for those who will eventually develop symptoms ($R_S^{pat}$) and those who will remain without symptoms ($R_A^{pat}$). Since HCWs were assumed to immediately self-isolate upon symptom onset, we calculated $R$ during pre-symptomatic ($R_S^{hcw}$) and asymptomatic states ($R_A^{hcw}$) only. In order to evaluate the maximum demand on the hospital capacity, we considered the total number of hospital-acquired infections among patients and HCWs over time. In addition, we computed the proportion of absent HCWs due to self-isolation (because of symptom onset or detection via screening or contact tracing) over time. We assessed the efficiency of screening and contact tracing interventions with respect to their positivity rates (proportion of detected infected individuals among tested individuals). We did not include individuals that developed symptoms prior to being tested in the positivity rate calculations since those were already detected and isolated in our model. We determined the proportion of transmissions attributed to different transmission routes (HCW-to-HCW, HCW-to-patient, patient-to-HCW). For every scenario, we calculated the mean and 95% percentiles over 100 simulation runs (95% uncertainty interval). We calculated positivity rates over time merging data from all simulation runs and computed 95% Bayesian beta-binomial credibility intervals.

**Results on transmission routes**

Our results show that for the considered simulation scenarios most of the nosocomial transmissions of the SARS-CoV-2 variant is mainly driven by transmissions between patients and HCWs (Appendix Figure 4). This is expected as we assumed that there is no direct contact between patients and the majority of contacts of HCWs are with patients. Furthermore, for most of the intervention scenarios, over 90% of transmissions occur in non-COVID wards where no use of PPE is assumed in the baseline scenario (Appendix Figure 6). Since in our model infected patients are transferred to COVID wards and infected HCWs are assumed to self-isolate immediately upon symptom onset, most transmissions take place during the pre-symptomatic stage of an infected individual (dark-grey bars in Appendix Figure 7). This is in line with a French study where secondary cases were exposed mainly in the pre-symptomatic phase.[15] When PPE is used throughout the hospital or HCWs are screened assuming a perfect test sensitivity, most transmissions are prevented (Figure 5 of the main text). In particular, transmissions that occur during non-symptomatic states in non-COVID wards are significantly reduced, decreasing their contribution to the overall number of transmissions (Appendix Figure 6-7).

**Results of sensitivity analyses**

We evaluate the changes of our results with respect to changes in our model parameters. We present the results and corresponding plots for the effective reproduction number $R_E$, the total number of nosocomial transmissions, and daily number of absent HCWs. The remaining plots can be found online: https://github.com/htahir2/covid_intra-hospital_model.git.

*PPE effectiveness*

We performed two sensitivity analyses to test the impact of PPE effectiveness values on our results:

a) 50% effective PPE
b) 70% effective PPE

Our sensitivity analyses show that the effective reproduction numbers and the total number of nosocomial transmissions increase with lower PPE effectiveness and decrease with higher PPE effectiveness, in particular for the "PPE in all wards" intervention scenario (compare Appendix Figure 12-13, Appendix 15-16, and Figure 3 of the main text). A similar effect can be observed for the daily percentage of HCW absenteeism (compare Appendix Figure 14, Appendix Figure 15, and Figure 6 of the main text). The relative impact of the different interventions on the reproduction number in comparison to the baseline scenario are similar to what we have observed in our main analysis. The only difference is that for a low value of PPE effectives of 50%, screening every three days with time-invariant perfect sensitivity is more effective in reducing the effective reproduction number, especially for pre-symptomatic HCWs. However, the use of 50% effective PPE in all wards still decreases the effective reproduction number more than the remaining interventions.

*Reproduction number*

We performed a sensitivity analysis to test the impact of equal reproduction numbers of symptomatically and asymptomatically infected individuals on our results (Appendix Figure 18-20). Furthermore, we show the model results for the reproduction numbers resulting from calibrating our model to data on the number of occupied beds by COVID-19 patients at the UMCU (Appendix Figure 21-23). Our sensitivity analyses show that the effective reproduction numbers, the total number of nosocomial transmissions as well as the daily percentage of HCW absenteeism increase with increasing basic reproduction number. In particular, when the reproduction number of asymptomatically infected individuals is as high as the one of symptomatically infected individuals, the respective effective reproduction numbers for asymptomatic patients and HCWs increase. The impact on the overall effective reproduction number is smaller, however, still notable. Qualitatively, our conclusions regarding the relative effect of the considered infection control interventions remain unchanged. For low reproduction numbers as it was the case for the nosocomial spread of the wild-type SARS-CoV-2 variant at UMCU, the numbers of nosocomial transmissions are very small and hence the relative impact of the intervention scenarios in comparison to each other and to the baseline scenario is smaller than for higher reproduction numbers. However, the qualitative conclusions remain unchanged.

*Increased HCW-to-HCW contact rate*

In our main analysis, we assume that HCWs meet other HCWs once every hour. In this sensitivity analysis, we relax this assumption by increasing the contact rates between HCWs to once every 30 minutes and evaluate the impact on our results (Appendix Figure 24-26). The effective reproduction numbers, the total number of nosocomial transmissions, and the daily percentage of HCW absenteeism increase when the contact rate between HCWs is increased. In particular, the effective reproduction numbers for HCWs increase but not those for symptomatic patients (Appendix Figure 24). Qualitatively, our conclusions with respect to the impact of the interventions on the hospital epidemic do not change with respect to this parameter.

*Test sensitivity*

We performed two sensitivity analyses:

a) assuming the test sensitivity to remain at the maximum after reaching its peak and
b) reducing the test sensitivity curve of the main analysis by 15%.

The respective test sensitivity curves varying from time since infection are shown in Appendix Figure 1. There are only minor differences in our results for both sensitivity scenarios (Appendix Figure 27-32 vs Figure 3 and Figures 5-6 of the main text).

### Implementation

The model was built using *Python* (version 3.6) using the library *Mesa* which is an open source agent-based modelling framework.[16] The code is available from: https://github.com/htahir2/covid_intra-hospital_model.git

*Initialization*

The model is initialized with non COVID patients admitted to normal ICU and normal wards. We assume 50% of the rooms (beds) in the normal ICU and normal wards are free at the moment of model initialization. There are three duty shifts in a day and in these duty shifts, HCWs (nurses and doctors) are assigned to all wards in the hospitals. We assume that the number of HCWs in the hospital remains constant throughout the simulation period. Every patient agent has its own unique characteristics such as ID, ward and room number, LoS, and disease state. Depending on the ward (normal ICU or normal ward), every patient is assigned a LoS from the given distributions at the time of admission (Appendix Table 1). HCWs also have unique characteristics such as ID, ward to which HCW is assigned, duty shift in which HCW is working, HCW wearing PPE or not, time being absent from work due to quarantine, and disease state. A patient or a HCW can only be in one of the following disease states at a particular moment: susceptible, exposed, moderate symptomatic, severe symptomatic, asymptomatic, or recovered. However, at the model initialization, all patients and HCWs are in susceptible state.

*Study period*

The simulation is run for 239 days in total, with an initial period of 59 days to get a stable non-COVID patient population in the hospital. The first symptomatic COVID-19 patient is admitted to the hospital on day 60.

*Scheduling*

A time step in our agent-based model represents 10 minutes. The following processes occur during the run time:

*New patient arrival*

Given an average daily patient arrival rate for the UMCU (Appendix Table 1), patients arrive at the hospital following a Poisson process and are randomly admitted to normal ICU and normal wards. The majority of these new daily patients are in a susceptible state but as mentioned earlier in the section "*Importation from community*", we use a community-prevalence-dependent, age-specific importation rate of exposed and asymptomatic patients into the hospital. Therefore, some patients from the daily new patient arrivals come in an exposed or asymptomatic state. Since the disease status of such patients is not known at the time of admission, they are admitted to normal ICU or normal wards. Depending on the patient ward (normal ICU or normal ward), the LOS is drawn from the appropriate distributions (Appendix Table 1).
The first symptomatic COVID-19 patient is admitted to the hospital on day 60 (based on UMCU COVID-19 admission data). Known symptomatic patients are admitted to either COVID wards or COVID ICUs depending on the severity of their symptoms. Moderate symptomatic patients are admitted to COVID wards, whereas severe symptomatic patients are admitted to COVID ICUs. For symptomatic COVID patients admitted to the hospital, LOS of the individual patient is sampled directly from the UMCU data.

*Patient discharge*
The remaining LOS of every patient is decremented at every time step. When the LOS of a patient reaches zero, the patient is discharged from the hospital. We do not model patient deaths.

*HCWs visiting patients*
At each time step, HCWs (nurses and doctors) from every ward visit individual patients. This is the moment where a contact between HCW and patient takes places. Single HCWs visit one single patient in one time step. When a HCW and a patient meet in a room and if one of them (patient or HCW) is infected with SARS-CoV-2, a transmission event can take place. As explained earlier in the section "*Transmission*", a Bernoulli trial using the average probability of transmission per contact with a susceptible person is carried out. If a trial is successful, a susceptible individual acquires infection. The patient may be in an exposed state and develop symptoms after an incubation period or be asymptomatically infected. All exposed individuals in the model follow symptomatic route whereas asymptomatic individuals follow asymptomatic route. To decide on this for a patient, a random number is drawn and if it is less than the specified proportion of asymptomatic patients ($P_A^p$), the patient state is changed to asymptomatic, otherwise exposed. Similarly, for a HCW if the random number is smaller than the specified proportion of asymptomatic HCWs ($P_A^h$), the HCW's state is changed to asymptomatic, otherwise the state of the susceptible HCW is changed to exposed. Infectiousness from symptomatic or asymptomatic individuals over time is different as explained earlier in the section "Infectiousness". For exposed patients and HCWs, an incubation period $t_{inc}$ is drawn from the Gamma distribution ($s(\tau)$) with a mean of 5·5 days.

*Exposed to infection*
For every individual in an exposed state, the incubation period is then decremented by 1 at every time step. When the incubation time of an exposed individual (patient or HCW) reaches zero, the individual is either moved to moderate or severe symptomatic state depending on the proportion of individuals developing severe symptoms ($P_s$).
For patients who develops severe symptoms, a sample is drawn from the LoS of severely infected patients (based on UMCU data) and the LoS of the respected patient is extended accordingly. Severely infected patients are then moved to one of the COVID-ICUs, whereas moderately infected patients are moved to any of the COVID wards.
A HCW who develops symptoms is sent home for an isolation period of seven days, and a susceptible HCW is added in the same ward and shift as a replacement.

*Infection to recovery*
Moderately or severely infected patients are assumed to recover either at the time of discharge or after a maximum period of 14 days or 35 days, respectively. Asymptomatically infected patients recover at the time of discharge or after maximum period of 14 days.
Moderately or severely infected HCWs return to work as recovered after the end of the isolation period of seven days. When a recovered HCW returns to work to a specific ward, a HCW is removed from that ward as we assume a constant HCW population. To do that, we first look in the list of susceptible HCWs in the same ward and duty shift. If that list is not empty, a susceptible HCW is randomly chosen and removed from the hospital HCWs population. If there is no susceptible HCWs in that ward and duty shift, we look further into the list of exposed and asymptomatic HCWs in the same ward and duty shift. If that list is not empty, then a randomly chosen exposed or asymptomatic HCW is removed from the hospital population. If this is also not successful, we randomly choose a recovered HCW from the same ward and duty shift and remove him from the hospital population. These steps are required to maintain a constant HCW population.

*HCWs meeting in the common areas*
Every hour, two HCWs meet in the common areas of every ward. For this, we randomly pick two HCWs from the list of HCWs working in a ward in a shift. If one of the randomly chosen HCW is exposed or asymptomatic and the other HCW is susceptible, a transmission event can take place. As explained earlier in the section "*HCWs visiting patients*", a Bernoulli trial using the average probability of transmission per contact with a susceptible person is carried out. If the trial is

successful, the susceptible HCW acquires infection and can either enter into an exposed state or an asymptomatic state. Next, a random number is drawn, and if smaller than the specified proportion of asymptomatic HCWs ($P_A^h$), the HCW state is changed to asymptomatic, otherwise the state of the HCW is changed to exposed. For an exposed HCW, an incubation period $t_{inc}$ is drawn from the Gamma distribution ($s(\tau)$) with a mean of 5·5 days.

*HCWs ward swap*

Before the start of each day, a certain proportion of HCWs ($W_h$) are randomly selected and their wards are changed (Appendix Table 1). In order to do that, we first loop over the list of active HCWs (the ones that are not isolated at home). Since we change wards of two HCW at the same time (ward swapping), we draw a random number for every HCW. If the random number is less than $W_h/2$, we select that specific HCW (HCW-A) to be moved to another ward. The next step is to find another HCW (HCW-B) in a ward different from the ward of HCW-A. For this, we again make a list of all the active nurses (if HCW-A was a nurse) or active doctors (if HCW-A was a doctor), and then randomly pick a HCW (HCW-B). Once we have selected both HCW-A and HCW-B, we can now swap the wards, duty shifts and PPE of the both HCWs. We repeat the above process for all active HCWs.

**Appendix Table 1. Model parameters**

| Name | Symbol | Description | Distribution/Value* | Source |
|---|---|---|---|---|
| Incubation period | $s(\tau)$ | Time between infection and symptom onset | Gamma distribution shape=5·807 scale = 0·948 mean = 5·510 SD = 2·284 | Lauer and colleagues[5] |
| Generation time | $\omega(\tau)$ | Time between becoming infected and subsequent onward transmission events | Weibull distribution Shape = 2.826 Scale = 6.839 mean = 6 days | Grassly and colleagues[9] |
| Proportion of asymptomatic infections among infected patients | $P_A^p$ | Proportion of infected patients that will experience no symptoms | 20% | Buitraga-Garcia and colleagues[7] |
| Proportion of asymptomatic infections among infected HCWs | $P_A^h$ | Proportion of infected HCWs that will experience no symptoms | 31% | Buitraga-Garcia and colleagues[7] |
| Proportion of severe symptomatic individuals | $P_s$ | Proportion of exposed individuals that will develop severe symptoms | 20% | Wu and colleagues[17] |
| Reproduction number of asymptomatic infectees for wild-type variant | $R_A^W$ | Mean number of infections caused by an individual asymptomatically infected with the wild-type SARS-CoV-2 variant | 0·5 | Calibrated to UMCU data |
| Reproduction number of symptomatic infectees for wild-type variant | $R_S^W$ | Mean number of infections caused by an individual symptomatically infected with the wild-type SARS-CoV-2 variant | 1·25 | Calibrated to UMCU data |
| Reproduction number of asymptomatic infectees for new virus variant | $R_A$ | Mean number of infections caused by an individual asymptomatically infected with the SARS-CoV-2 variant | 0·8 (1·95) | Based on $R_A^W$ with 56% higher transmissibility, varied in sensitivity analysis |
| Reproduction number of symptomatic infectees for new virus variant | $R_S$ | Mean number of infections caused by an individual symptomatically infected with the SARS-CoV-2 variant | 1·95 | Based on $R_A^W$ with 56% higher transmissibility |
| Peak sensitivity of RT-PCR test for SARS-CoV-2 | | Maximum sensitivity of the RT-PCR diagnostic test for SARS-CoV-2 | 93·1% (79%) | Grassly and colleagues[9], varied in sensitivity analyses |
| Proportion of HCWs that work in the same ward as during their previous shift | $W_h$ | Proportion of HCWs that change wards they were working in their previous shift | 95% (baseline) 100% (intervention) | Assumed |
| PPE effectiveness | | Reduction in infectiousness upon contact between an infected and susceptible individual (includes PPE efficacy and adherence) | 0% (50%, 70%) | Assumed |
| Isolation period for HCWs | | Amount of time HCWs have to isolate after symptom onset or after being detected by screening or contact tracing | 7 days | Assumed |
| Recovery time for asymptomatic infection | $\gamma_A$ | Mean duration of an asymptomatic infection | 14 days | Assumed |

| | | | | |
|---|---|---|---|---|
| Recovery time for symptomatic (moderate, severe) infection | $\gamma_S$ | Mean duration of a symptomatic infection | 14 days<br>35 days | Liu and colleagues[6] |
| LoS of non-COVID patients in ICU | | | Lognormal<br>meanlog = 0·37<br>sdlog = 0·82 | Fitted distributions to UMCU data from 2014-2017 |
| LoS of non-COVID patients in normal ward | | | Weibull<br>shape = 0·92<br>scale = 4·18 | Fitted distributions to UMCU data from 2014-2017 |
| LoS of moderately infected patients | | | Gamma<br>shape = 1·59<br>scale = 0·05 | Fitted distributions to UMCU data from 2020 |
| LoS of severely infected patients | | | Gamma<br>shape = 1·88<br>scale = 0·25 | Fitted distributions to UMCU data from 2020 |
| Patient-nurse ratio | | 1:1 (COVID ICU),<br>2:1 (COVID ward),<br>1:1 (Normal ICU),<br>4:1 (Regular ward) | | Assumed |
| Patient-doctor ratio | | 6:1 (COVID ICU, COVID ward, Normal ICU),<br>10:1 (Regular ward) | | Assumed |
| Time spent with patient (ward dependent) | | Min 10 minutes,<br>Max 30 minutes | | |
| Duty shifts of HCWs per day | | 3 shifts | | |
| Rounds per shift | | Nurses: 6<br>Doctors: 2 | | |
| Contact rates (per shift) | $c_{n,n}$ | Average number of contacts between nurses | 4·6 | From simulation |
| | $c_{n,p}$ | Average number of contacts that a nurse has with patients | 19·07 | |
| | $c_{n,d}$ | Average number of contacts that a nurse has with doctors | 3 | |
| | $c_{p,n}$ | Average number of contacts that a patient has with nurses | 6 | |
| | $c_{p,d}$ | Average number of contacts that a patient has with doctors | 2 | |
| | $c_{d,d}$ | Average number of contacts between doctors | 0·43 | |
| | $c_{d,p}$ | Average number of contacts a doctor has with patients | 17·4 | |
| | $c_{d,n}$ | Average number of contacts a doctor has with nurses | 3 | |
| Daily arrival rate of non-COVID patients | | Number of patients that arrive at the hospital per day for non-COVID related reasons | 40 patients per day | Based on UMCU data from 2014-2017 assuming 50% decrease during study period |
| HCW population | | Constant number of HCWs working in the hospital per day | 870 | |

* Mean or median values were used from literature; range was used in the sensitivity analyses.

**Appendix Figure 1. PCR test sensitivity over time since infection**. The empirical estimate based on published data as reported by Grassly and colleagues is shown (black dots).[9] Two sensitivity analyses were performed: 1) assuming the test sensitivity remains at the maximum after reaching its peak (dark grey dashed) and 2) test sensitivity curve of the main analysis scaled down to 85% (light grey dotted).

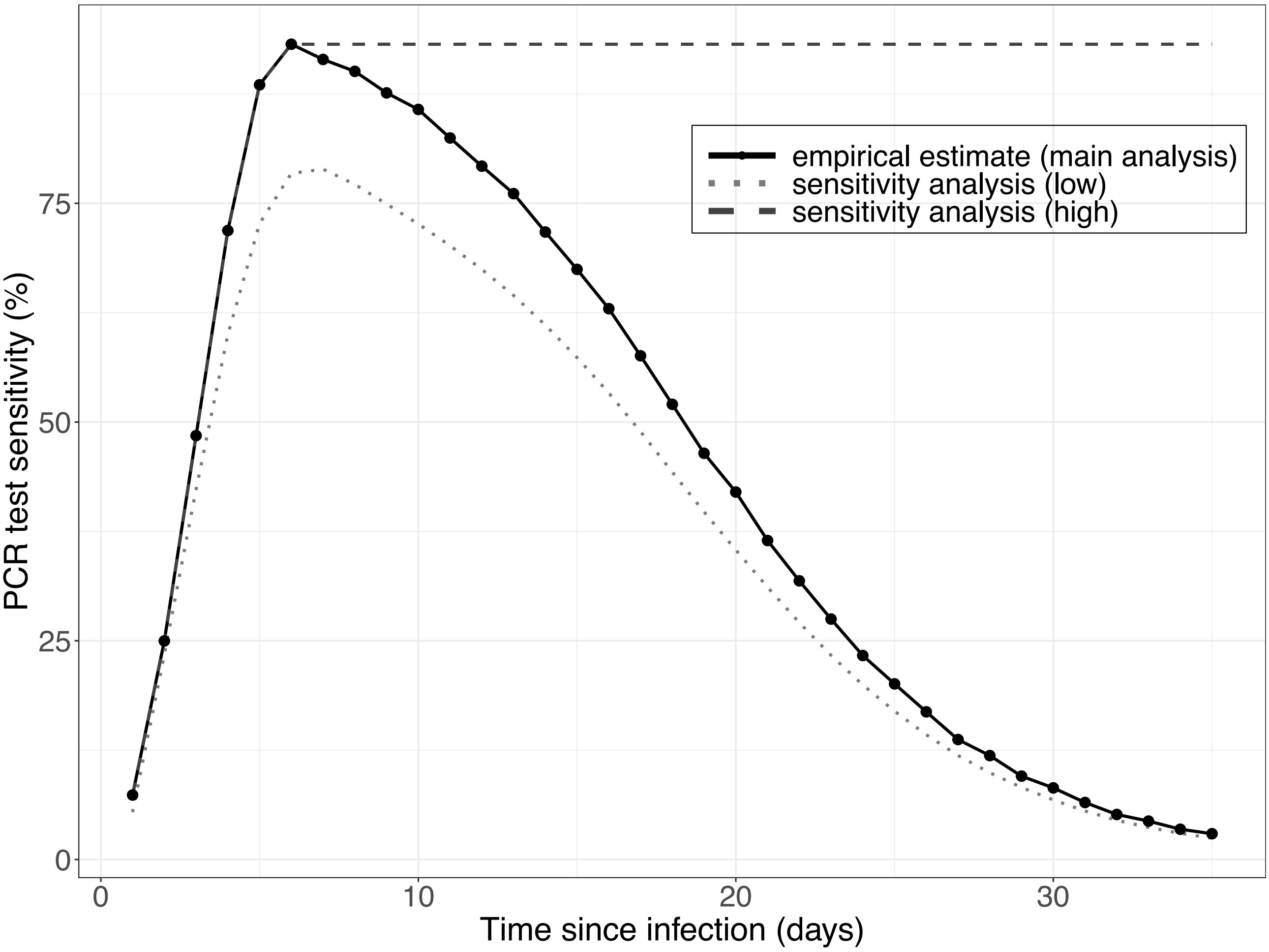

**Appendix Figure 2. Proportion of detected nosocomial transmissions of the SARS-CoV-2 variant for each simulation scenario.** Results shown are based on $R_S$=1·95 and $R_A$=0·8 (reproduction numbers for the SARS-CoV-2 variant with 56% higher transmissibility with respect to the wild-type SARS-CoV-2 variant). The colored rectangular bars with black borders represent the mean proportion of nosocomial infections detected in the hospital due to symptom onset or detection by an intervention (over 100 simulation runs). The lighter colored bars represent the remaining proportions of nosocomial transmissions undetected. These comprise patients infected with SARS-CoV-2 who are discharged to community in a pre-symptomatic or asymptomatic state. The grey error bars the respective 95% uncertainty intervals.

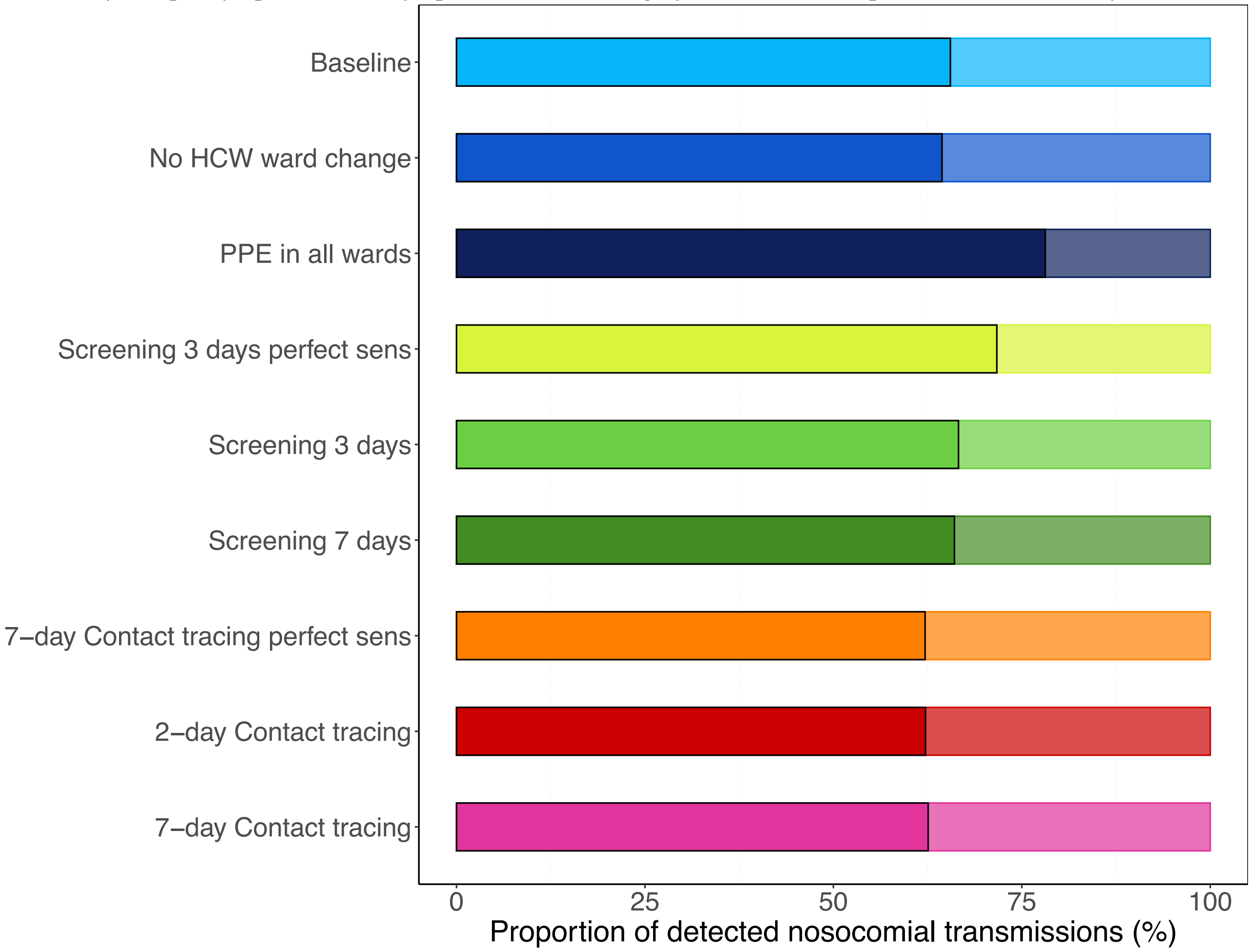

**Appendix Figure 3. Peak numbers of nosocomial infections with the SARS-CoV-2 variant among patients and HCWs during the hospital epidemic for each simulation scenario.** Results shown are based on $R_S$=1·95 and $R_A$=0·8 (reproduction numbers for the SARS-CoV-2 variant with 56% higher transmissibility with respect to the wild-type SARS-CoV-2 variant). The colored rectangular bars represent the mean numbers (over 100 simulation runs) and the grey error bars the respective 95% uncertainty intervals.

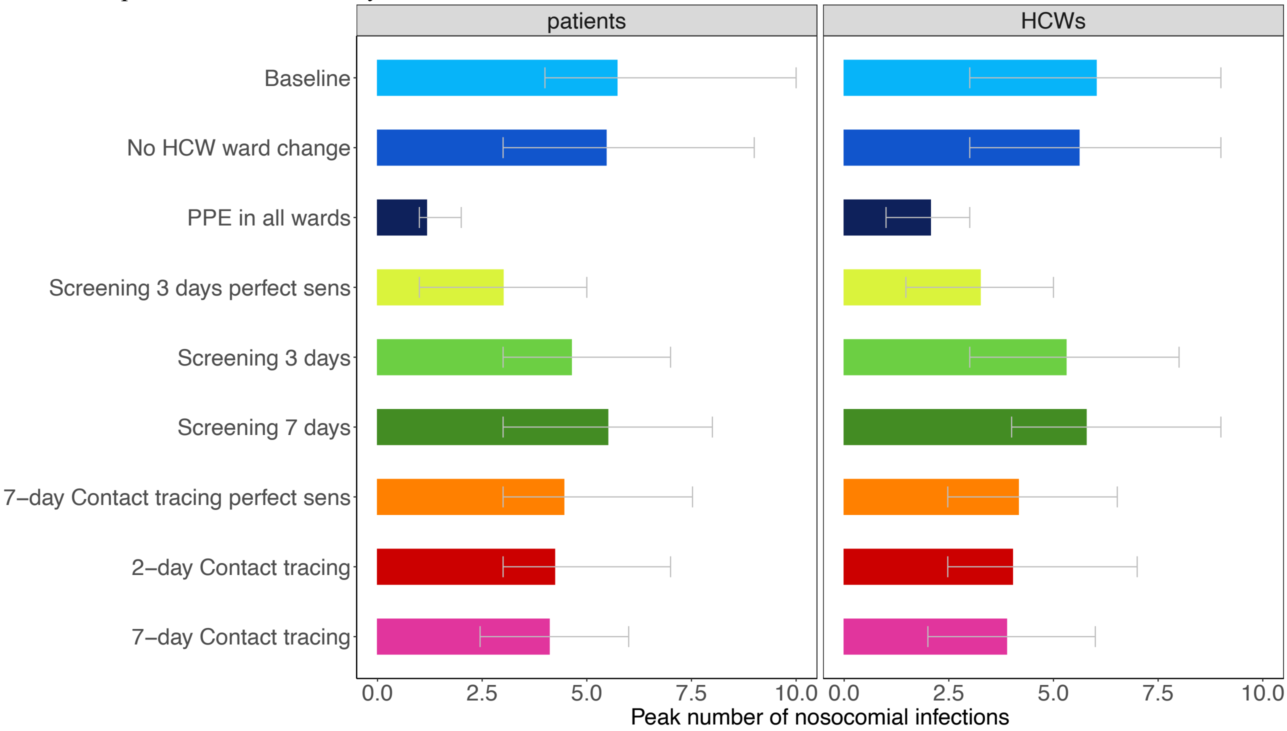

**Appendix Figure 4. Transmission route contributions for nosocomial transmissions of the SARS-CoV-2 variant for each simulation scenario**. Three different transmission routes are considered: From patient to HCW (Patient-HCW), from HCW to patient (HCW-patient), and from HCW to HCW (HCW-HCW). The colored rectangular bars represent the mean percentage of transmissions (averaged over 100 simulations) due to the respective transmission route for each simulation scenario. The grey error bars represent the respective 95% uncertainty intervals. For screening every 3 days and 7-day contact tracing, we considered two different test sensitivity scenarios: time-invariant perfect test sensitivity (perfect sens) and time-varying imperfect test sensitivity.

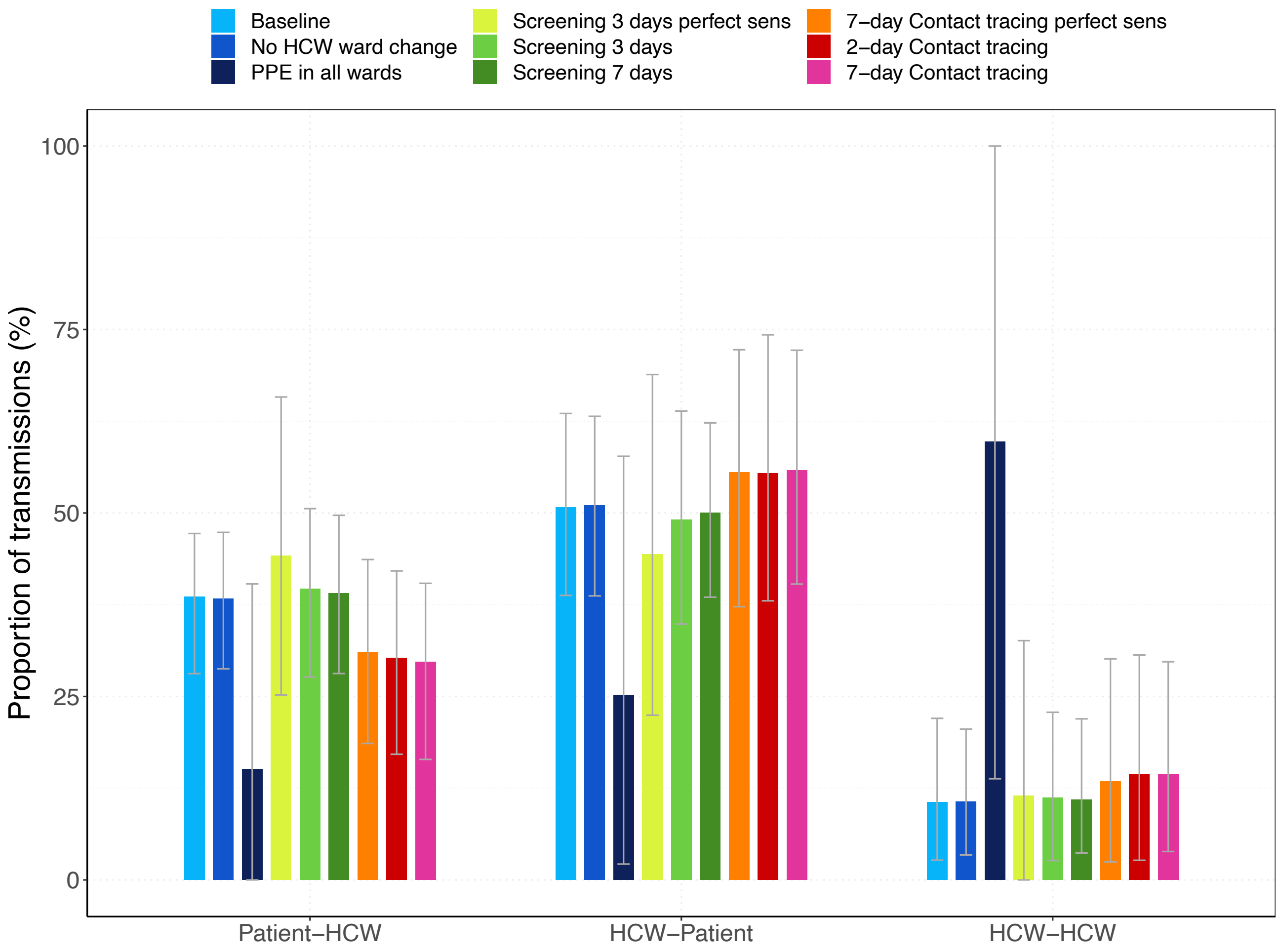

**Appendix Figure 5. Proportion of transmissions from HCWs and from patients for each simulation scenarios.** Mean percentage of total transmissions (averaged over 100 simulation runs) that occurred from HCWs vs from patients are shown in stacked bar plots. Results shown are based on $R_S$=1·95 and $R_A$=0·8 (reproduction numbers for the SARS-CoV-2 variant with 56% higher transmissibility with respect to the wild-type SARS-CoV-2 variant).

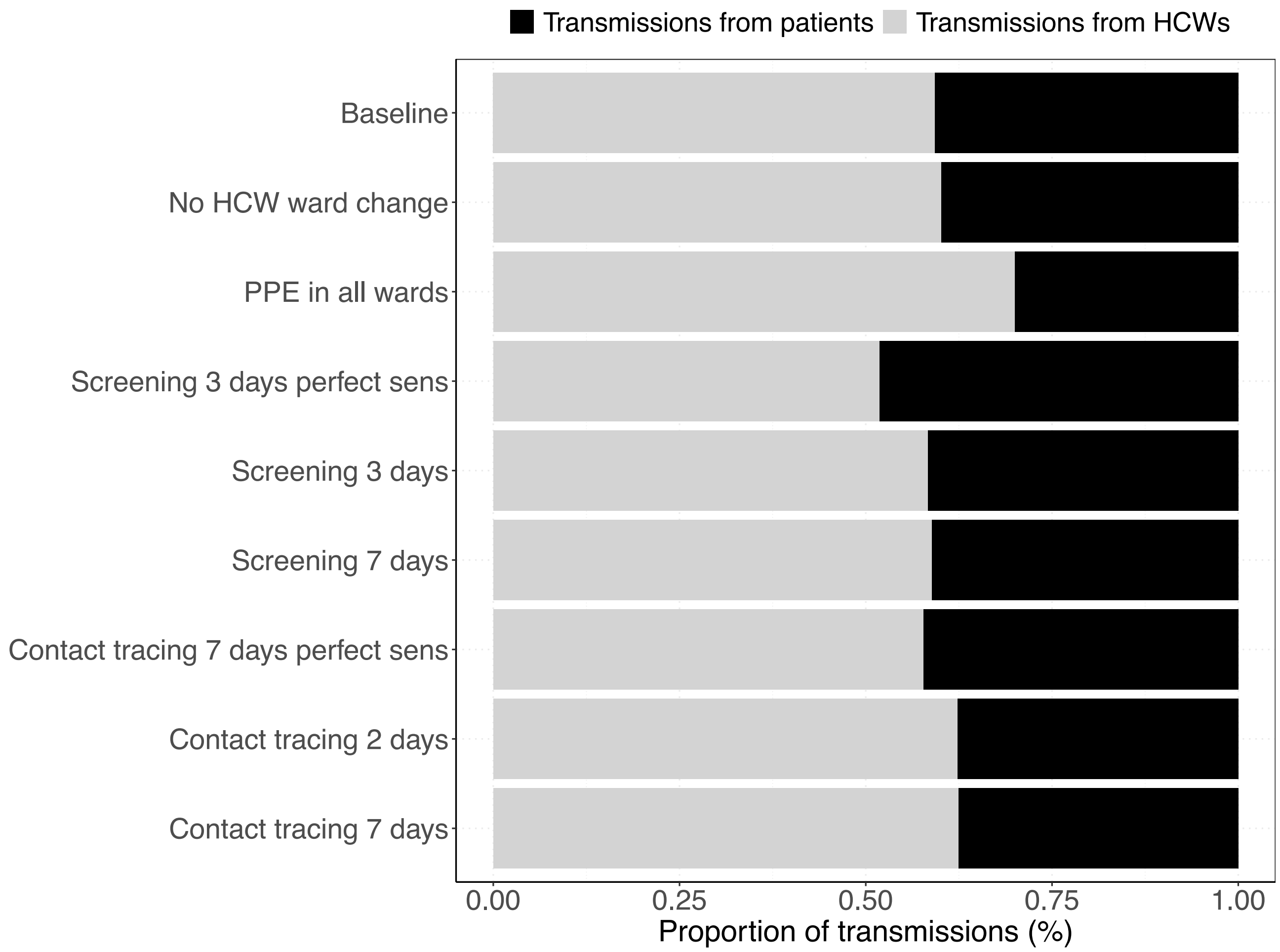

**Appendix Figure 6. Proportion of nosocomial transmissions in COVID- and non-COVID wards for each simulation scenarios.** The mean percentages of nosocomial transmissions (averaged over 100 simulation runs) in COVID and non-COVID wards are shown in stacked bar plots. Results shown are based on $R_S$=1·95 and $R_A$=0·8 (reproduction numbers for the SARS-CoV-2 variant with 56% higher transmissibility with respect to the wild-type SARS-CoV-2 variant).

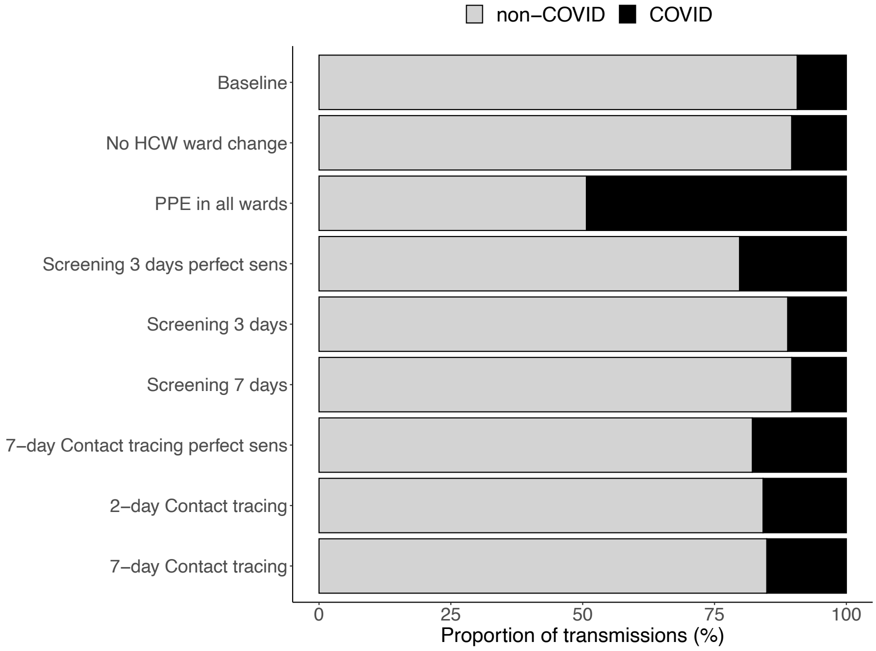


**Appendix Figure 7. Proportion of transmissions during different infection states for each simulation scenario.** The mean percentages of transmissions (averaged over 100 simulations) that occurred while the infected individual was in an asymptomatic, pre-symptomatic, or symptomatic state are shown in stacked bar plots. Results shown are based on $R_S$=1·95 and $R_A$=0·8 (reproduction numbers for the SARS-CoV-2 variant with 56% higher transmissibility with respect to the wild-type SARS-CoV-2 variant).

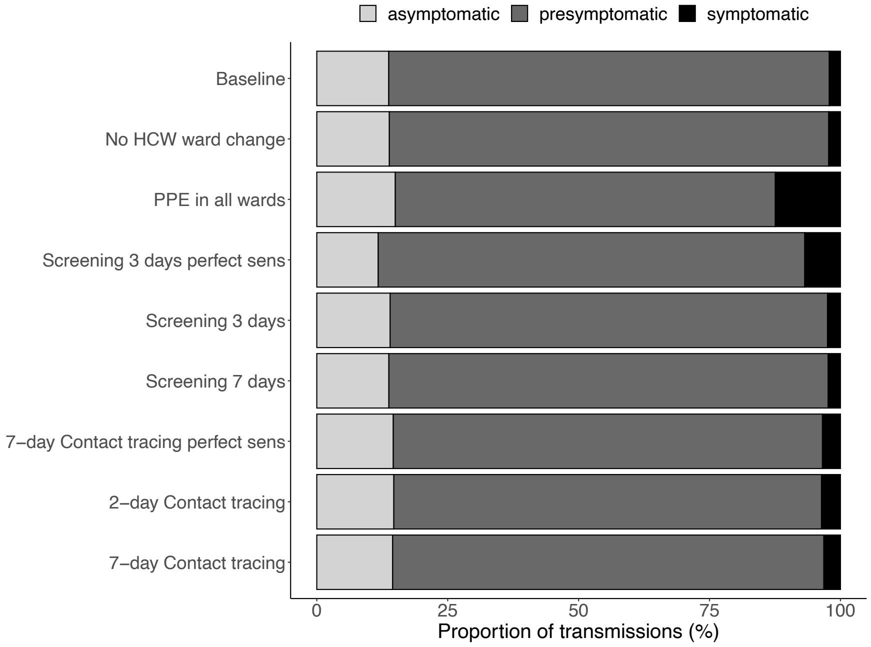

**Appendix Figure 8. Positivity rate of screening interventions for different prevalence ranges.** Results shown are based on $R_S$=1·95 and $R_A$=0·8 (reproduction numbers for the SARS-CoV-2 variant with 56% higher transmissibility with respect to the wild-type SARS-CoV-2 variant). **(**A) Screening every three days with constant perfect test sensitivity. (B) Screening every three days with imperfect, time-varying test sensitivity. (C) Screening every seven days with imperfect, time-varying test sensitivity. On each day when HCWs were screened, the number of positive tested HCWs among the total number of screened HCWs is computed. The prevalence values on the day when HCWs were screened is divided into categories. For each prevalence category, the positivity rate was computed by the total number of positive tested HCWs divided by the total number of screened HCWs (merging values of all simulations) and is shown as a point. The error bars represent the 95% binomial proportion confidence intervals.

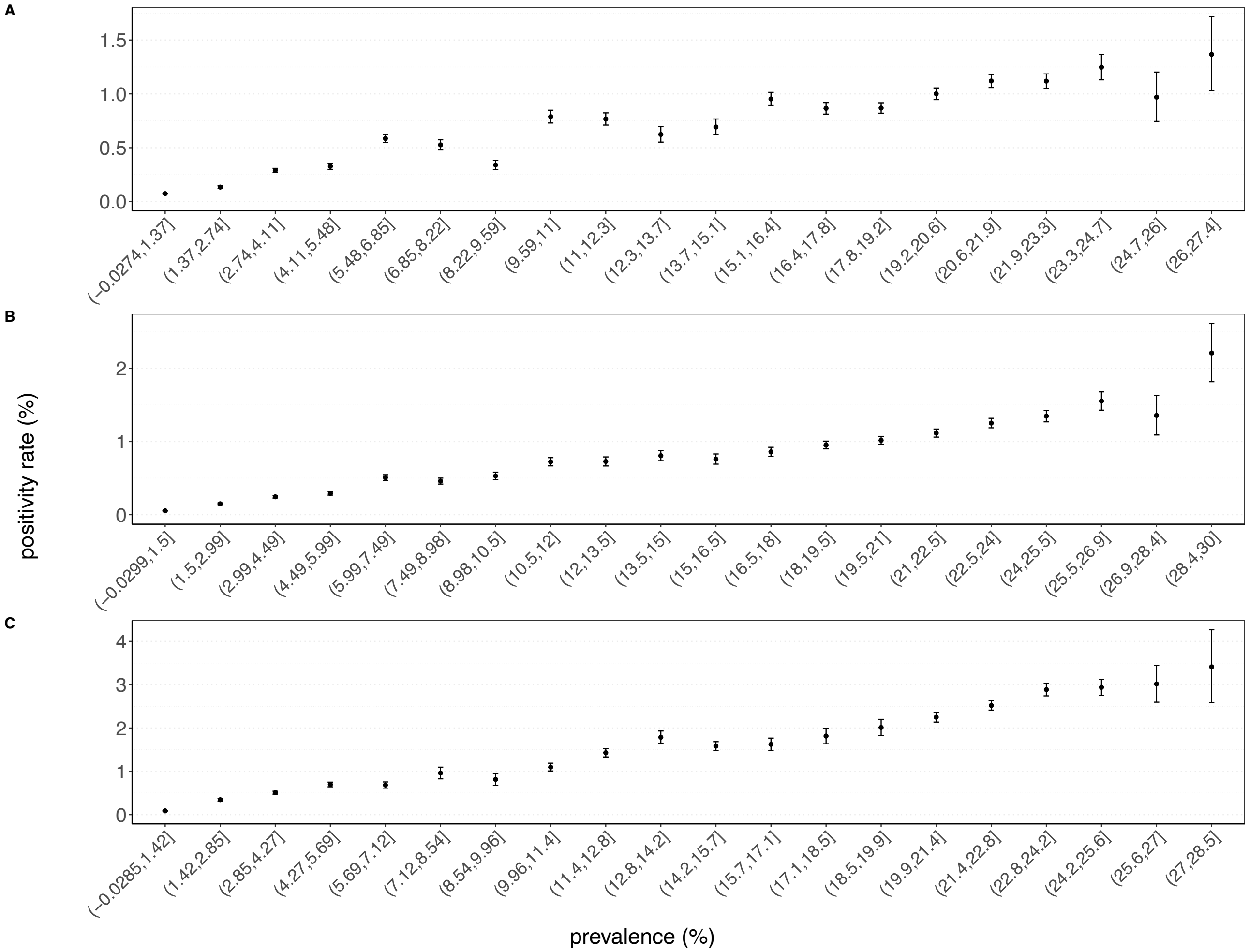

**Appendix Figure 9. Positivity rate of contact tracing interventions for different prevalence ranges.** Results shown are based on $R_S$=1·95 and $R_A$=0·8 (reproduction numbers for the SARS-CoV-2 variant with 56% higher transmissibility with respect to the wild-type SARS-CoV-2 variant). (A) Contact tracing of contacts two days prior to symptom onset with perfect test sensitivity. (B) Contact tracing of contacts two days prior to symptom onset with time-varying imperfect test sensitivity. (C) Contact tracing of contacts seven days prior to symptom onset with time-varying, imperfect test sensitivity. For each index case (symptomatically infected HCW), the number of positive contacts and total number of contacts that are traced is computed in each simulation. The prevalence values on the day when an index case was traced, is divided into categories. For each prevalence category, the positivity rate is computed by the total number of positive divided by the total number of traced contacts (all simulations merged) and is shown as a point. The error bars represent the binomial proportion confidence interval.

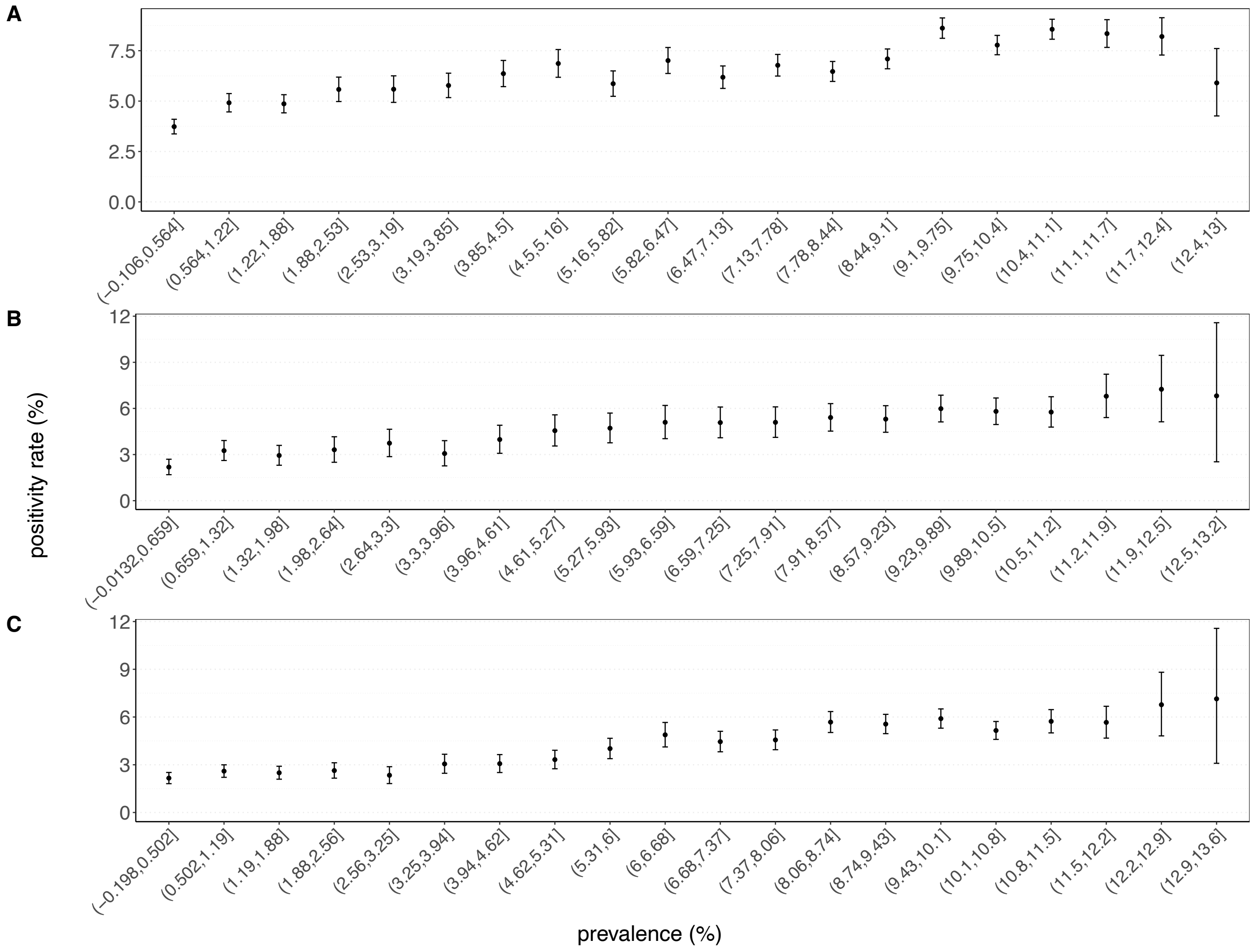

**Appendix Figure 10. Number of patients admitted to UMCU with a SARS-CoV-2 infection between 27 February and 2 August 2020.** Patients were either admitted to an ICU or to another ward in the hospital.

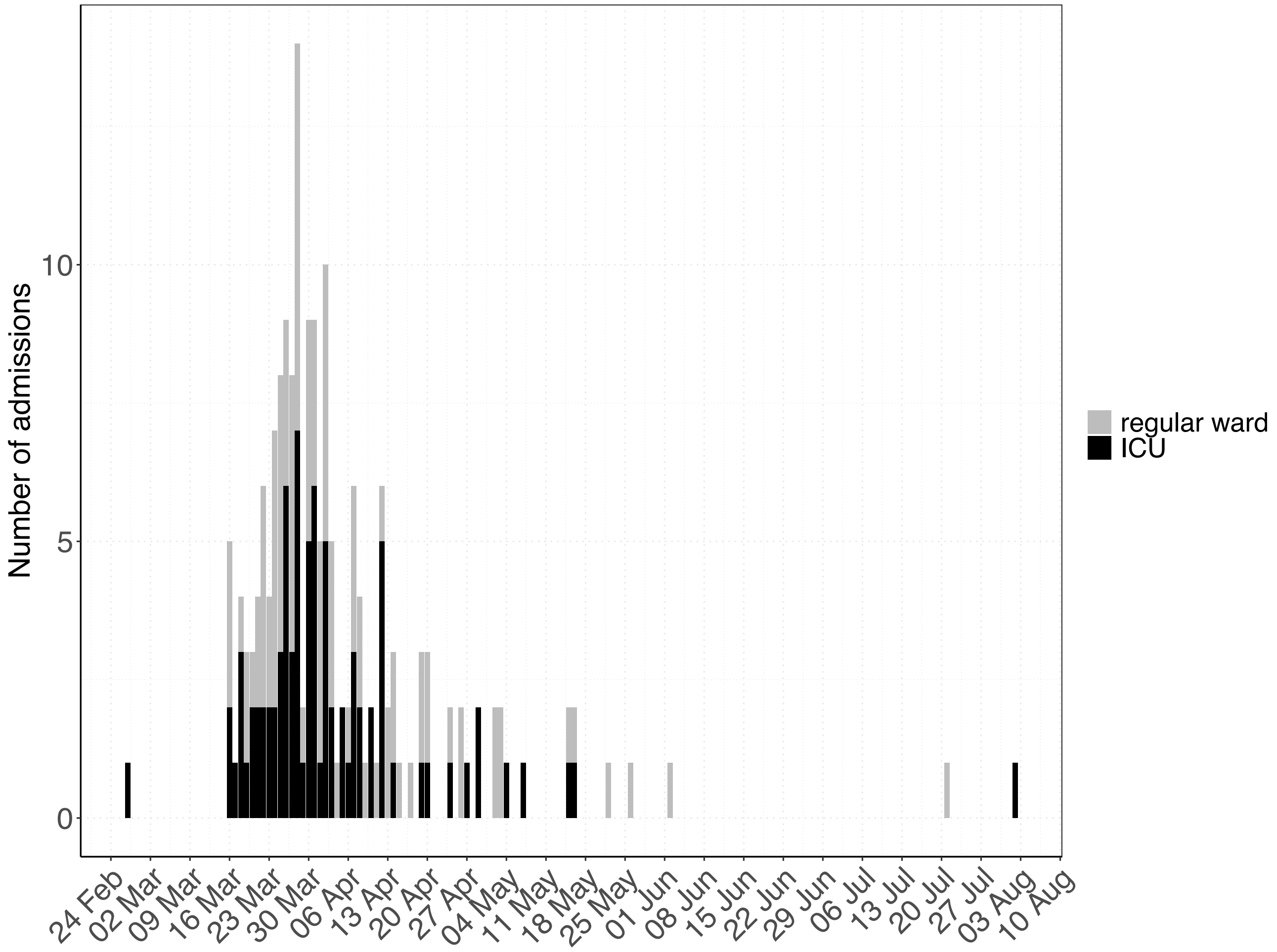

**Appendix Figure 11. Length of stay data of UMCU and fitted distributions for non-COVID and COVID patients in the hospital.** (A)-(B) Histograms show the length of stay distributions for patients admitted to the UMCU between 2014 and 2017. The bold lines represent the fitted probability distributions. The length of stay of patients admitted for non-COVID reasons to the ICUs and to normal wards follow a lognormal distribution and Weibull distribution, respectively. (C)-(D) Histograms show the length of stay distributions for patients admitted with a SARS-CoV-2 infection to the UMCU between 27 February and 24 August 2020. The bold lines represent the fitted probability distributions. The length of stay of patients admitted with a SARS-CoV-2 infection to ICUs and to normal wards follow gamma distributions. The parameters of the probability distributions can be found in Appendix Table 1.

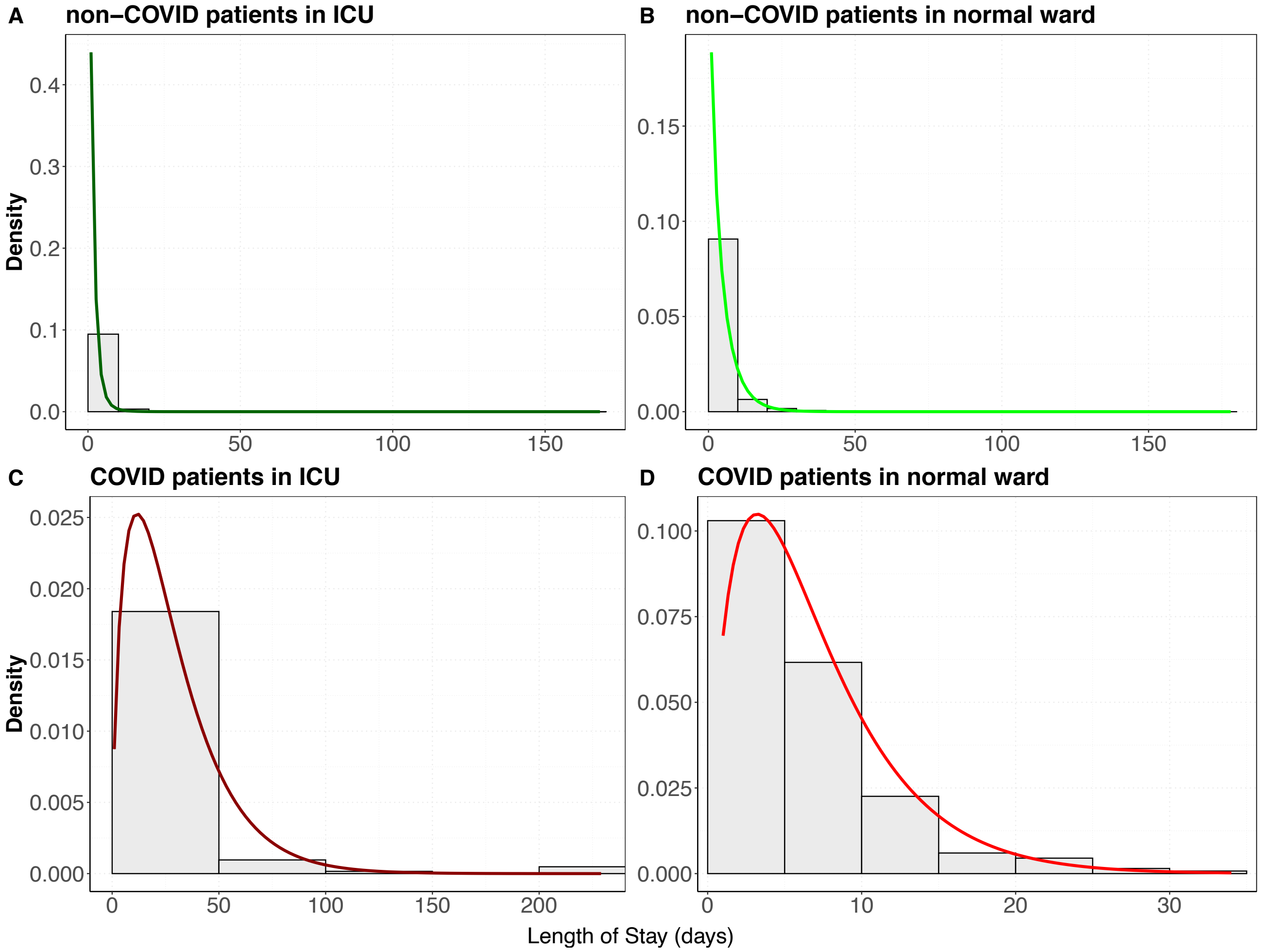

**Appendix Figure 12. Effective reproduction numbers for the nosocomial spread of the SARS-CoV-2 variant for each simulation scenario assuming 50% effective PPE.** Results shown are based on $R_S$=1·95 and $R_A$=0·8 (reproduction numbers for the SARS-CoV-2 variant with 56% higher transmissibility with respect to the wild-type SARS-CoV-2 variant). (A) For each simulation scenario, violin and boxplots of the overall reproduction numbers (for pre-/symptomatic and asymptomatic patients and HCWs combined) are shown (over 100 simulations). (B) For each simulation scenario, violin and boxplots of the reproduction numbers for pre-/symptomatic and asymptomatic individuals are shown (over 100 simulations). Note that since HCWs are assumed to immediately self-isolate upon symptom onset, the reproduction number is assigned to the pre-symptomatic state. The horizontal dashed line represents a reproduction number of 1. For screening every three days and contact tracing seven days prior to symptom onset of SARS-CoV-2 infected HCWs, we considered two different test sensitivity scenarios: time-invariant perfect test sensitivity (perfect sens) and imperfect test sensitivity varying from time since infection.

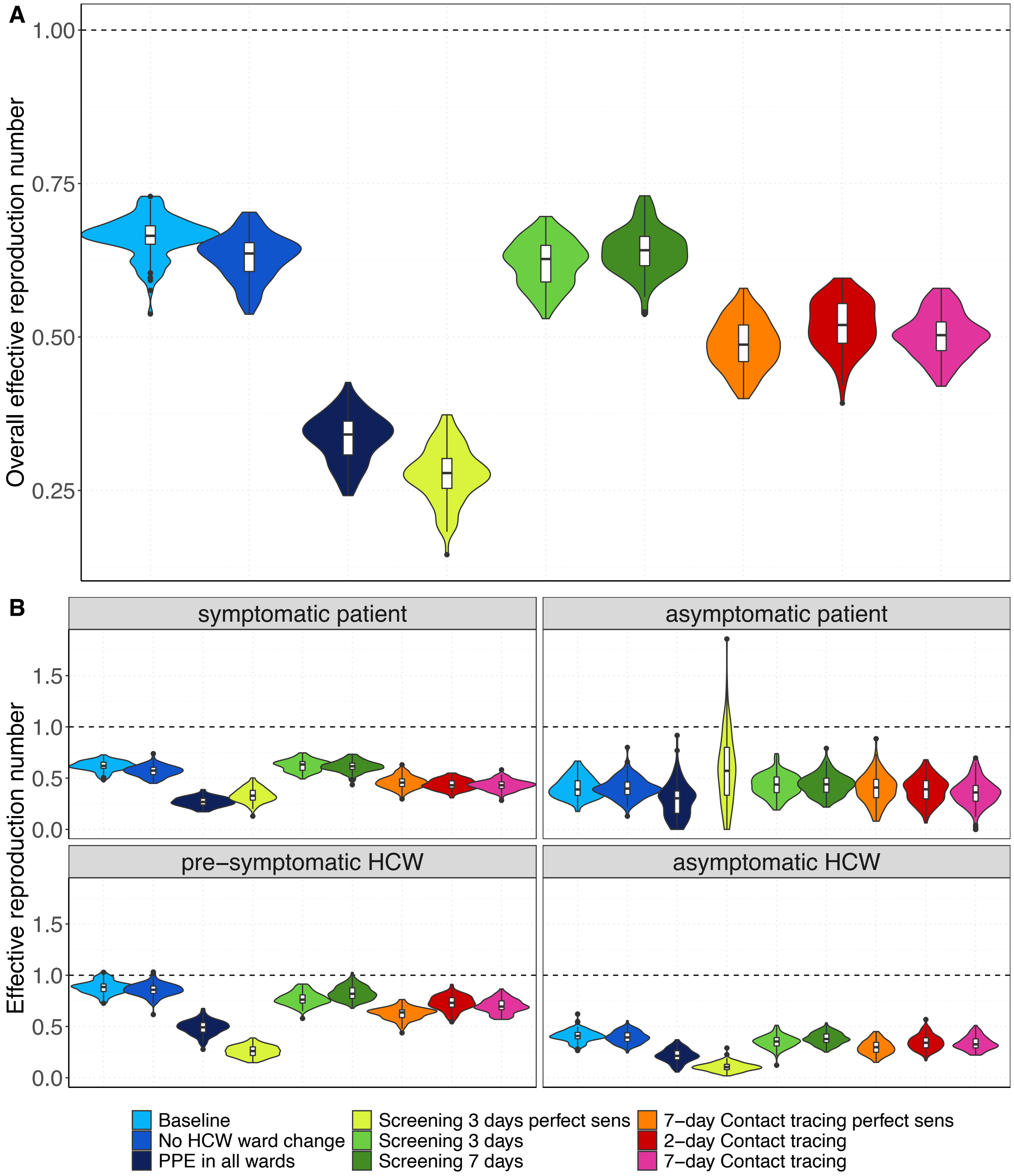

**Appendix Figure 13. Number of nosocomial transmissions of the SARS-CoV-2 variant for each simulation scenario assuming 50% effective PPE.** Results shown are based on $R_S$=1·95 and $R_A$=0·8 (reproduction numbers for the SARS-CoV-2 variant with 56% higher transmissibility with respect to the wild-type SARS-CoV-2 variant). The full rectangular bar height represents the mean total number of nosocomial transmissions during the whole study period (over 100 simulation runs). The grey error bars represent the corresponding 95% uncertainty intervals. Patients that acquire a SARS-CoV-2 nosocomial infection may be diagnosed in the hospital (due to symptom onset during hospital stay or due to detection by an intervention) or discharged to the community in a pre-symptomatic or asymptomatic state. The rectangular bars with the black border represent the mean number of individuals (patients and HCWs) infected with SARS-CoV-2 and diagnosed in the hospital. The lighter rectangular bars represent the remaining mean number of patients discharged to community undiagnosed. For screening every 3 days and 7-day contact tracing, we considered two different test sensitivity scenarios: time-invariant perfect test sensitivity (perfect sens) and time-varying imperfect test sensitivity.

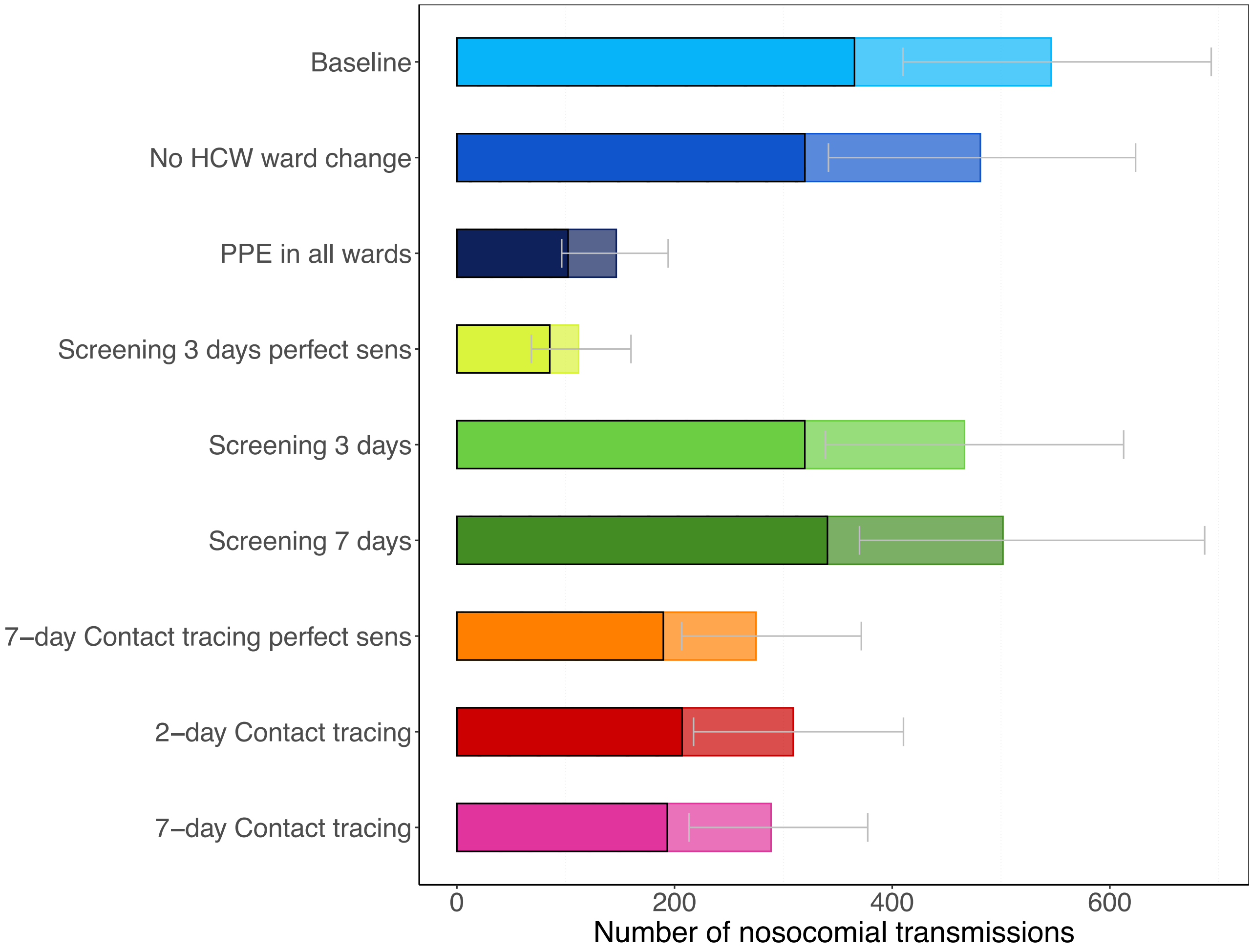

**Appendix Figure 14. Daily percentage of absent HCWs during the hospital epidemic for each simulation scenario assuming 50% effective PPE.** Results shown are based on R_S=1·95 and R_A=0·8 (reproduction numbers for the SARS-CoV-2 variant with 56% higher transmissibility with respect to the wild-type SARS-CoV-2 variant). The 7-day moving average of the mean percentage (over 100 simulation runs) of HCWs absent from work due to symptom onset or a detected SARS-CoV-2 infection screening or contact tracing is shown. For screening every 3 days and contact tracing 7 days prior to symptom onset of SARS-CoV-2 infected HCWs, we considered two different test sensitivity scenarios: time-invariant perfect test sensitivity (perfect sens) and time-varying imperfect test sensitivity.

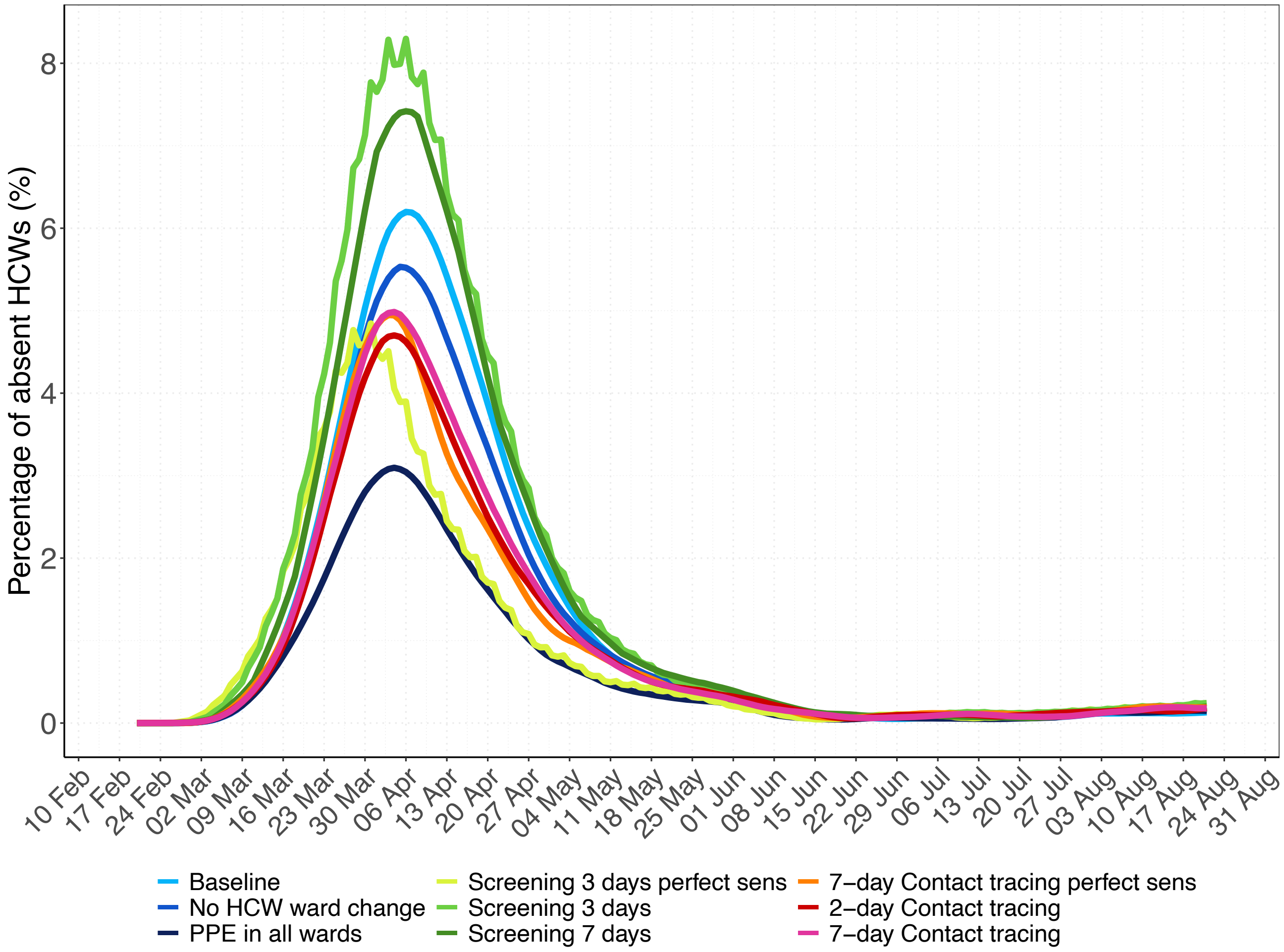

**Appendix Figure 15. Effective reproduction numbers for the nosocomial spread of the SARS-CoV-2 variant for each simulation scenario assuming 70% effective PPE.** Results shown are based on $R_S$=1·95 and $R_A$=0·8 (reproduction numbers for the SARS-CoV-2 variant with 56% higher transmissibility with respect to the wild-type SARS-CoV-2 variant). (A) For each simulation scenario, violin and boxplots of the overall reproduction numbers (for pre-/symptomatic and asymptomatic patients and HCWs combined) are shown (over 100 simulations). (B) For each simulation scenario, violin and boxplots of the reproduction numbers for pre-/symptomatic and asymptomatic individuals are shown (over 100 simulations). Note that since HCWs are assumed to immediately self-isolate upon symptom onset, the reproduction number is assigned to the pre-symptomatic state. The horizontal dashed line represents a reproduction number of 1. For screening every three days and contact tracing seven days prior to symptom onset of SARS-CoV-2 infected HCWs, we considered two different test sensitivity scenarios: time-invariant perfect test sensitivity (perfect sens) and imperfect test sensitivity varying from time since infection.

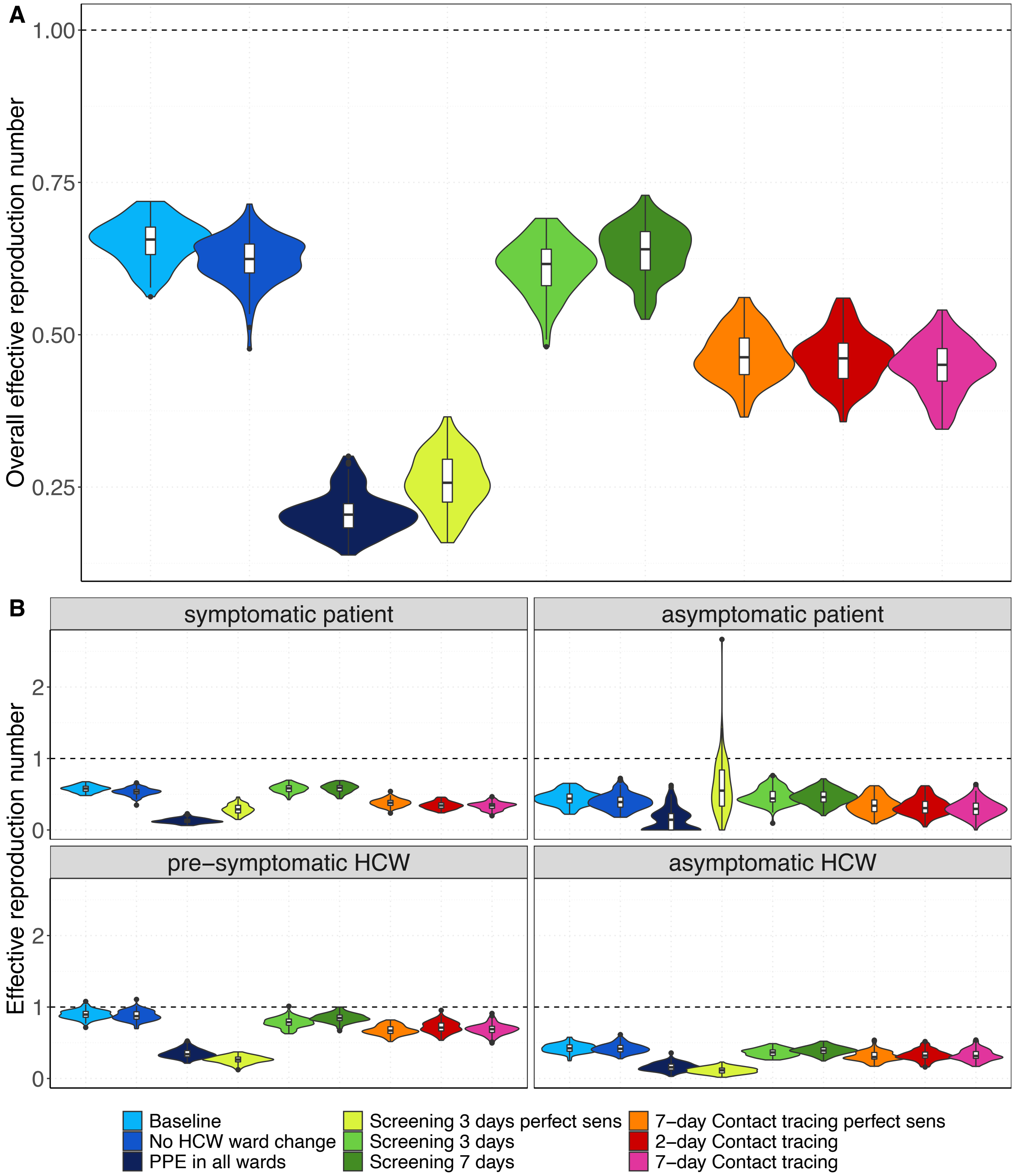

**Appendix Figure 16. Number of nosocomial transmissions of the SARS-CoV-2 variant for each simulation scenario assuming 70% effective PPE.** Results shown are based on $R_S$=1·95 and $R_A$=0·8 (reproduction numbers for the SARS-CoV-2 variant with 56% higher transmissibility with respect to the wild-type SARS-CoV-2 variant). The full rectangular bar height represents the mean total number of nosocomial transmissions during the whole study period (over 100 simulation runs). The grey error bars represent the corresponding 95% uncertainty intervals. Patients that acquire a SARS-CoV-2 nosocomial infection may be diagnosed in the hospital (due to symptom onset during hospital stay or due to detection by an intervention) or discharged to the community in a pre-symptomatic or asymptomatic state. The rectangular bars with the black border represent the mean number of individuals (patients and HCWs) infected with SARS-CoV-2 and diagnosed in the hospital. The lighter rectangular bars represent the remaining mean number of patients discharged to community undiagnosed. For screening every 3 days and 7-day contact tracing, we considered two different test sensitivity scenarios: time-invariant perfect test sensitivity (perfect sens) and time-varying imperfect test sensitivity.

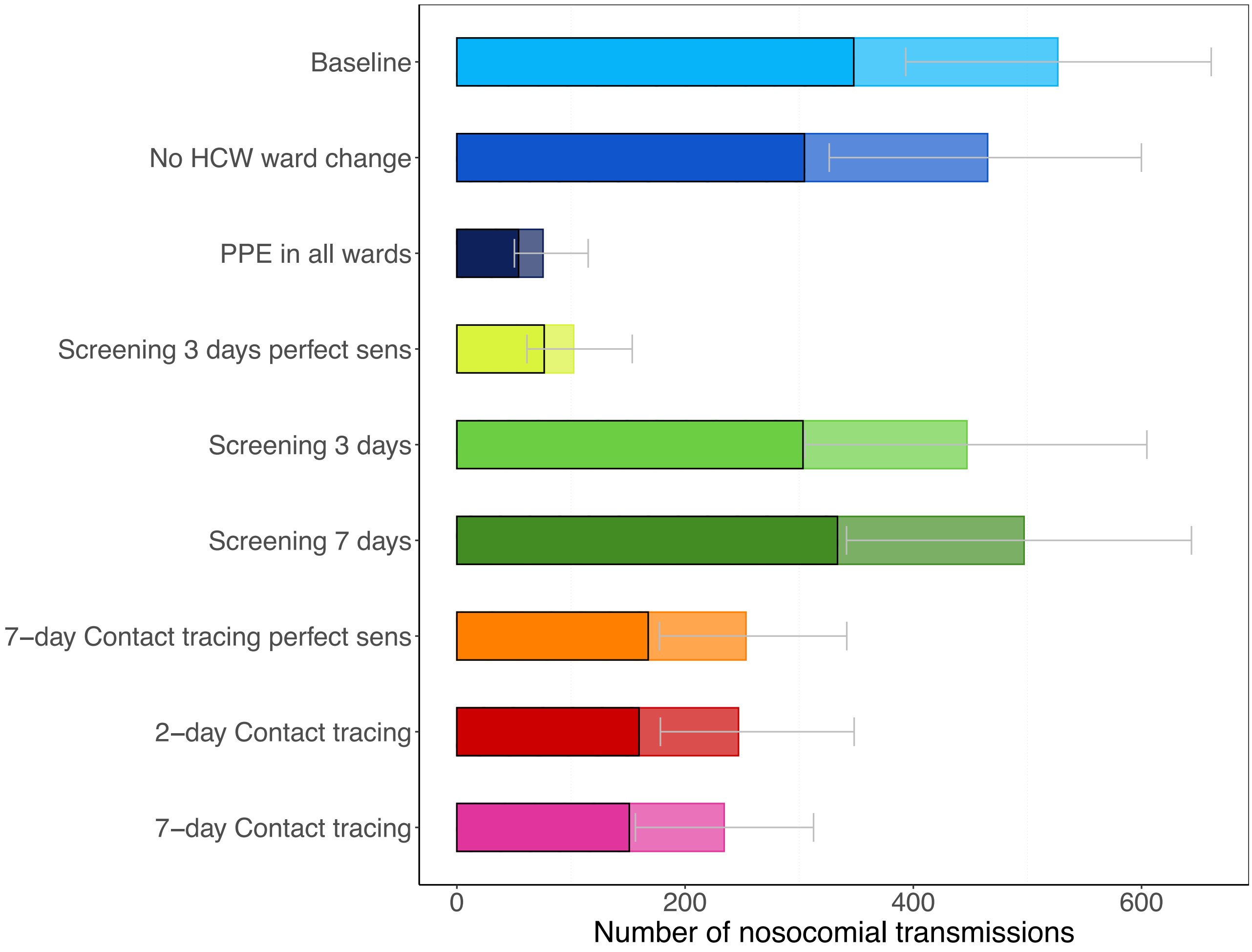

**Appendix Figure 17. Daily percentage of absent HCWs during the hospital epidemic for each simulation scenario assuming 70% effective PPE.** Results shown are based on R_S=1·95 and R_A=0·8 (reproduction numbers for the SARS-CoV-2 variant with 56% higher transmissibility with respect to the wild-type SARS-CoV-2 variant). The 7-day moving average of the mean percentage (over 100 simulation runs) of HCWs absent from work due to symptom onset or a detected SARS-CoV-2 infection screening or contact tracing is shown. For screening every 3 days and contact tracing 7 days prior to symptom onset of SARS-CoV-2 infected HCWs, we considered two different test sensitivity scenarios: time-invariant perfect test sensitivity (perfect sens) and time-varying imperfect test sensitivity.

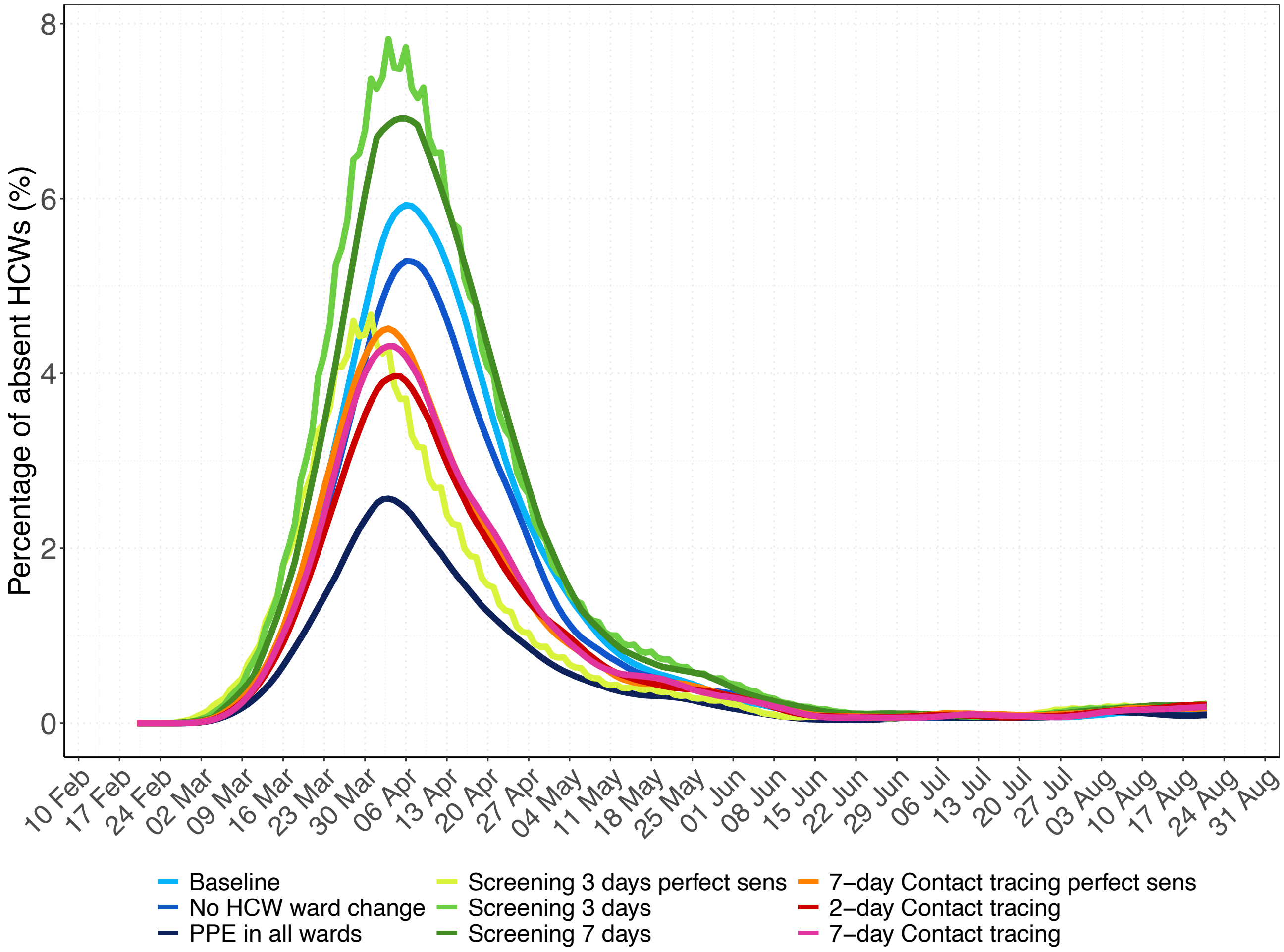

**Appendix Figure 18. Effective reproduction numbers for the nosocomial spread of the SARS-CoV-2 variant for each simulation scenario assuming equal reproduction numbers for symptomatically and asymptomatically infected individuals $R_S$=1·95 and $R_A$=1·95.** (A) For each simulation scenario, violin and boxplots of the overall reproduction numbers (for pre-/symptomatic and asymptomatic patients and HCWs combined) are shown (over 100 simulations). (B) For each simulation scenario, violin and boxplots of the reproduction numbers for pre-/symptomatic and asymptomatic individuals are shown (over 100 simulations). Note that since HCWs are assumed to immediately self-isolate upon symptom onset, the reproduction number is assigned to the pre-symptomatic state. The horizontal dashed line represents a reproduction number of 1. For screening every three days and contact tracing seven days prior to symptom onset of SARS-CoV-2 infected HCWs, we considered two different test sensitivity scenarios: time-invariant perfect test sensitivity (perfect sens) and imperfect test sensitivity varying from time since infection.

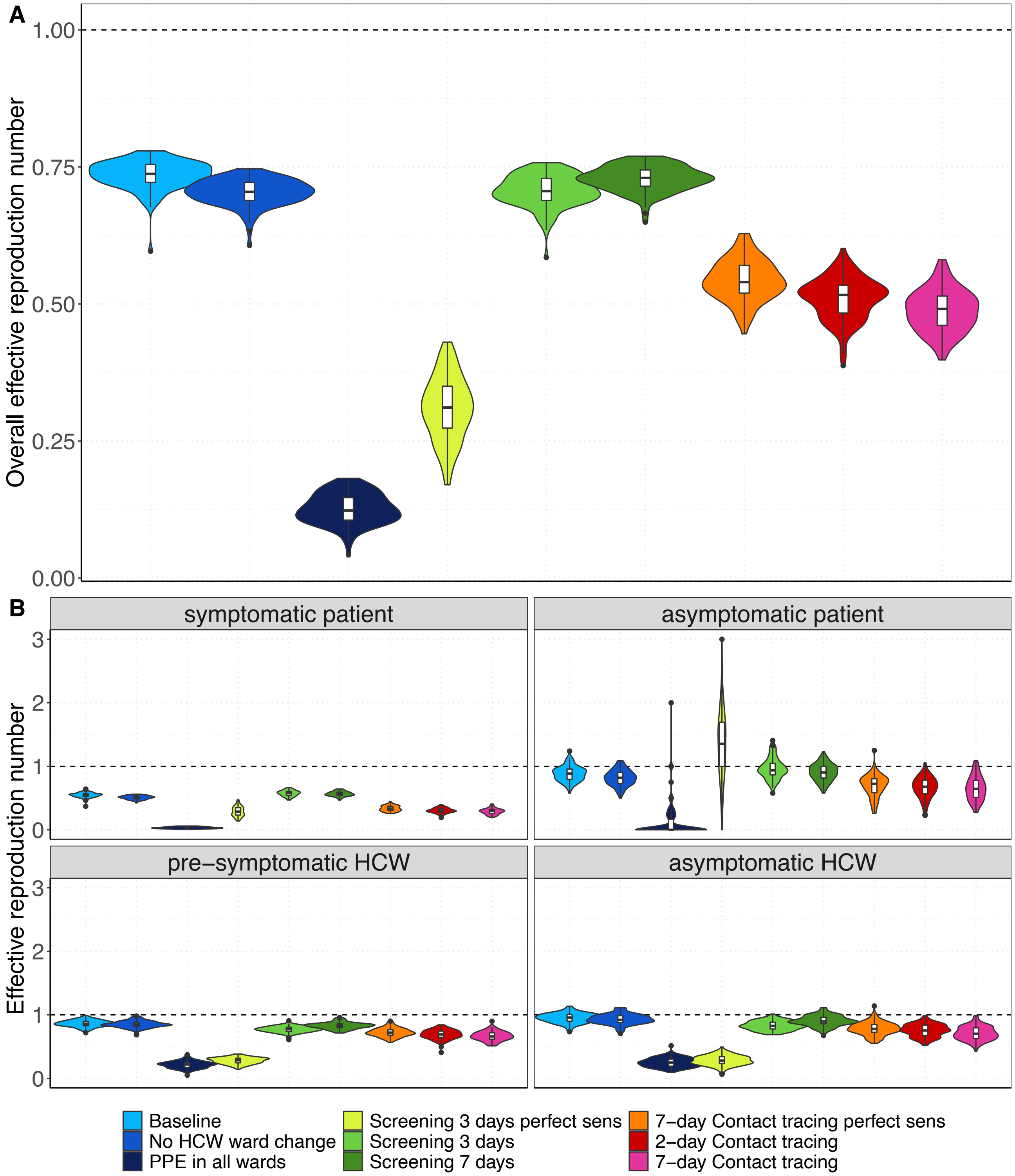

**Appendix Figure 19. Number of nosocomial transmissions of the SARS-CoV-2 variant for each simulation scenario assuming equal reproduction numbers for symptomatically and asymptomatically infected individuals $R_S$=1·95 and $R_A$=1·95.** The full rectangular bar height represents the mean total number of nosocomial transmissions during the whole study period (over 100 simulation runs). The grey error bars represent the corresponding 95% uncertainty intervals. Patients that acquire a SARS-CoV-2 nosocomial infection may be diagnosed in the hospital (due to symptom onset during hospital stay or due to detection by an intervention) or discharged to the community in a pre-symptomatic or asymptomatic state. The rectangular bars with the black border represent the mean number of individuals (patients and HCWs) infected with SARS-CoV-2 and diagnosed in the hospital. The lighter rectangular bars represent the remaining mean number of patients discharged to community undiagnosed. For screening every 3 days and 7-day contact tracing, we considered two different test sensitivity scenarios: time-invariant perfect test sensitivity (perfect sens) and time-varying imperfect test sensitivity.

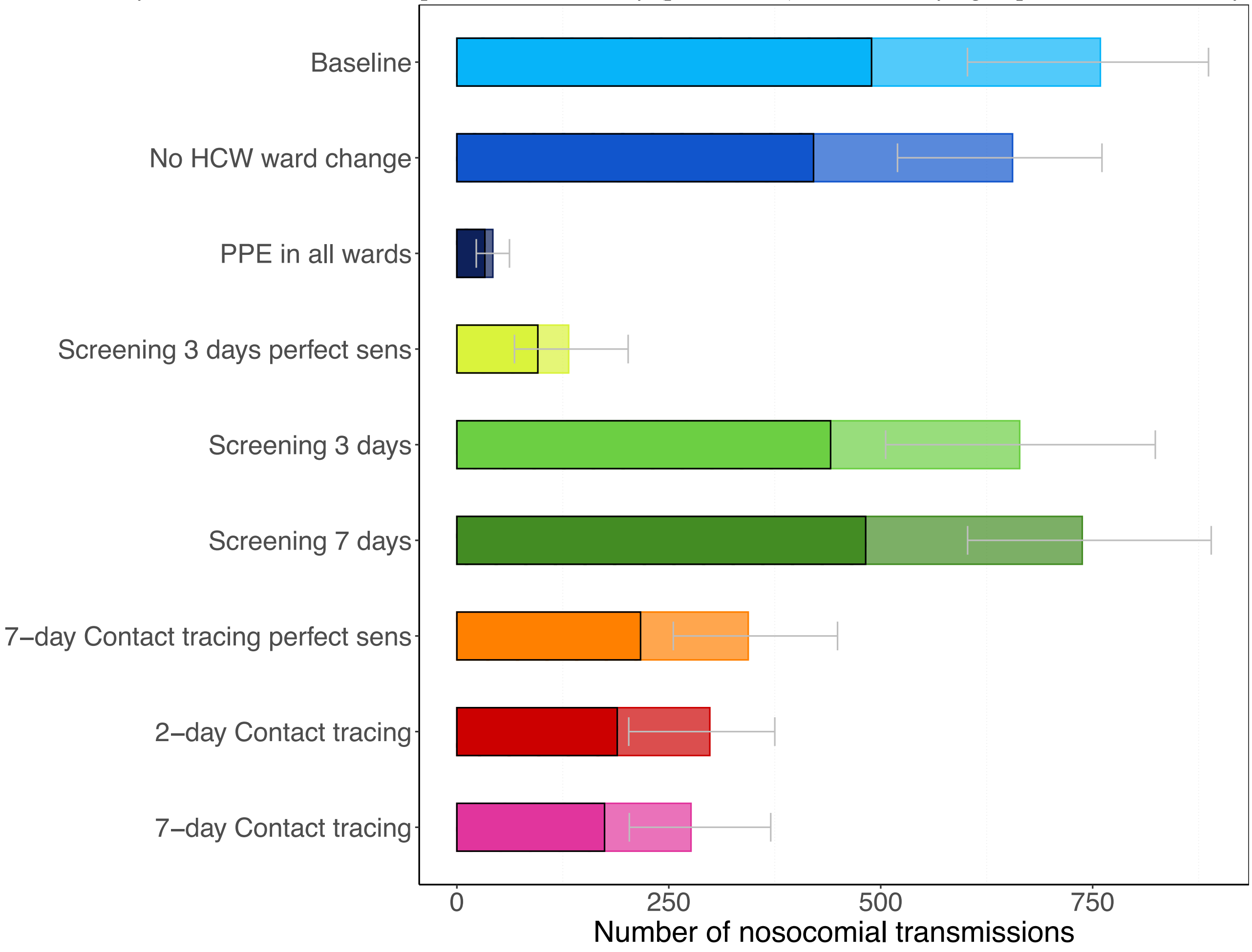

**Appendix Figure 20. Daily percentage of absent HCWs during the hospital epidemic for each simulation scenario assuming equal reproduction numbers for symptomatic and asymptomatic individuals $R_S$=1·95 and $R_A$=1·95.** The 7-day moving average of the mean percentage (over 100 simulation runs) of HCWs absent from work due to symptom onset or a detected SARS-CoV-2 infection screening or contact tracing is shown. For screening every 3 days and contact tracing 7 days prior to symptom onset of SARS-CoV-2 infected HCWs, we considered two different test sensitivity scenarios: time-invariant perfect test sensitivity (perfect sens) and imperfect test sensitivity from time since infection.

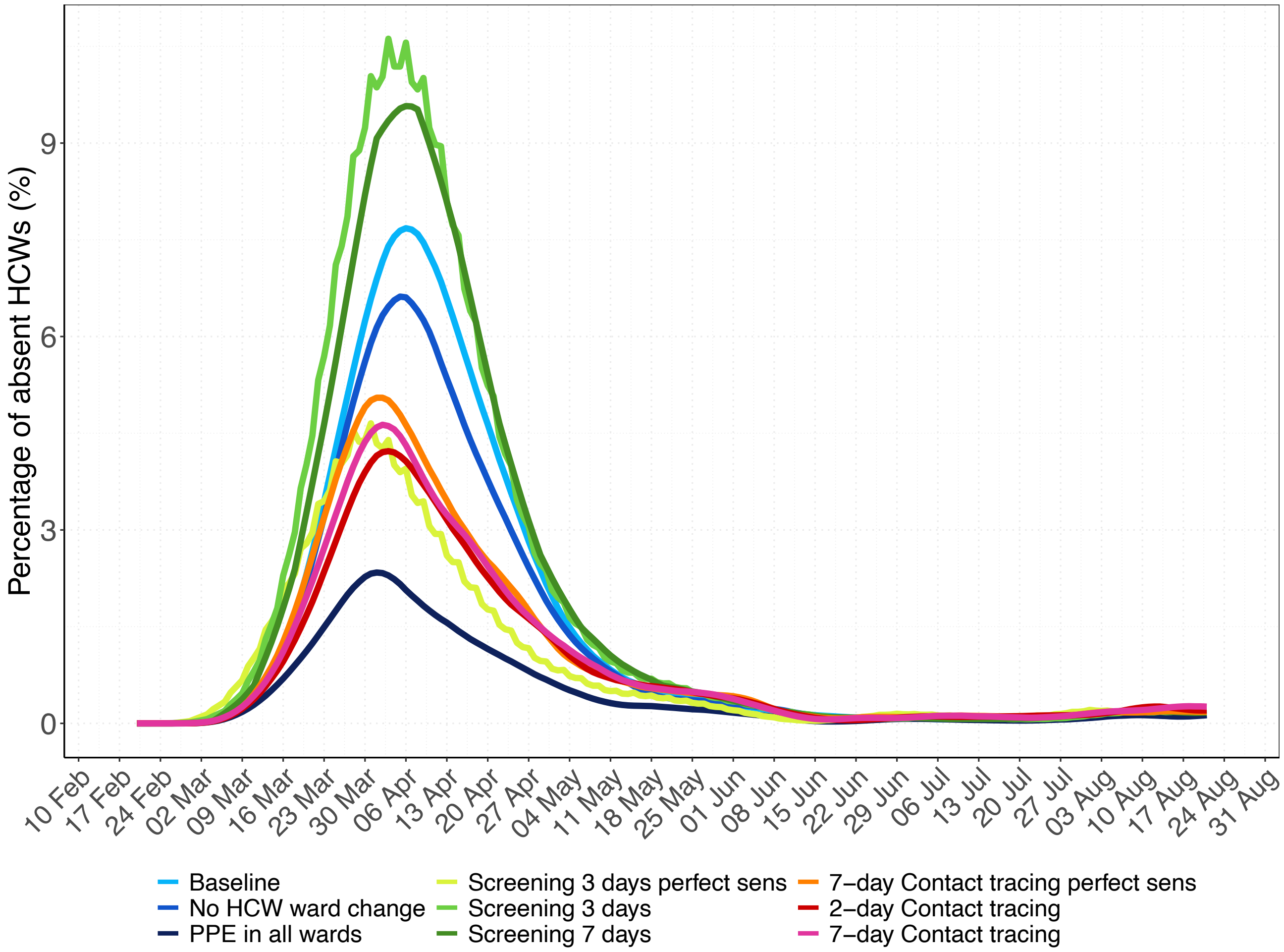

**Appendix Figure 21. Effective reproduction numbers for each simulation scenario assuming reproduction numbers of the wild-type SARS-CoV-2 variant.** Results shown are based on $R_S$=1·25 and $R_A$=0·5. (A) For each simulation scenario, violin and boxplots of the overall reproduction numbers (for pre-/symptomatic and asymptomatic patients and HCWs combined) are shown (over 100 simulations). (B) For each simulation scenario, violin and boxplots of the reproduction numbers for pre-/symptomatic and asymptomatic individuals are shown (over 100 simulations). Note that since HCWs are assumed to immediately self-isolate upon symptom onset, the reproduction number is assigned to the pre-symptomatic state. The horizontal dashed line represents a reproduction number of 1. For screening every three days and contact tracing seven days prior to symptom onset of SARS-CoV-2 infected HCWs, we considered two different test sensitivity scenarios: time-invariant perfect test sensitivity (perfect sens) and imperfect test sensitivity varying from time since infection.

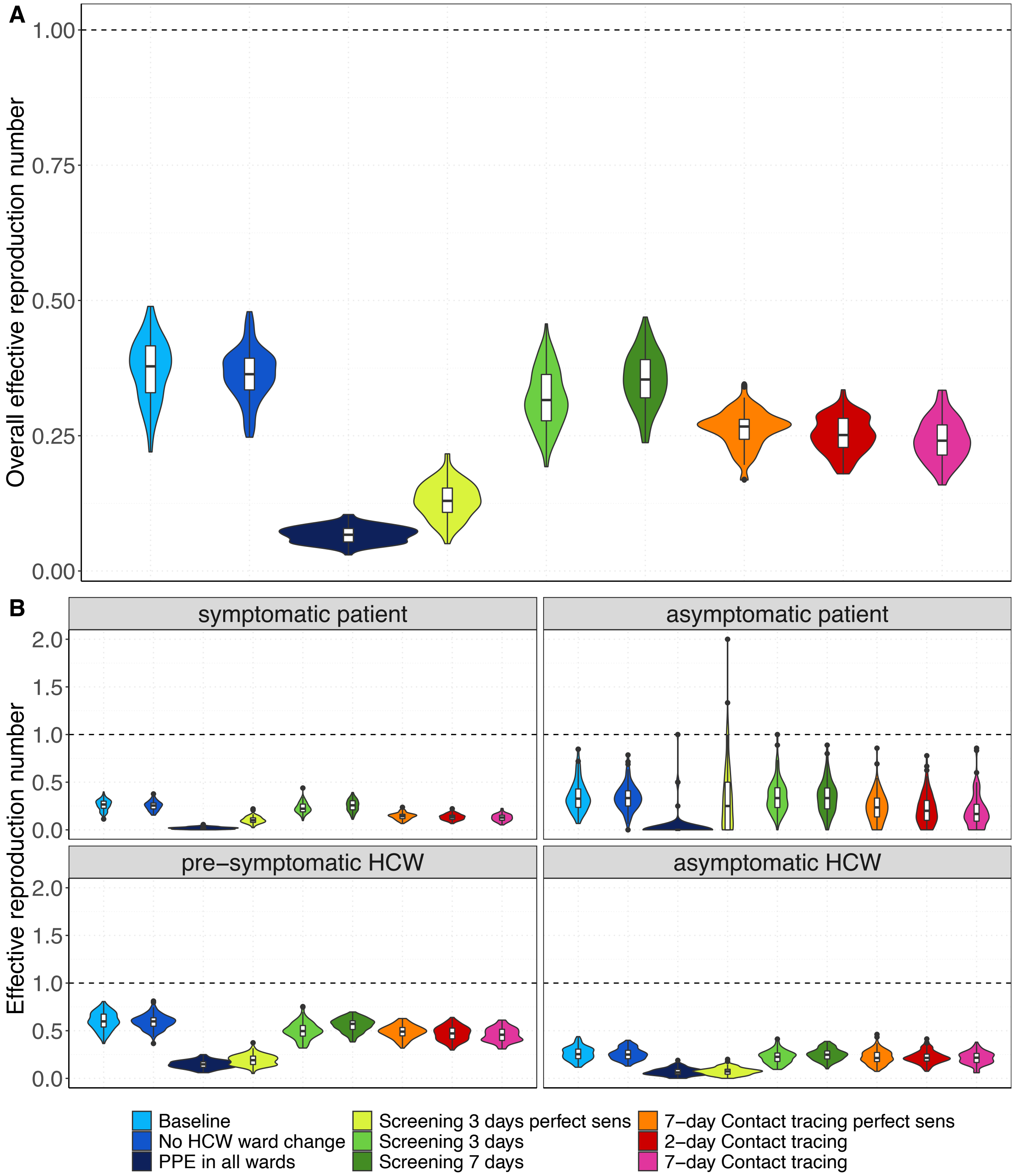

**Appendix Figure 22. Number of nosocomial transmissions of the SARS-CoV-2 variant for each simulation scenario assuming reproduction numbers of the wild-type SARS-CoV-2 variant.** Results shown are based on $R_S$=1·25 and $R_A$=0·5. The full rectangular bar height represents the mean total number of nosocomial transmissions during the whole study period (over 100 simulation runs). The grey error bars represent the corresponding 95% uncertainty intervals. Patients that acquire a SARS-CoV-2 nosocomial infection may be diagnosed in the hospital (due to symptom onset during hospital stay or due to detection by an intervention) or discharged to the community in a pre-symptomatic or asymptomatic state. The rectangular bars with the black border represent the mean number of individuals (patients and HCWs) infected with SARS-CoV-2 and diagnosed in the hospital. The lighter rectangular bars represent the remaining mean number of patients discharged to community undiagnosed. For screening every 3 days and 7-day contact tracing, we considered two different test sensitivity scenarios: time-invariant perfect test sensitivity (perfect sens) and time-varying imperfect test sensitivity.

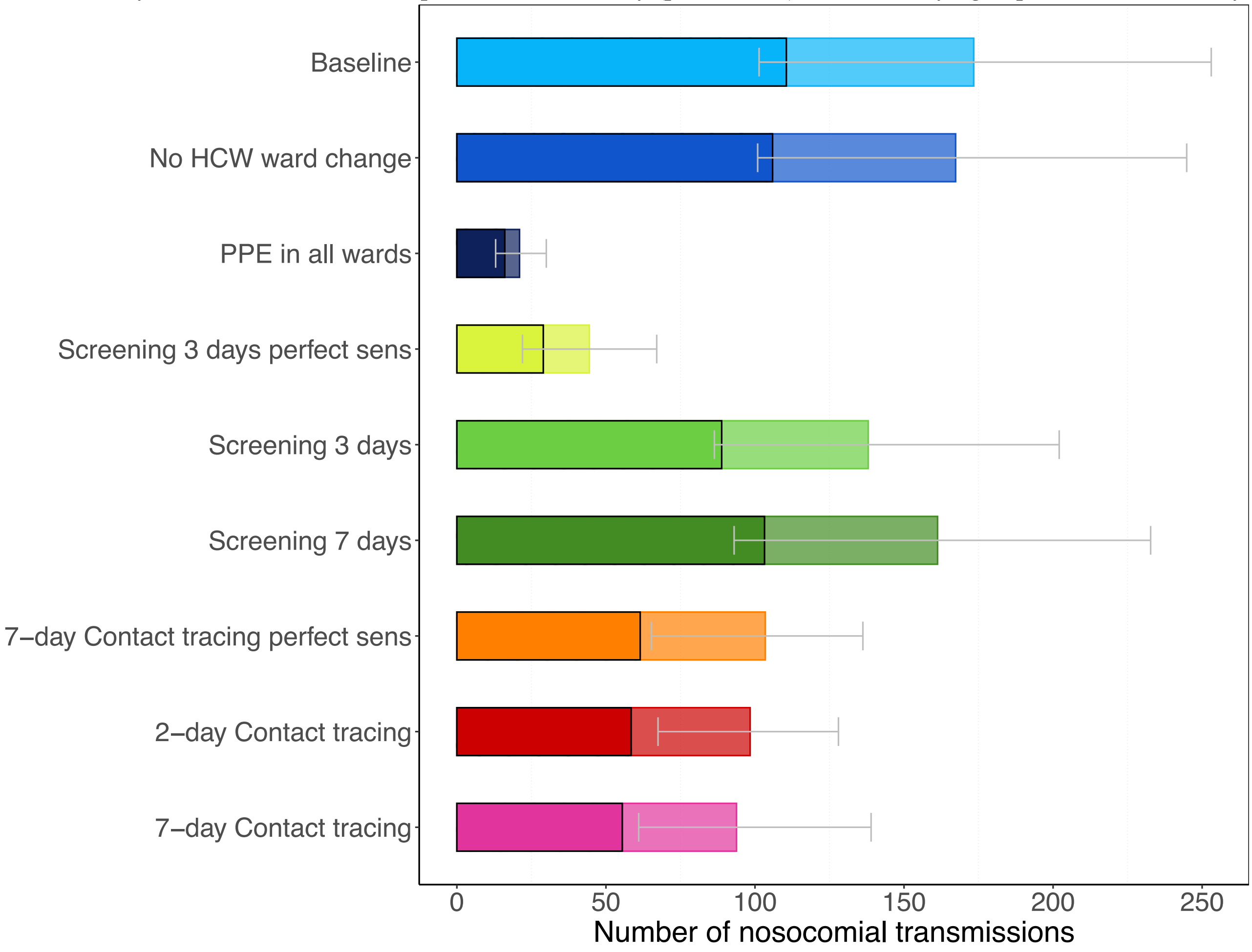

**Appendix Figure 23. Daily percentage of absent HCWs during the hospital epidemic for each simulation scenario assuming reproduction numbers of the wild-type SARS-CoV-2 variant.** Results shown are based on $R_S$=1·25 and $R_A$=0·5. The 7-day moving average of the mean percentage (over 100 simulation runs) of HCWs absent from work due to symptom onset or a detected SARS-CoV-2 infection screening or contact tracing is shown. For screening every 3 days and contact tracing 7 days prior to symptom onset of SARS-CoV-2 infected HCWs, we considered two different test sensitivity scenarios: time-invariant perfect test sensitivity (perfect sens) and imperfect test sensitivity from time since infection.

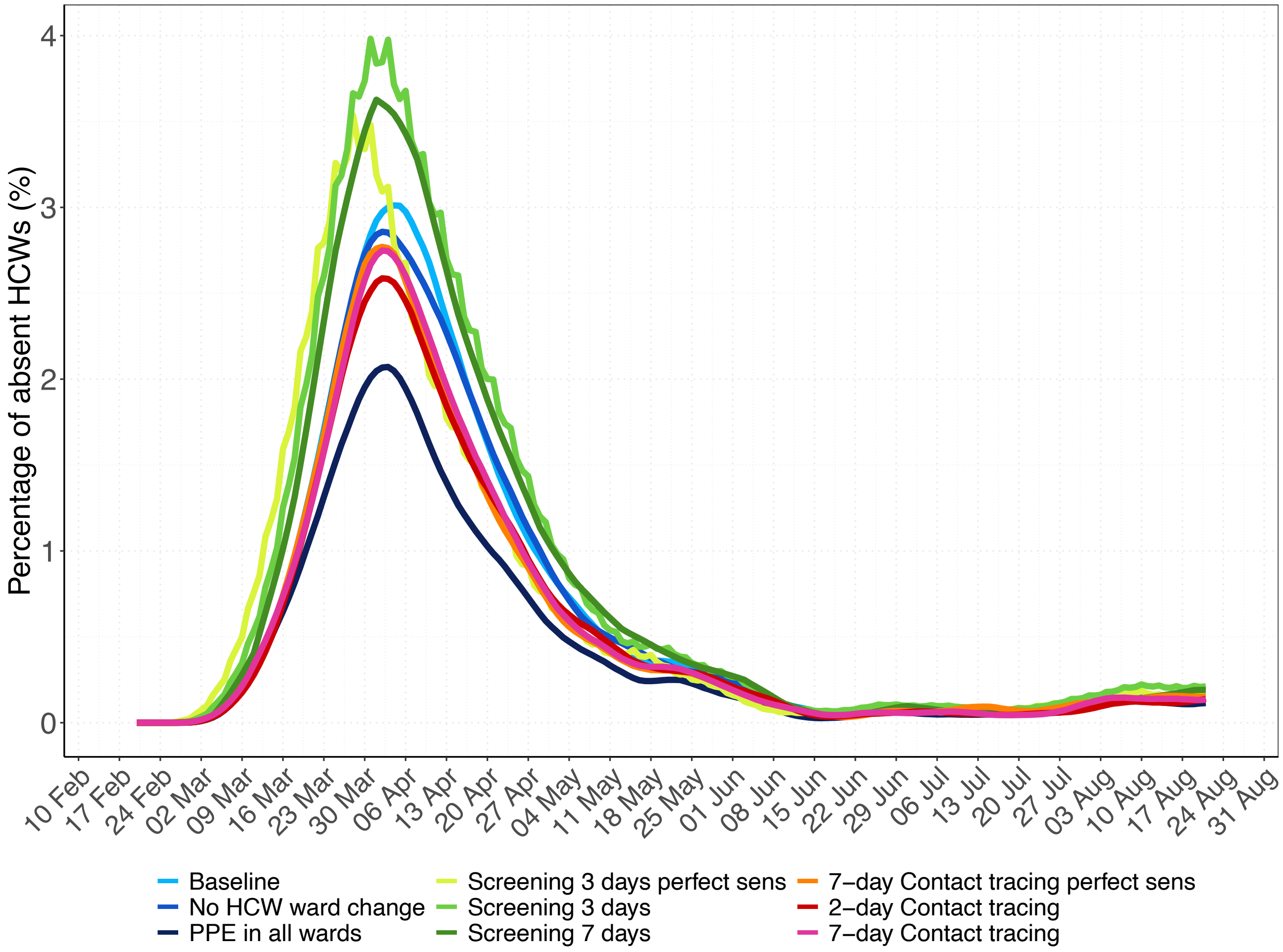

**Appendix Figure 24. Effective reproduction numbers for the nosocomial spread of the SARS-CoV-2 variant for each simulation scenario assuming higher contact rates between HCWs.** Results shown are based on $R_S$=1·95 and $R_A$=0·8 (reproduction numbers for the SARS-CoV-2 variant with 56% higher transmissibility with respect to the wild-type SARS-CoV-2 variant). (A) For each simulation scenario, violin and boxplots of the overall reproduction numbers (for pre-/symptomatic and asymptomatic patients and HCWs combined) are shown (over 100 simulations). (B) For each simulation scenario, violin and boxplots of the reproduction numbers for pre-/symptomatic and asymptomatic individuals are shown (over 100 simulations). Note that since HCWs are assumed to immediately self-isolate upon symptom onset, the reproduction number is assigned to the pre-symptomatic state. The horizontal dashed line represents a reproduction number of 1. For screening every three days and contact tracing seven days prior to symptom onset of SARS-CoV-2 infected HCWs, we considered two different test sensitivity scenarios: time-invariant perfect test sensitivity (perfect sens) and imperfect test sensitivity varying from time since infection.

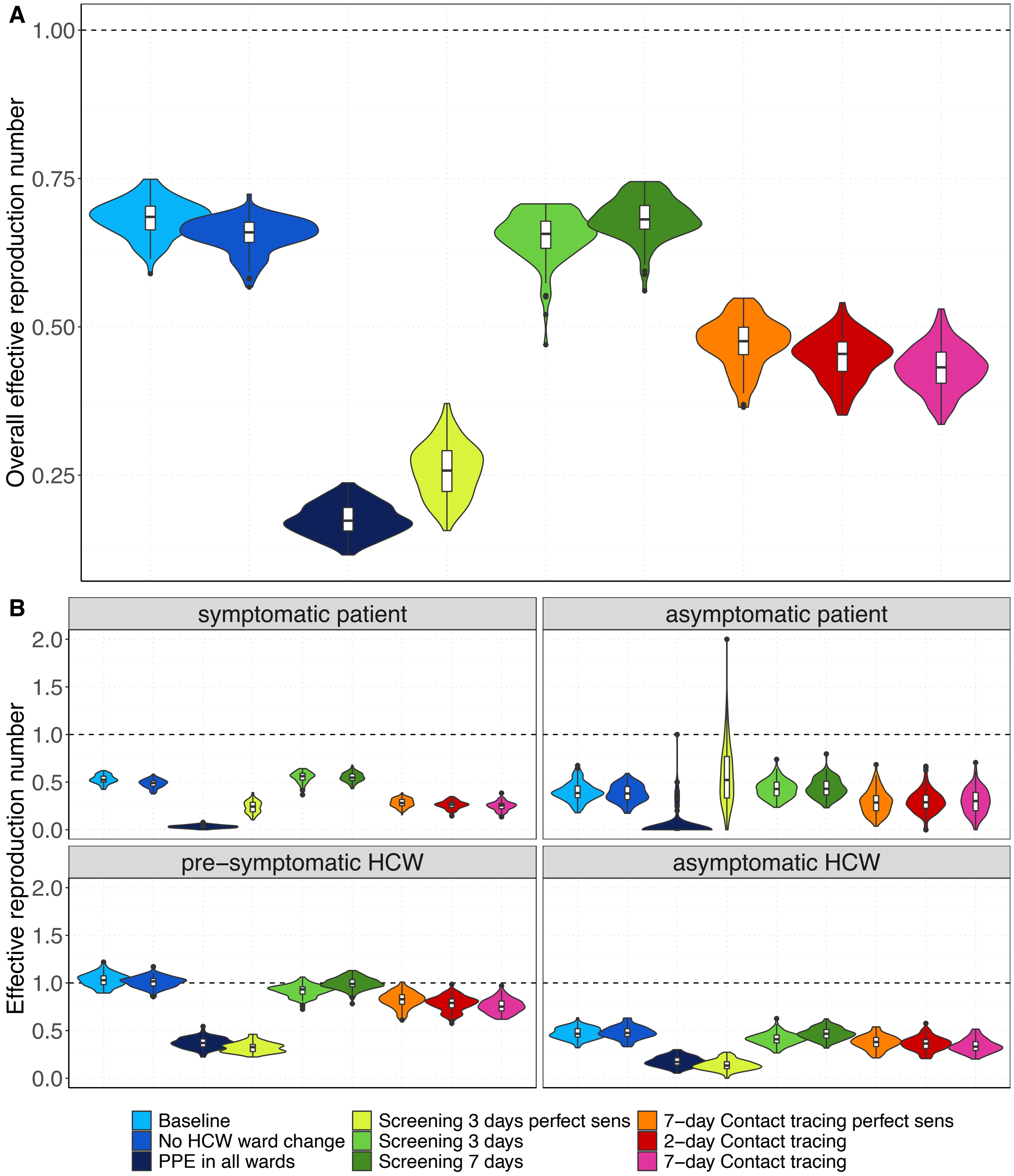

**Appendix Figure 25. Number of nosocomial transmissions of the SARS-CoV-2 variant for each simulation scenario assuming higher contact rates between HCWs.** Results shown are based on $R_S$=1·95 and $R_A$=0·8 (reproduction numbers for the SARS-CoV-2 variant with 56% higher transmissibility with respect to the wild-type SARS-CoV-2 variant). The full rectangular bar height represents the mean total number of nosocomial transmissions during the whole study period (over 100 simulation runs). The grey error bars represent the corresponding 95% uncertainty intervals. Patients that acquire a SARS-CoV-2 nosocomial infection may be diagnosed in the hospital (due to symptom onset during hospital stay or due to detection by an intervention) or discharged to the community in a pre-symptomatic or asymptomatic state. The rectangular bars with the black border represent the mean number of individuals (patients and HCWs) infected with SARS-CoV-2 and diagnosed in the hospital. The lighter rectangular bars represent the remaining mean number of patients discharged to community undiagnosed. For screening every 3 days and 7-day contact tracing, we considered two different test sensitivity scenarios: time-invariant perfect test sensitivity (perfect sens) and time-varying imperfect test sensitivity.

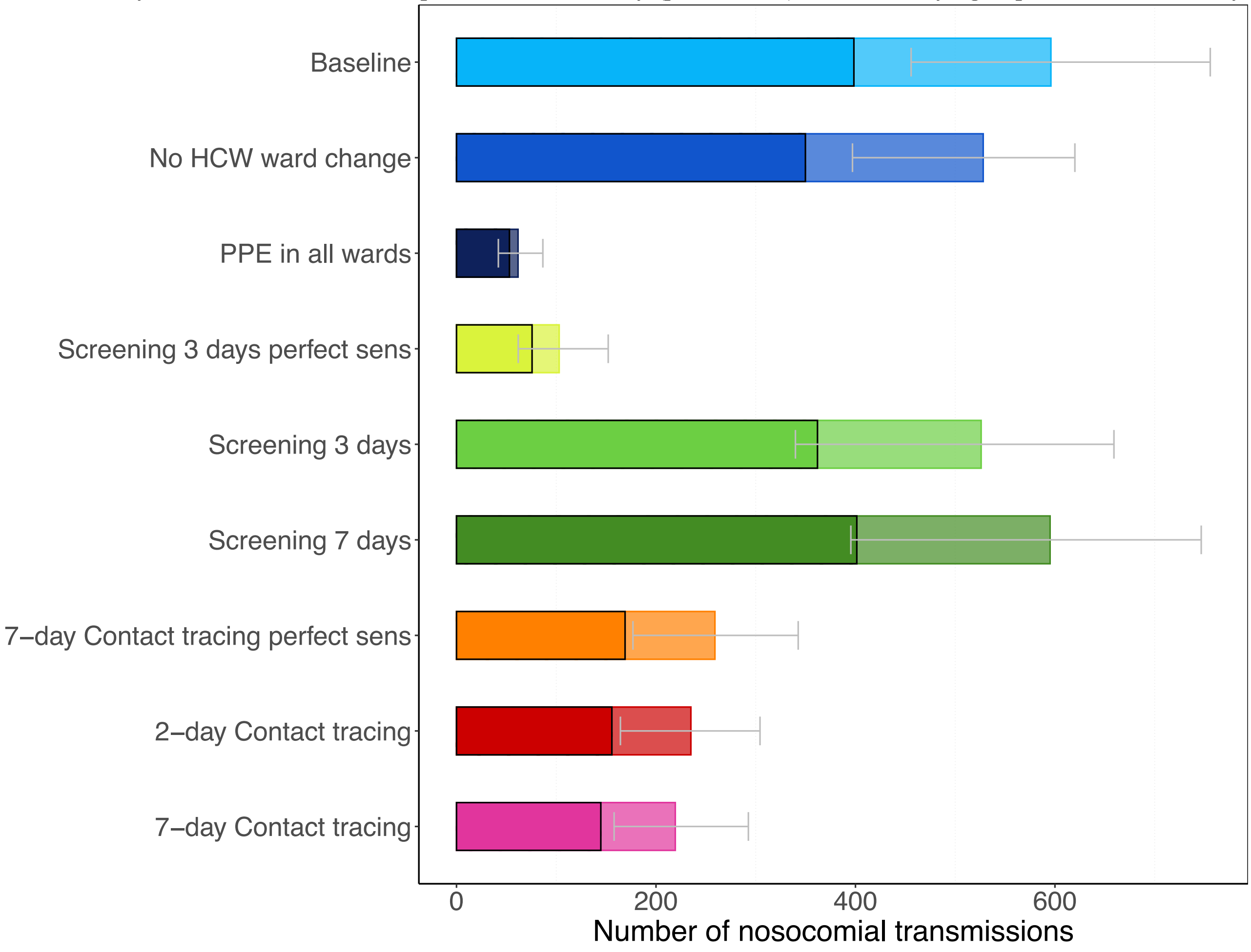

**Appendix Figure 26. Daily percentage of absent HCWs during the hospital epidemic for each simulation scenario assuming higher contact rates between HCWs.** Results shown are based on $R_S$=1·95 and $R_A$=0·8 (reproduction numbers for the SARS-CoV-2 variant with 56% higher transmissibility with respect to the wild-type SARS-CoV-2 variant). The 7-day moving average of the mean percentage (over 100 simulation runs) of HCWs absent from work due to symptom onset or a detected SARS-CoV-2 infection screening or contact tracing is shown. For screening every 3 days and contact tracing 7 days prior to symptom onset of SARS-CoV-2 infected HCWs, we considered two different test sensitivity scenarios: time-invariant perfect test sensitivity (perfect sens) and imperfect test sensitivity from time since infection.

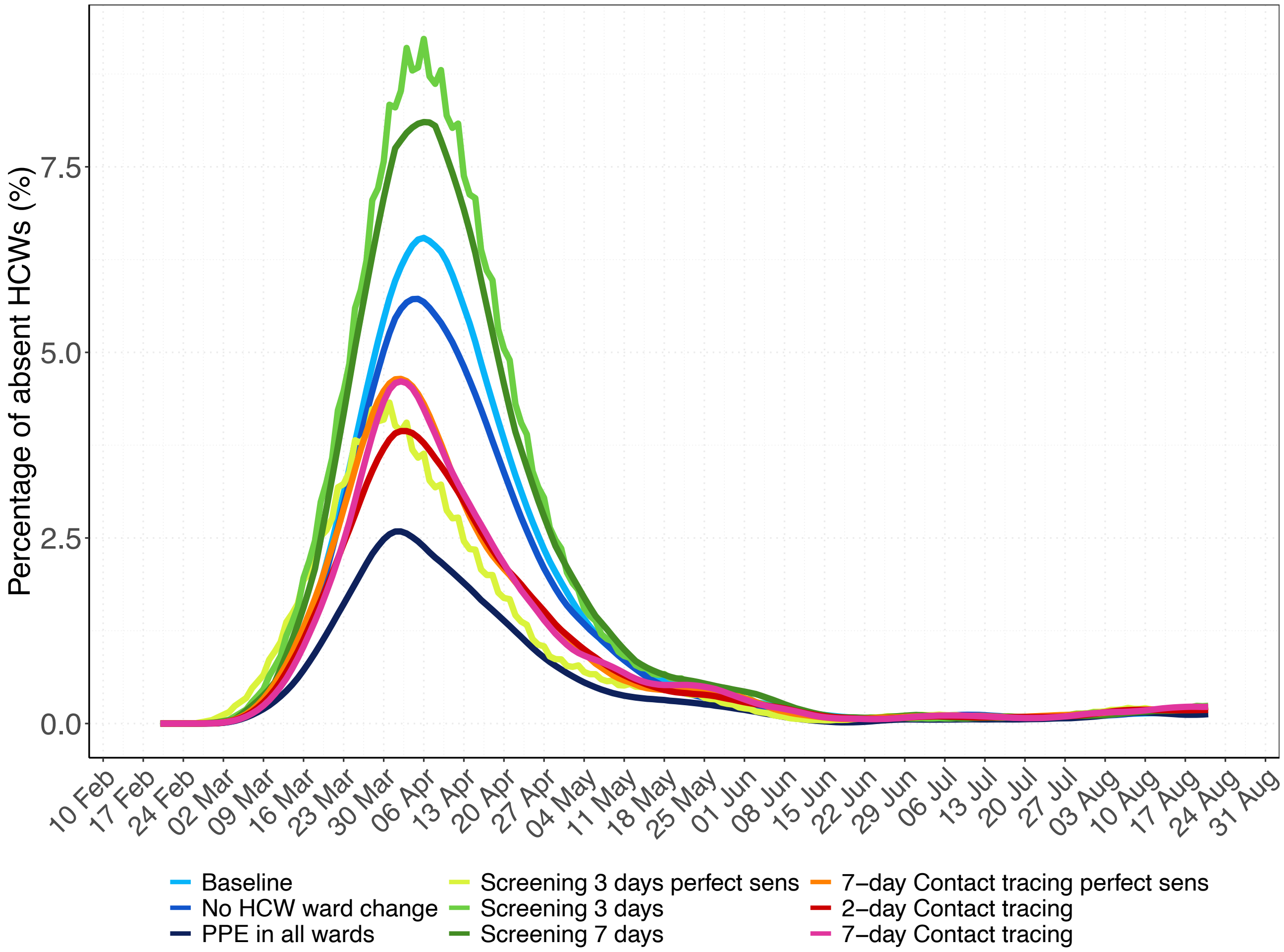

**Appendix Figure 27. Effective reproduction numbers for the nosocomial spread of the SARS-CoV-2 variant for the high test sensitivity scenario.** Results shown are based on $R_S$=1·95 and $R_A$=0·8 (reproduction numbers for the SARS-CoV-2 variant with 56% higher transmissibility with respect to the wild-type SARS-CoV-2 variant). (A) For each simulation scenario, violin and boxplots of the overall reproduction numbers (for pre-/symptomatic and asymptomatic patients and HCWs combined) are shown (over 100 simulations). (B) For each simulation scenario, violin and boxplots of the reproduction numbers for pre-/symptomatic and asymptomatic individuals are shown (over 100 simulations). Note that since HCWs are assumed to immediately self-isolate upon symptom onset, the reproduction number is assigned to the pre-symptomatic state. The horizontal dashed line represents a reproduction number of 1. For screening every three days and contact tracing seven days prior to symptom onset of SARS-CoV-2 infected HCWs, we considered two different test sensitivity scenarios: time-invariant perfect test sensitivity (perfect sens) and imperfect test sensitivity varying from time since infection.

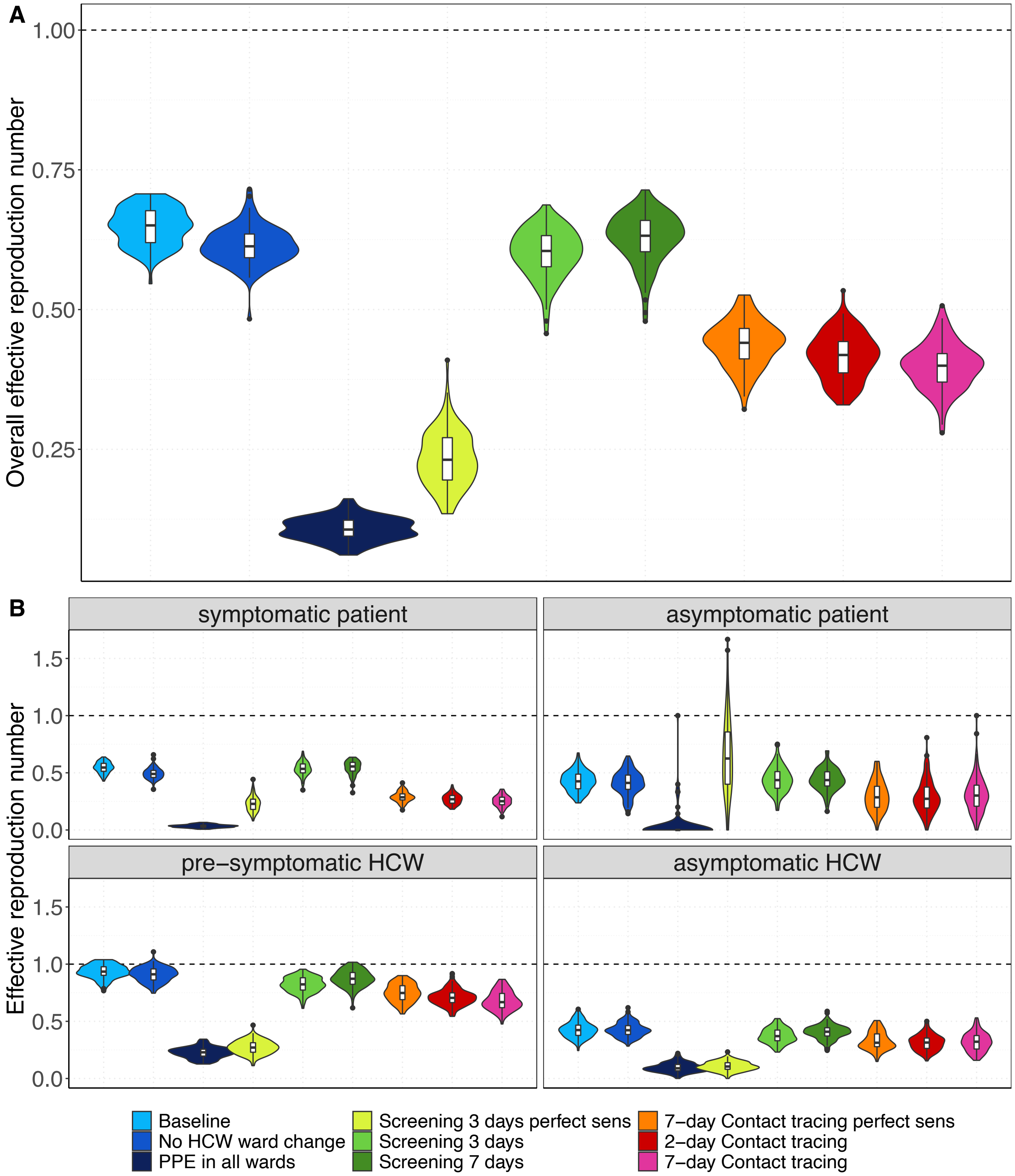

**Appendix Figure 28. Number of nosocomial transmissions of the SARS-CoV-2 variant for each simulation scenario for the high test sensitivity scenario.** Results shown are based on $R_S$=1·95 and $R_A$=0·8 (reproduction numbers for the SARS-CoV-2 variant with 56% higher transmissibility with respect to the wild-type SARS-CoV-2 variant). The full rectangular bar height represents the mean total number of nosocomial transmissions during the whole study period (over 100 simulation runs). The grey error bars represent the corresponding 95% uncertainty intervals. Patients that acquire a SARS-CoV-2 nosocomial infection may be diagnosed in the hospital (due to symptom onset during hospital stay or due to detection by an intervention) or discharged to the community in a pre-symptomatic or asymptomatic state. The rectangular bars with the black border represent the mean number of individuals (patients and HCWs) infected with SARS-CoV-2 and diagnosed in the hospital. The lighter rectangular bars represent the remaining mean number of patients discharged to community undiagnosed. For screening every 3 days and 7-day contact tracing, we considered two different test sensitivity scenarios: time-invariant perfect test sensitivity (perfect sens) and time-varying imperfect test sensitivity.

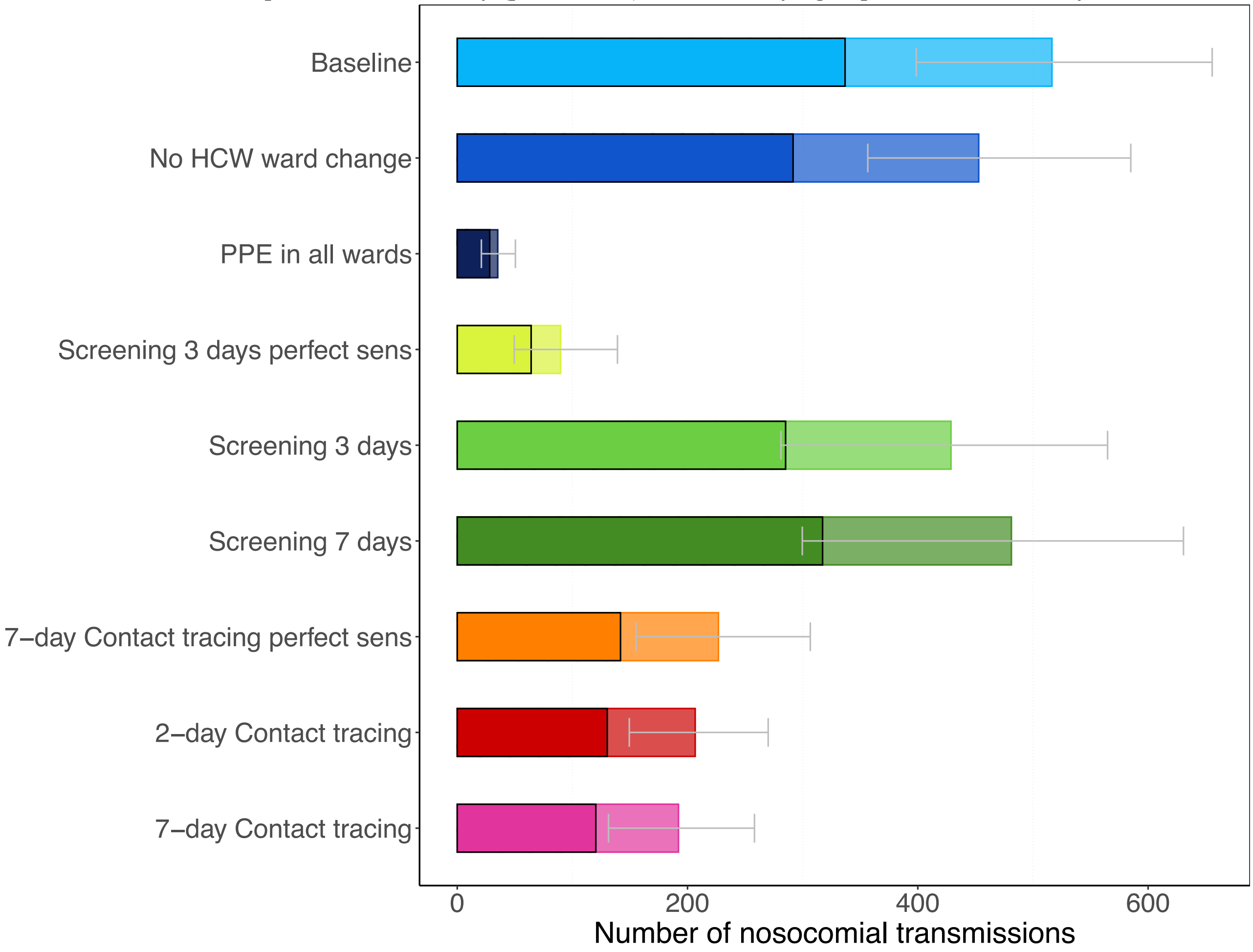

**Appendix Figure 29. Daily percentage of absent HCWs during the hospital epidemic for each simulation scenario for the high test sensitivity scenario.** Results shown are based on $R_S$=1·95 and $R_A$=0·8 (reproduction numbers for the SARS-CoV-2 variant with 56% higher transmissibility with respect to the wild-type SARS-CoV-2 variant). The 7-day moving average of the mean percentage (over 100 simulation runs) of HCWs absent from work due to symptom onset or a detected SARS-CoV-2 infection screening or contact tracing is shown. For screening every 3 days and contact tracing 7 days prior to symptom onset of SARS-CoV-2 infected HCWs, we considered two different test sensitivity scenarios: time-invariant perfect test sensitivity (perfect sens) and imperfect test sensitivity from time since infection.

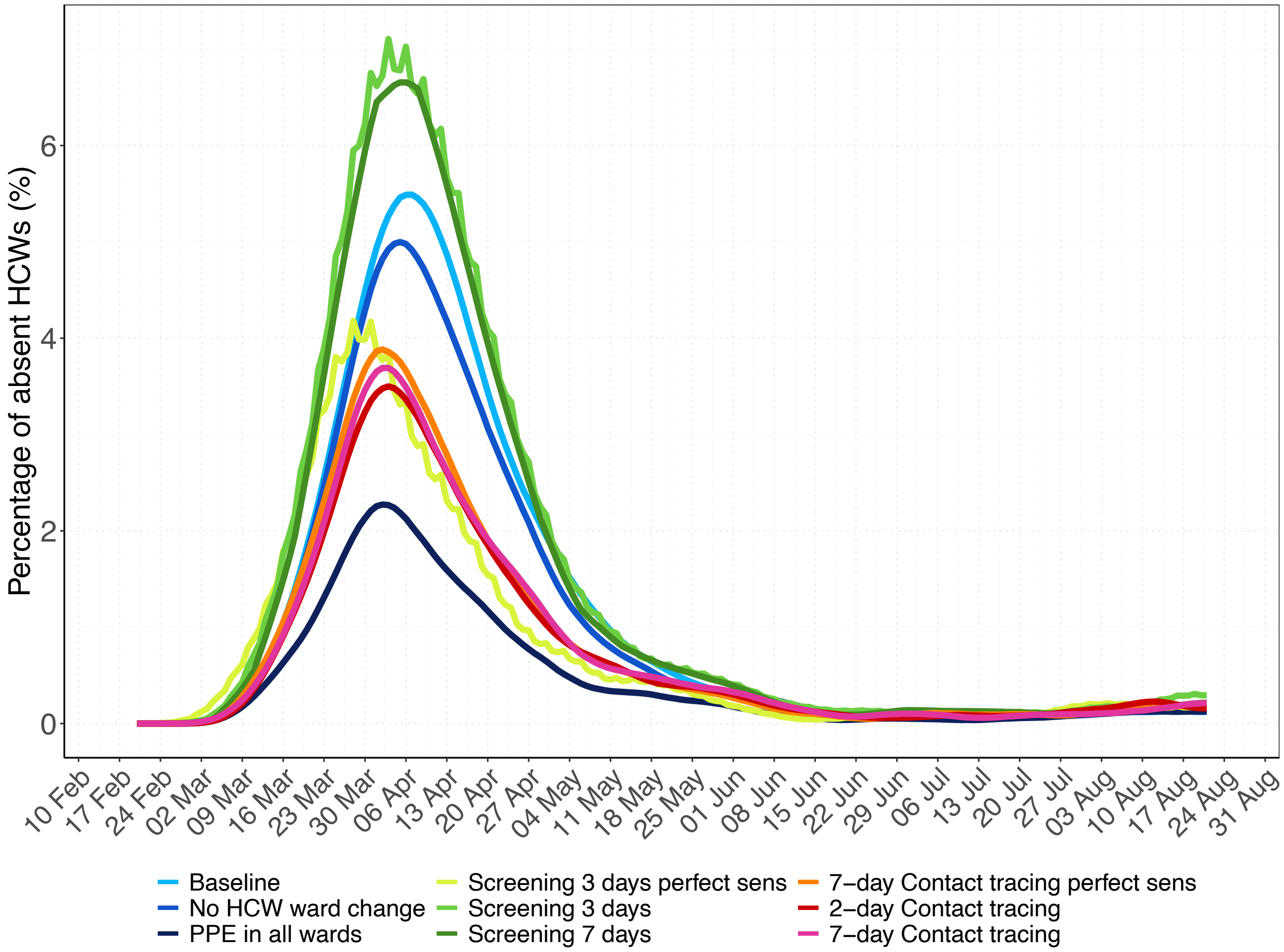

**Appendix Figure 30. Effective reproduction numbers for the nosocomial spread of the SARS-CoV-2 variant for the high test sensitivity scenario.** Results shown are based on $R_S$=1·95 and $R_A$=0·8 (reproduction numbers for the SARS-CoV-2 variant with 56% higher transmissibility with respect to the wild-type SARS-CoV-2 variant). (A) For each simulation scenario, violin and boxplots of the overall reproduction numbers (for pre-/symptomatic and asymptomatic patients and HCWs combined) are shown (over 100 simulations). (B) For each simulation scenario, violin and boxplots of the reproduction numbers for pre-/symptomatic and asymptomatic individuals are shown (over 100 simulations). Note that since HCWs are assumed to immediately self-isolate upon symptom onset, the reproduction number is assigned to the pre-symptomatic state. The horizontal dashed line represents a reproduction number of 1. For screening every three days and contact tracing seven days prior to symptom onset of SARS-CoV-2 infected HCWs, we considered two different test sensitivity scenarios: time-invariant perfect test sensitivity (perfect sens) and imperfect test sensitivity varying from time since infection.

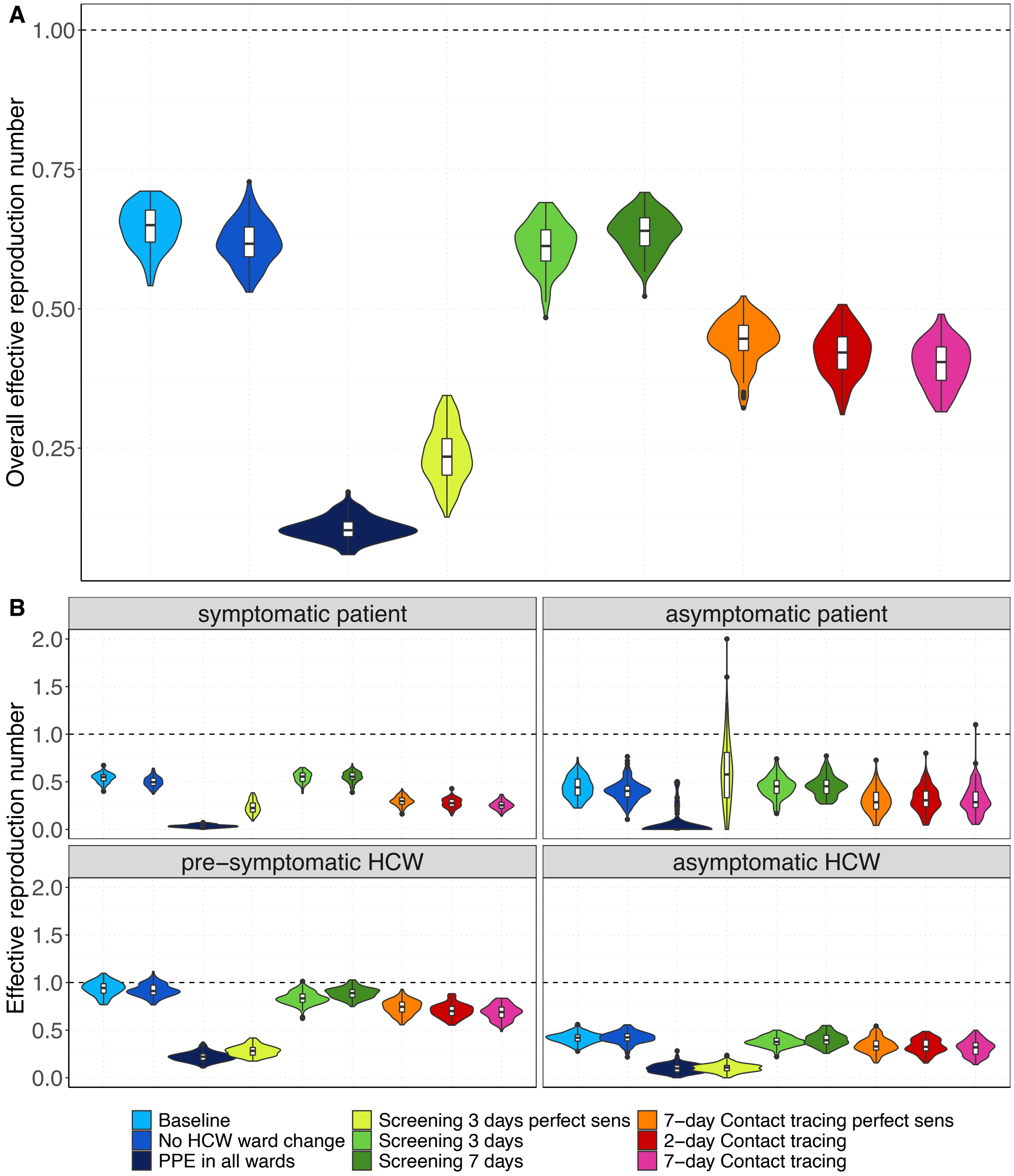

**Appendix Figure 31. Number of nosocomial transmissions of the SARS-CoV-2 variant for each simulation scenario for the low test sensitivity scenario.** Results shown are based on $R_S$=1·95 and $R_A$=0·8 (reproduction numbers for the SARS-CoV-2 variant with 56% higher transmissibility with respect to the wild-type SARS-CoV-2 variant). The full rectangular bar height represents the mean total number of nosocomial transmissions during the whole study period (over 100 simulation runs). The grey error bars represent the corresponding 95% uncertainty intervals. Patients that acquire a SARS-CoV-2 nosocomial infection may be diagnosed in the hospital (due to symptom onset during hospital stay or due to detection by an intervention) or discharged to the community in a pre-symptomatic or asymptomatic state. The rectangular bars with the black border represent the mean number of individuals (patients and HCWs) infected with SARS-CoV-2 and diagnosed in the hospital. The lighter rectangular bars represent the remaining mean number of patients discharged to community undiagnosed. For screening every 3 days and 7-day contact tracing, we considered two different test sensitivity scenarios: time-invariant perfect test sensitivity (perfect sens) and time-varying imperfect test sensitivity.

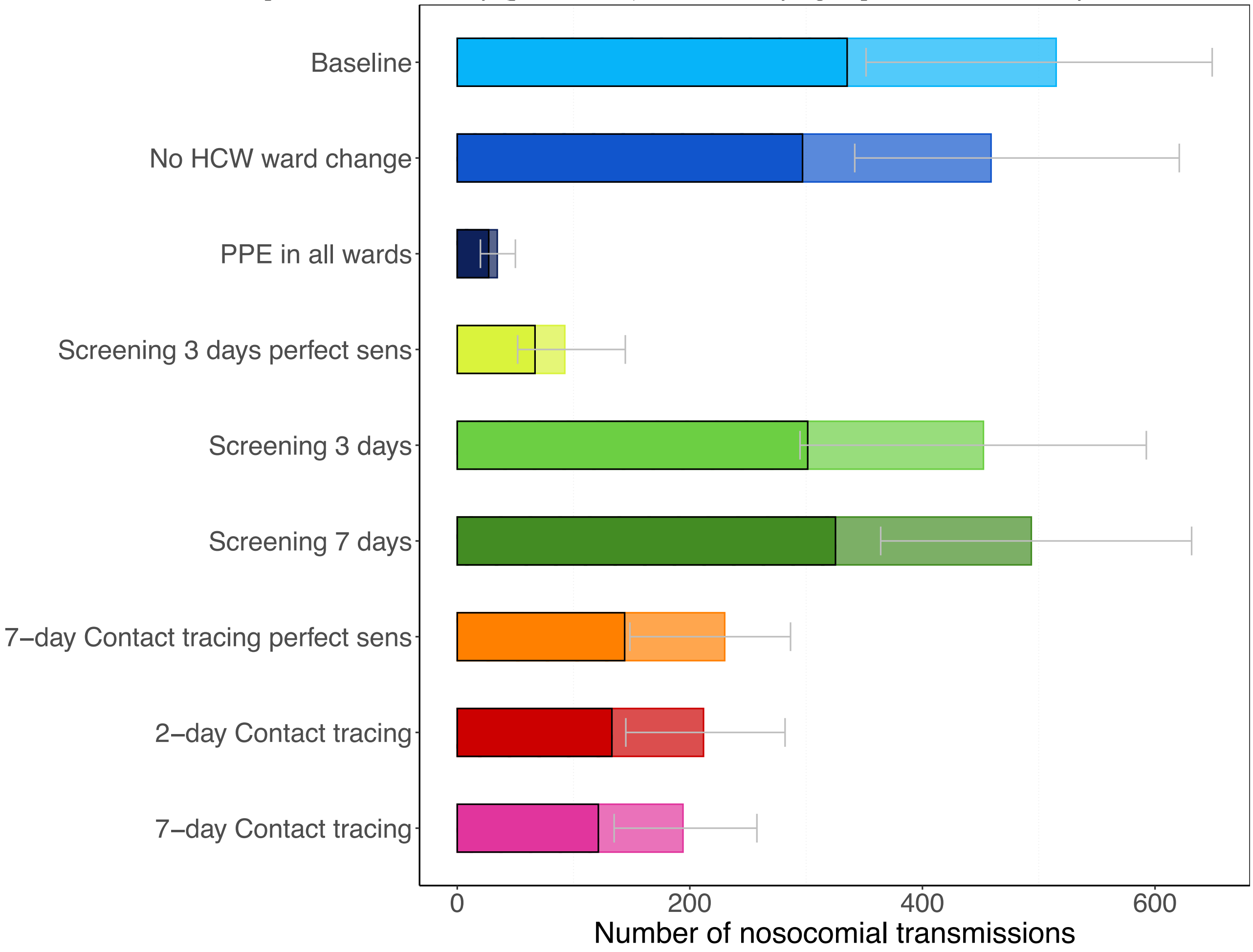

**Appendix Figure 32. Daily percentage of absent HCWs during the hospital epidemic for each simulation scenario for the low test sensitivity scenario.** Results shown are based on $R_S$=1·95 and $R_A$=0·8 (reproduction numbers for the SARS-CoV-2 variant with 56% higher transmissibility with respect to the wild-type SARS-CoV-2 variant). The 7-day moving average of the mean percentage (over 100 simulation runs) of HCWs absent from work due to symptom onset or a detected SARS-CoV-2 infection screening or contact tracing is shown. For screening every 3 days and contact tracing 7 days prior to symptom onset of SARS-CoV-2 infected HCWs, we considered two different test sensitivity scenarios: time-invariant perfect test sensitivity (perfect sens) and imperfect test sensitivity from time since infection.

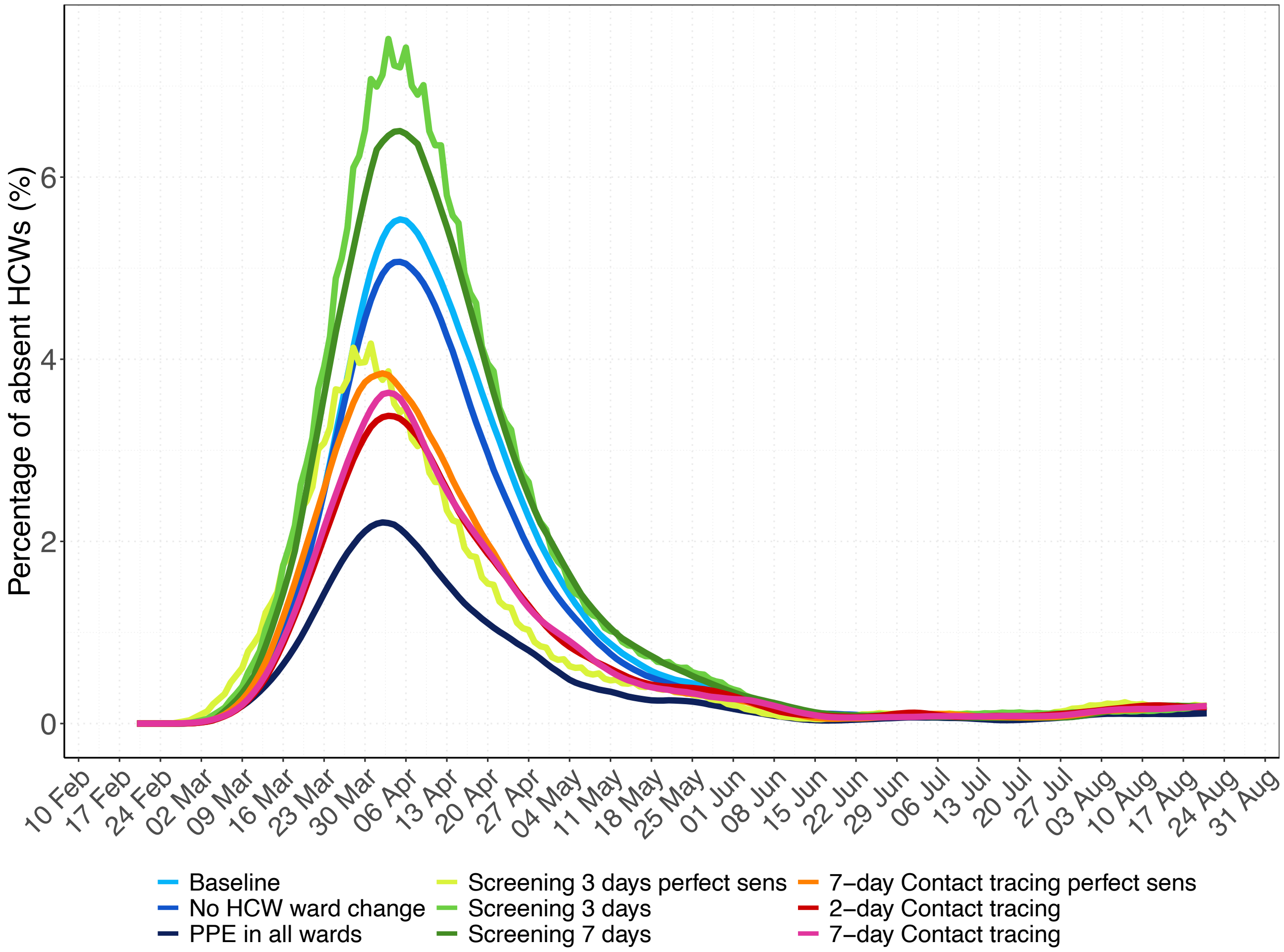
